# The association between the incidence of Lyme disease in the United States and indicators of greenness and land cover

**DOI:** 10.1101/2022.06.27.22276972

**Authors:** Sydney Westra, Mark S. Goldberg, Kamel Didan

## Abstract

**Background:** Lyme disease is the most common vector-borne illness in the United States. Incidence is related to specific environmental conditions such as temperature, metrics of land cover, and species diversity.

**Objective:** To determine whether greenness, as measured by the Normalized Difference Vegetation Index (NDVI), and other selected indices of land cover were associated with the incidence of Lyme disease in the northeastern USA, 2000-2018.

**Materials and Methods:** We conducted an ecological analysis of incidence rates in counties of 15 “high” incidence states and the District of Columbia for 2000-2018. Annual counts of Lyme disease by county were obtained from the US Centers for Disease Control and values of NDVI were acquired from the Moderate Resolution Imaging Spectroradiometer instrument aboard Terra and Aqua Satellites. County-specific values of population density, area of land and water were obtained from the US Census. Using quasi-Poisson regression, multivariable associations were estimated between the incidence of Lyme disease NDVI, land cover variables, human population density, and calendar year.

**Results:** We found that incidence increased by 7.1% per year (95% confidence interval: 6.8-8.2%). Land cover variables showed complex non-linear associations with incidence: average county-specific NDVI showed a ‘u-shaped” association, the standard deviation of NDVI showed a monotonic upward relationship, population density showed a decreasing trend, areas of land and water showed “n”-shaped relationships. We found an interaction between average and standard deviation of NDVI, with the highest average NDVI category, increased standard deviation of NDVI showed the greatest increase in rates.

**Discussion:** These associations cannot be interpreted as causal but indicate that certain patterns of land cover may have the potential to increase exposure to infected ticks and thereby may contribute indirectly to increased rates. Public health interventions could make use of these results in informing people where risks may be high.

## Introduction

The US Centers for Disease Control (CDC) reports that there are more than 300,000 cases of Lyme disease each year in the United States, making it the most prevalent vector-borne disease (CDC, 2019). Lyme disease is transmitted through the bite of a tick that has been infected with *Borrelia burgdorferi* spirochetes (CDC, 2019). In the Northeastern United States, *Ixodes scapularis*, commonly referred to as the black-legged tick, is the most abundant vector for Lyme disease (CDC, 2019).

Ticks become infected with *Borrelia burgdorferi* by feeding on an organism infected with this bacterium. Small rodents are the primary sources of infection, with the white-footed mouse being the principal reservoir in eastern North America (Allan et al., 2003). Other species of mammals and birds may also act as reservoirs, such as eastern chipmunks and American Robins (Mather, 1993). An infected larval tick will then molt into a nymph and transmit the bacterium in its next blood meal. Lyme disease is seasonal, with high rates of infections occurring in spring and summer, especially during the first week of July (Schwartz et al., 2017). The life stage of the tick occurs in a specific temperature range (Alonso-Carné et al., 2015).

One expects an increasing trend in the abundance of ticks, and hence of the incidence of Lyme disease, from urban areas to lower population density areas. Those areas with a mixture of habitats that include forest, various types of open and mixed areas, as well as wildland-urban interfaces should have lower densities of ticks (Diuk-Wasser et al., 2021). Wood and Lafferty (2013) suggested that biodiversity played an important role in infections when comparing rates in urban, semi-urban, to rural areas but were not convinced of the effect of biodiversity within forests (Wood and Lafferty, 2013). They also suggested non-linear associations between incidence and land cover whereby incidence increased with the degree of forest cover until a certain level of biodiversity becomes protective (“n-shaped curve”).

Metrics of land cover may be interpreted as indirect measures of the density of infected ticks (Kotchi et al., 2021) but may also represent the potential for human exposures to infected ticks, and so it is plausible that potential exposures would increase with increasing density of green landscapes including forests (Diuk-Wasser et al., 2021; Kilpatrick et al., 2017; Wood and Lafferty, 2013). Infections may also occur in urbanites when they travel to areas of higher densities of infected ticks, but infections can also occur in urban areas (Hamer et al., 2012; Jobe DA, 2007; VanAcker et al., 2019).

Remote sensing has been proposed as a tool for assessing vegetation cover (Gabriele-Rivet et al., 2017). In particular, the Normalized Difference Vegetation Index (NDVI) (Huete et al., 2002; Tucker et al., 2005) provides a measurement of vegetation canopy “greenness” (Myneni et al., 1995), as it measures a combination of leaf chlorophyll, leaf area, canopy cover, and structure. This is quantified through the ratio of the absorption of red light (R) by photosynthesis and reflectance of near-infrared by leaves (Tucker, 1979). NDVI is calculated as (NIR – R)/ (NIR + R). Values of NDVI range from -1.0 to +1.0, where very low and negative values indicate an absence of vegetation, as found in areas of barren soils, snow, or water, and high values correspond to dense vegetation, as found in temperate or tropical forests (Trishchenko et al., 2002).

With the considerations above, our objective was to determine whether the incidence of Lyme disease in the northeastern part of the United States was associated with NDVI and other metrics of land cover.

## Materials and Methods

### Incidence of Lyme disease

Counts of new cases of Lyme disease by county were obtained from the Centers for Disease Control for the years 2000-2018 (CDC, 2019). Surveillance of Lyme disease was established in the early 1990s, but we used counts starting in 2000 because this was the first year of available remote sensing data from MODIS (Justice et al., 2002). Since 1991, local and state health departments are required to report confirmed cases to the CDC (Schwartz et al., 2017).

Over 95% of all Lyme disease cases in the United States occur in the 15 states classified as having high incidence rates (at least 10 confirmed cases per 100,000 persons for the prior three reporting years) (CDC, 2019). We thus included in our analyses Connecticut, Delaware, Maine, Maryland, Massachusetts, Minnesota, New Hampshire, New Jersey, New York, Pennsylvania, Rhode Island, Vermont, Virginia, West Virginia, Wisconsin, and the District of Columbia.

### Normalized Difference Vegetation Index (NDVI)

We used high fidelity preprocessed NDVI time series available at the Climate Modelling Grid at a resolution of 0.05 degrees (Didan and Barreto, 2016a). NDVI data covering the period (2000-2016) was acquired from the Land Processes Data Active Archive Center (Didan and Barreto, 2016a) from the Moderate Resolution Imaging Spectroradiometer (Shu et al., 2020). MODIS is an instrument on board both Terra and Aqua satellites, operated by NASA since 2000 (Justice et al., 1998). MODIS acquires quasi-daily multispectral surface reflectance images of the Earth at 250 m, 500 m, and 1 km resolutions.

The full NDVI time series is a 35 years long multi-sensor NDVI record at 5.6 km resolution (Climate Modelling Grid derived from the remote sensing instruments AVHRR (1981-1999), SPOT (1998-2002) and MODIS (2000-2016) (Didan and Barreto, 2016b). This high-fidelity time series is free from spatial gaps that result from clouds and atmospheric contaminants, like heavy aerosols We obtained the NDVI time series from the Vegetation Index and Phenology Laboratory at the University of Arizona.

This time series was generated from the finer 250 m resolution MODIS daily observations. Surface reflectance observations were calibrated, corrected for atmosphere attenuation, aerosols, and screened for clouds (Justice et al., 1998). The daily surface reflectance were then filtered, refined and composited into multiday value-added measures of vegetation greenness (Huete et al., 2002; Huete et al., 2010), such as NDVI, and any remaining spatial gaps resulting from cloud screening were filled-in using a simple moving window regression or with long term average data if the gaps were persistent for long periods of time (Didan and Barreto, 2016b). These resulting high fidelity data records are used for long-term vegetation-climate interactions, detecting changes, modelling, and other derivative studies like public health (Jamison et al., 2015; Tourre et al., 2008).

The resulting NDVI images provide a spatially explicit measure of vegetation canopy greenness, health, and productivity over space and time (Myneni and Williams, 1994) and serves as a proxy for many structural and functional ecosystem parameters (Krishnaswamy et al., 2009).

We only considered the month of July in this study as it corresponds to the time of high ecosystem productivity and peak incidence of Lyme disease (Schwartz et al., 2017). For July of each year, we spatially aggregated the pixel data at the county level and generated multiple statistical parameters that represent the distribution of NDVI within the county. (Supplement Figure S1 shows the geographical distribution in the continental USA of maximum NDVI for July 2000.)

Because the NDVI data record stops at 2016 and the CDC Lyme disease data ends in 2018, we extended the NDVI record through years 2017 and 2018 using a simple regression model for average and the standard deviation of NDVI. This latter statistic may be interpreted as indicating fragmented areas of greenness within a county. This prediction model uses a county-specific linear regression in which we regressed the natural logarithm of NDVI versus time to generate predicted county-specific values for the missing two years.

### County-specific characteristics

Population counts at the county level were obtained from the US Census (Census, 2020) downloaded as county populations from 2000-2010 and 2010-2018. The population for the year 2010 was used from the county population 2010-2018 dataset, as new estimates incorporated the most up-to-date data for this year. In addition, we obtained from the census annual estimates of total land area, area of water, and we computed population density (Census, 2020).

## Statistical Analysis

Counts of Lyme disease were not identified by any personal characteristics, such as age and sex. As the outcome data are counts (or equivalently rates), the statistical model should follow approximately a Poisson distribution (Cameron and Trivedi, 2013). We thus used generalized linear models (McCullagh and Nelder, 1989) in which the natural logarithm of rates (or counts with a fixed offset for the logarithm of the population) was regressed against a linear combination of covariables. The counts were over-dispersed, so we used quasi-likelihood Poisson and negative binomial models to account for non-Poisson variability.

The variables included in the models were calendar year, population density, total land area, area of water, average NDVI, and the standard deviation of NDVI. These variables were computed for each year from 2000-2018. Weather variables were not included because we had annual data of counts of Lyme disease.

We made use of natural cubic splines to model the functional form of each covariable. These smoothers have a tunable parameter (number of knots or degrees of freedom, df) that allows for a different level of smoothness, with a higher number of knots or degrees of freedom allowing for more variability (less smoothing). We used the following algorithm to select the degrees of freedom: we modelled each covariable separately, adjusting for calendar year, and investigated the pattern of the graphs from 2 df to 6 df. The final value for the number of df was assessed visually by comparing the graphs and selecting the lowest df that revealed the pattern without having unnecessary variability that would likely be due to random fluctuations. We then modelled all of the covariables together, and this is the main result of the analysis where we present graphs of marginal effects (Lüdecke, 2018).

As well, because the patterns were complex, to provide quantitative estimates of rates, we computed rate ratios between selected cut points for each covariable, adjusting for all of the other covariables (Cao et al., 2006).

### Sensitivity analyses

We also conducted sensitivity analyses using various cut-points of the population of each county, and we conducted separate analyses of each of the 14 states (the District of Columbia was excluded because it comprises only one area).

We investigated interactions between selected variables, and we assessed these interactions using the likelihood ratio test as well as through fitted marginal graphs. As it is exceedingly difficult to graph the interaction of two nonlinear functions, we categorized one variable and then fit the fully adjusted model on the subset of data points defined by the different values of the categorical variable.

In addition, because there are reports of under-reporting of cases in many states (Rutz et al., 2018; Schiffman et al., 2018; Schwartz et al., 2017; White et al., 2018) we conducted a number of additional analyses excluding certain states as well as certain years.

## Results

Table 1 shows the distribution of the variables included in the analysis (2000-2018 for the selected states). The area of some counties was too small to measure the variability of NDVI, and thus these counties were excluded from the substantive analyses (193 observations).

**Table 1:** Distribution of county-specific values of counts and rates of Lyme disease, NDVI and selected other variables, 2000-2018 (580 counties, 15 states, 11,020 observations)^1^.

|  | Minimum | 1<sup>st</sup><br>Quartile | Median | Mean | 3<sup>rd</sup> Quartile | Maximum | Standard<br>deviation |
| --- | --- | --- | --- | --- | --- | --- | --- |
| Number of cases of Lyme disease | 0 | 1 | 6 | 46.1 | 40 | 1720 | 104.1 |
| Rates of Lyme disease (per 100,000) | 0 | <0.1 | 11.6 | 40.2 | 48.4 | 1,581.2 | 73.2 |
| Total population | 2,196 | 20,368 | 44,324 | 141,210 | 122,142 | 2,611,232 | 271,081.4 |
| Population density (per km<sup>2</sup>) | 1.0 | 14.9 | 32.0 | 290.9 | 117.3 | 27,681.8 | 1,480.5 |
| Average NDVI | 0.22 | 0.75 | 0.81 | 0.78 | 0.85 | 0.92 | 0.10 |
| Standard deviation of NDVI | <0.01 | 0.03 | 0.05 | 0.06 | 0.08 | 0.33 | 0.04 |
| Total land area (km<sup>2</sup>) | 19.4 | 914.6 | 1,353.5 | 1,652.0 | 2,021.1 | 17,278.0 | 1,515.2 |
| Area of water (km<sup>2</sup>) | 0.05 | 9.8 | 31.2 | 197.7 | 98.9 | 4,890.6 | 520.6 |
| Rural area (km<sup>2</sup>) | 0.0 | 792.0 | 1,231.9 | 1,537.0 | 1,881.4 | 17,253.8 | 1,534.6 |

<sup>1</sup> The following eight counties were excluded from the variance, standard deviation, 25%, 50%, 75% of NDVI: Charlottesville City, VA; Colonial Heights City, VA; Frankline City, VA; Fredericksburg City, VA; Galax City, VA; Hopewell City, Va; Martinsville City, Va; and Winchester City, VA.

The counties included in our analyses varied dramatically in area and population density and this type of heterogeneity is a cardinal characteristic of this administrative unit. The ratio of the population size between the largest to the smallest county was 1,200 and that of the total land area was 78. Rates of Lyme disease (per 100,000 persons) varied from 0 to 1,581.2 with a mean of 40.2 and standard deviation of 0.73. Counts of Lyme disease did not follow a Poisson distribution: the mean number of incident cases was 46.1 (range from 0 to 1,720) and the variance was 10,837 (ratio of variance to mean of 235), implying considerable overdispersion (Table 1).

Average county specific NDVI also varied considerably, from a very low greenness of 0.22 to a maximum of 0.92 (dense forest cover). There was also considerable variability in NDVI across counties, as shown by distribution of its standard deviation. The predictions of NDVI to 2017 and 2018 were similar to the observed values for 2000-2016 (Supplemental Figure S2).

We present results for the quasi-Poisson model, as the results with the negative binomial model were similar (see Supplement Figure S3). Figure 1 shows the fitted response curves (Lüdecke, 2018) for the model that included all variables. The estimate of the dispersion parameter in the quasi-Poisson model was 0.0012, close to the crude value. Calendar year was modeled using a linear function, the standard deviation of NDVI was modeled using two df, and all other variables were modelled using three df.

**Figure 1.**
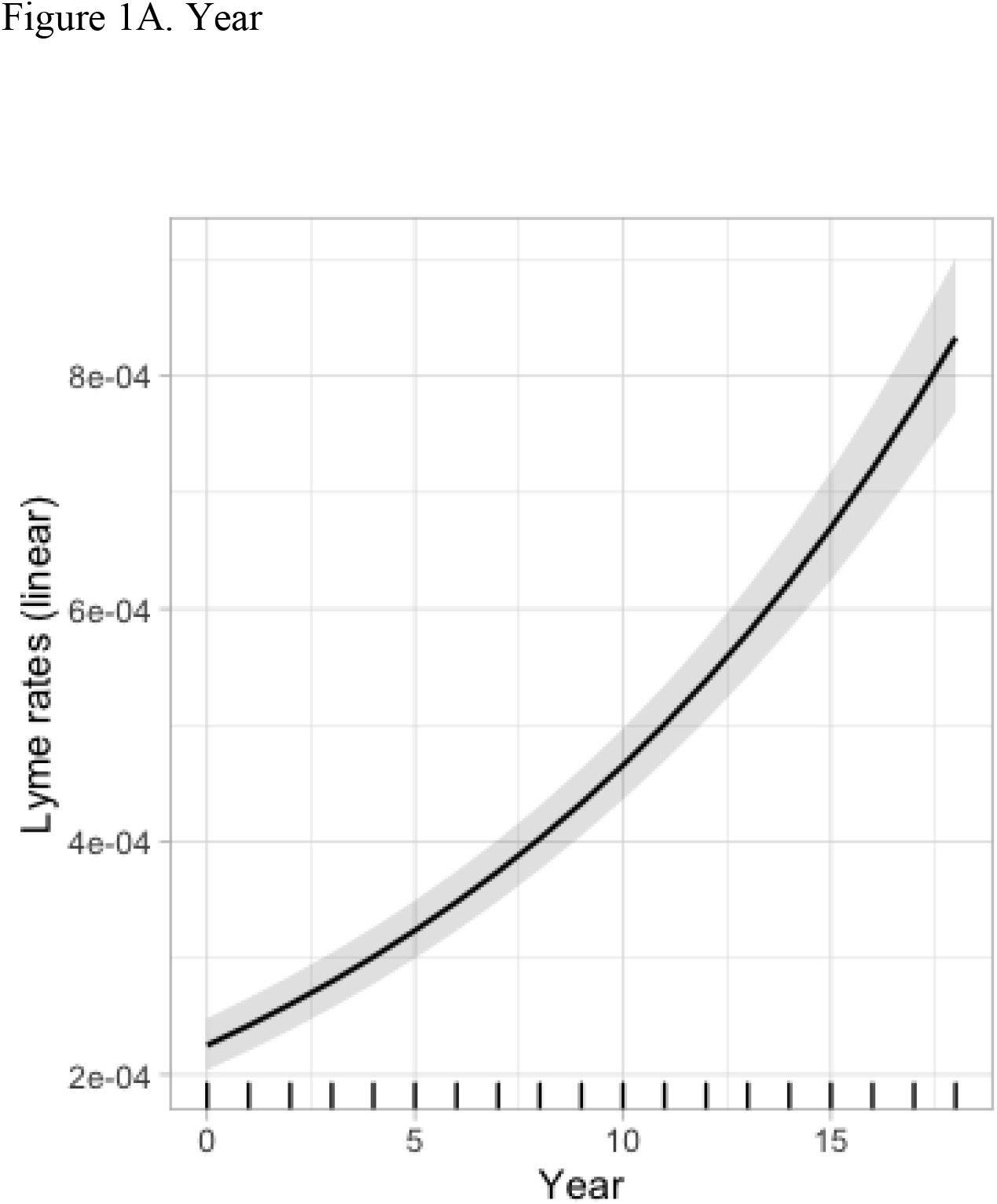
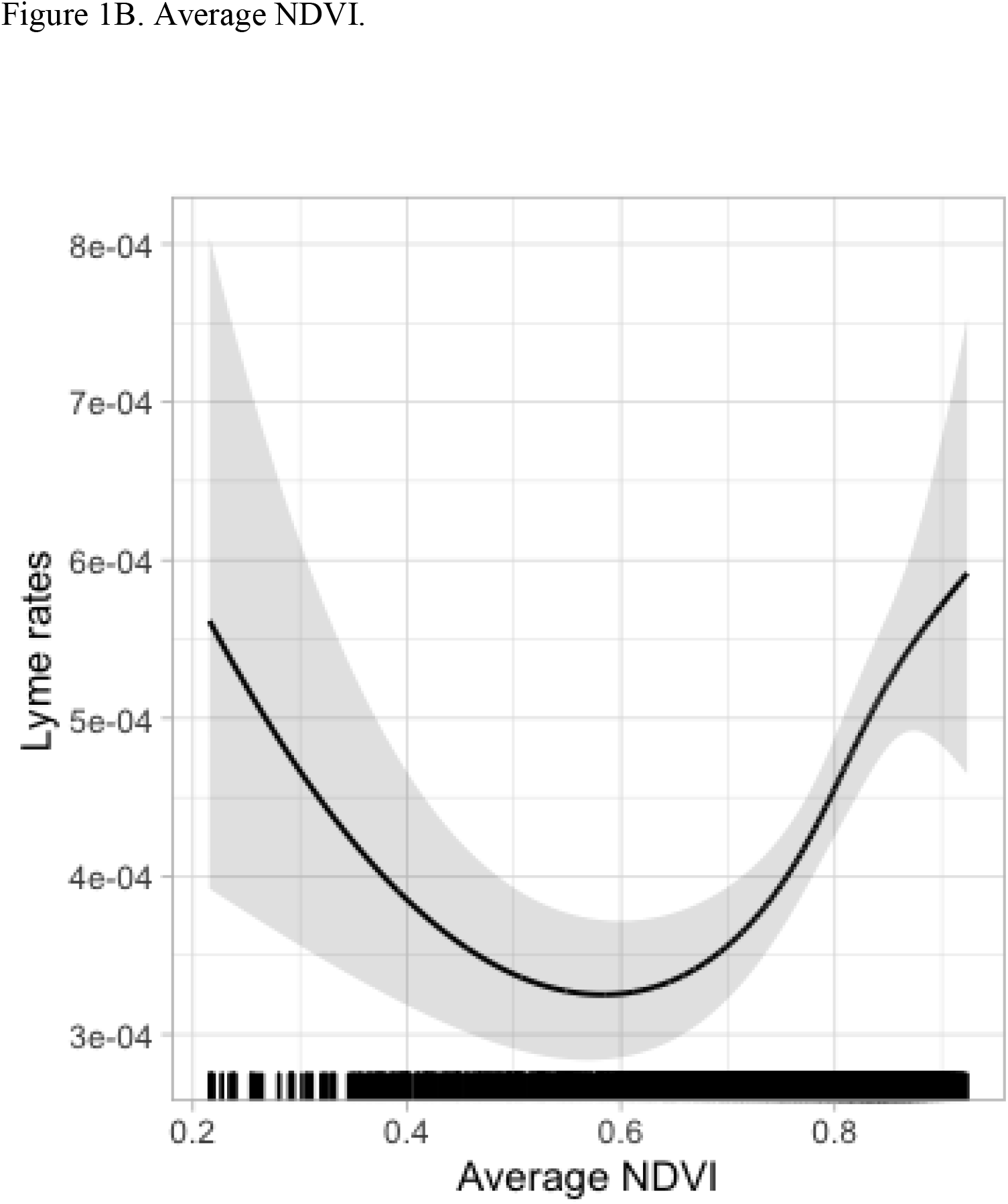
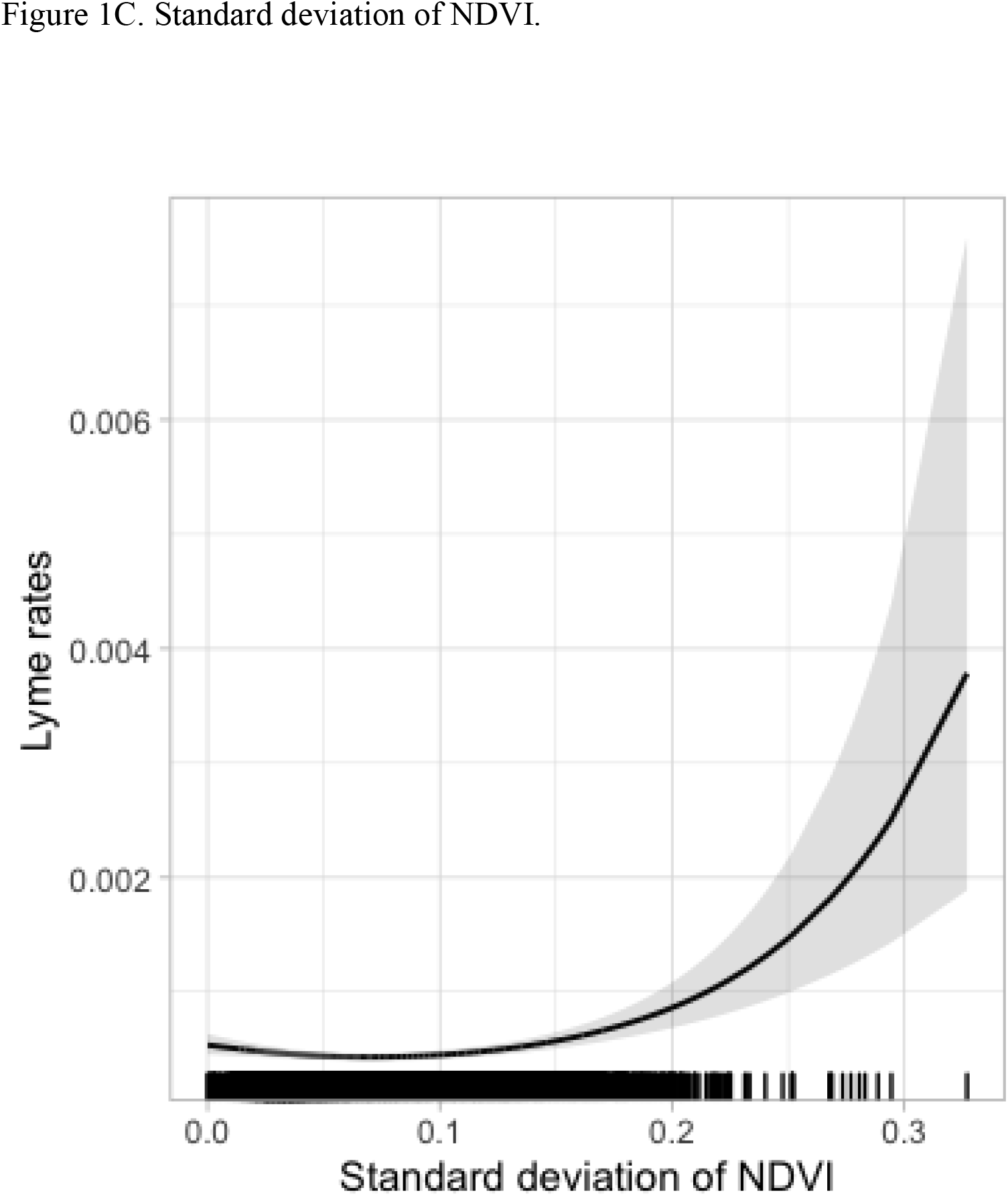
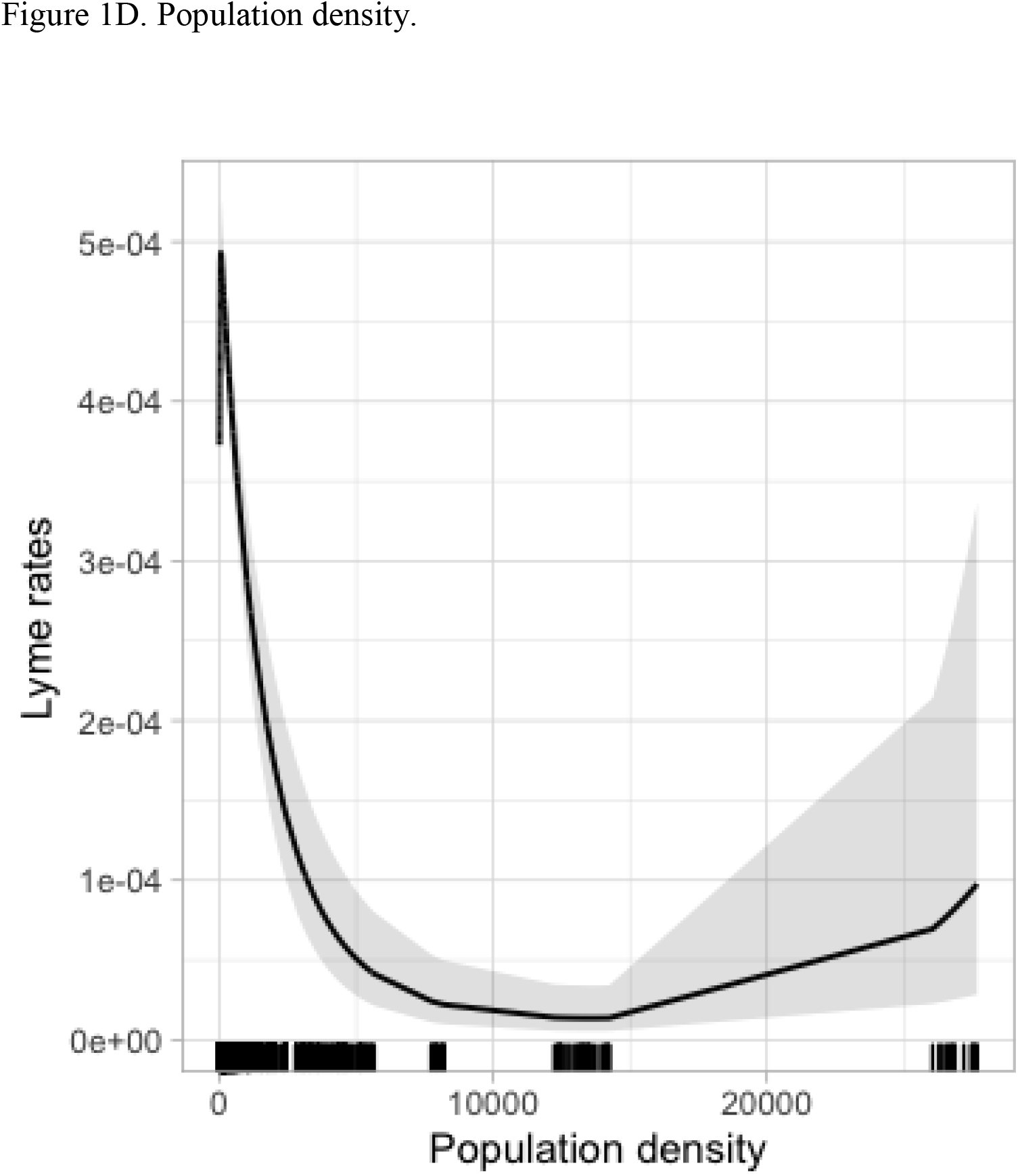
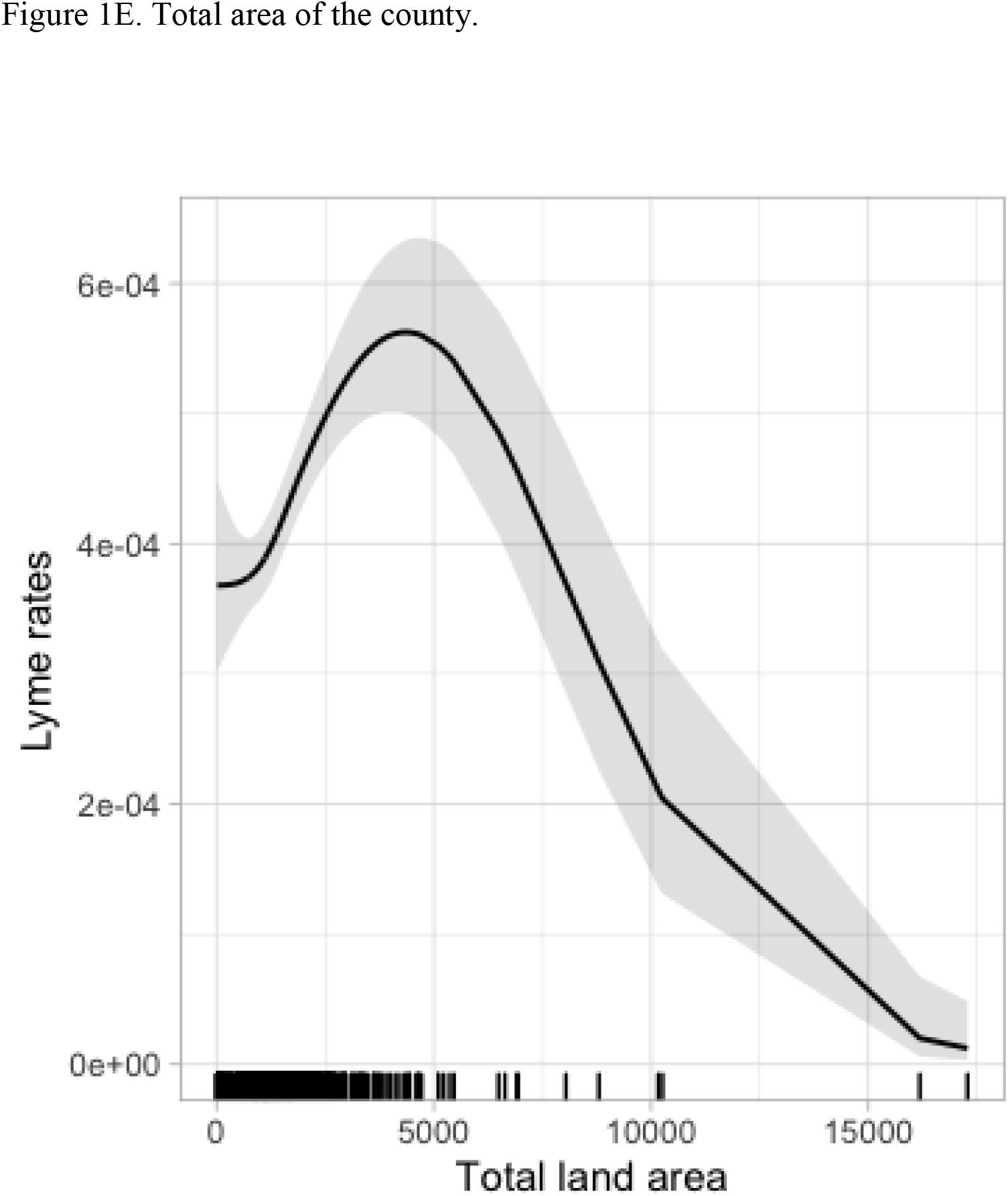
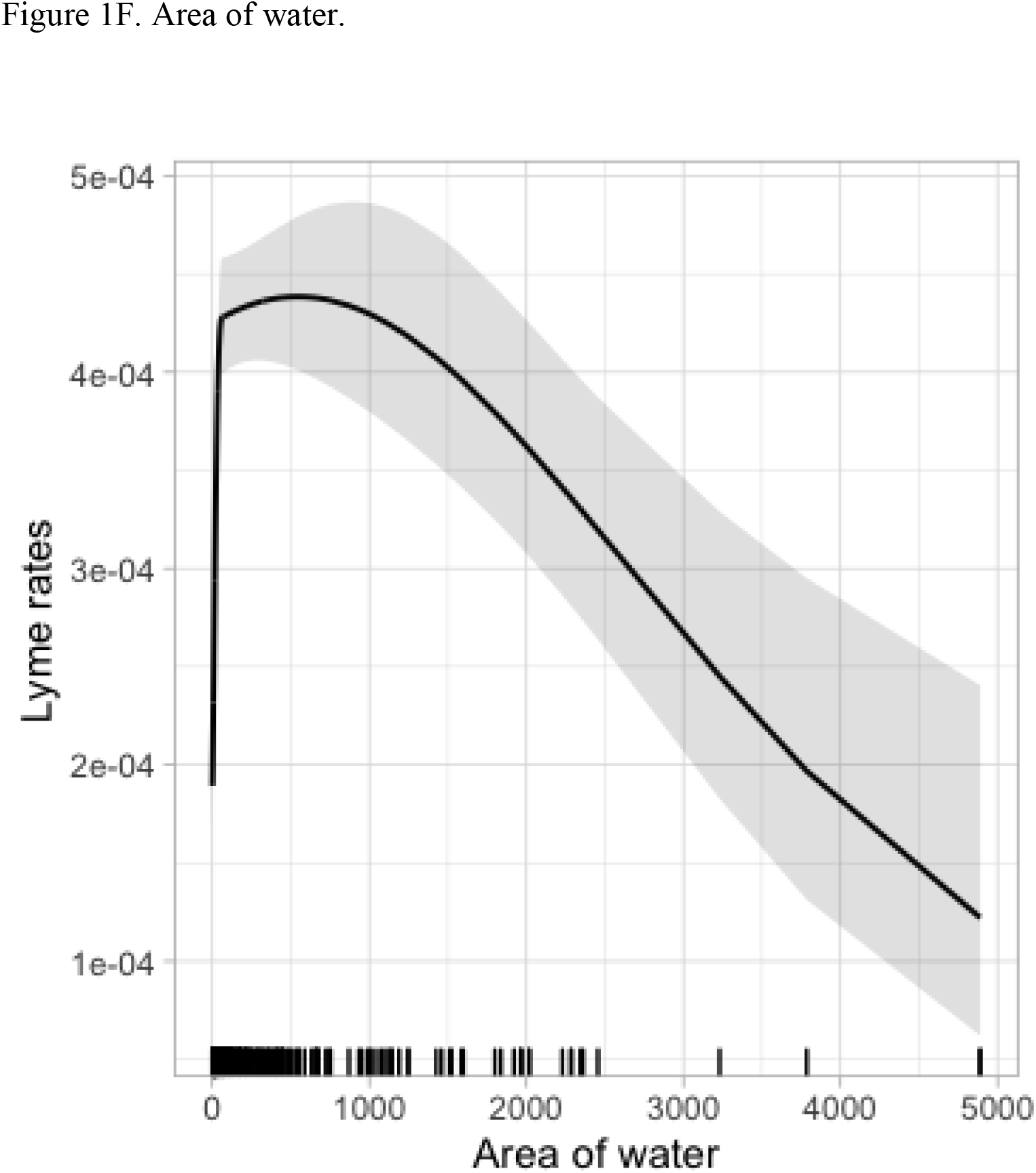
**Predicted rates of Lyme disease from a quasi-Poisson model that included year as a linear term, and natural cubic splines for average NDVI, the standard deviation of NDVI, population density, area of water, and total area of the county for the years 2000-2018, including 580 counties in 15 states (11,020 observations). Three degrees of freedom were used for all variables but standard deviation of NDVI, which took 2 df. The solid line represents the predicted value and the grey area surrounding the line represents the pointwise 95% confidence intervals. The rug plot along the x-axis shows where the observations lie.**

Figure 1A shows the predicted response function for calendar year, adjusted for all the other variables in the model. The grey bands represent pointwise 95% confidence intervals (CI) around the maximum likelihood estimate (solid line). The rug plot above the x-axis shows the placement of the values for year. In this plot, each tick on the rug plot represents a value for year, with each county contributing 19 values to the analyses. Although we fitted a linear term for year, the reason the graph shows some curvature is because the predicted values were computed by taking their exponent. The estimated annual increase in rates was 7.1% (95%CI: 6.8-8.2%).

Figure 1B shows the marginal response patterns for average NDVI where we found a non-monotonic function for rates of Lyme disease whereby adjusted rates decreased between about 0.2 to about 0.6 and then increased from 0.6 to 1. The rate ratio for an increase of average NDVI from 0.2 to 0.6 was 0.56 (95%CI: 0.38-0.82), showing that higher average NDVI values conferred lower rates of Lyme disease and the rate ratio from 0.6 to 0.85 was 1.60 (95%CI: 1.37-1.88), showing a 60% increase of rates of Lyme disease for this increment of average NDVI.

For the standard deviation of NDVI (Figure 1C), we found a monotonic increase in the rate ratio, with a modest change from 0 to 0.2 and then increasing more dramatically, although the confidence intervals were wide at the upper end. Namely, for greater variability in NDVI, related to land fragmentation from cities, water, roads, etc., the rate ratio between a value of 0 to 0.2 was 1.61 (95%CI: 1.25-2.07) and from 0.2 to 0.3 it was 3.12 (95%CI: 2.14-4.56).

Figure 1D shows the predicted marginal relationship for population density. Most counties had population densities under 10,000 per km^2^ with three clusters at higher values representing larger urban centers. These three clusters were due to only three counties and the different population densities in each cluster represented values for different years, hence the 95% confidence bands were very wide. The upward line at zero is an artifact of seven counties having very low population densities (<2 per km^2^). Rates decreased up to a population density of about 12,000 and then increased slowly. The rate ratio from a population density of 500 persons per km^2^ up to 10,000 was 0.04 (95%CI: 0.02-0.10) and from 10,000 to 30,000 it was 9.49 (95%CI: 1.84-49.02). Counties with the highest population densities such as New York County or Philadelphia County led to considerable uncertainty at the high end.

Figures 1E and 1F show the marginal response functions for total land area and area of water, respectively. Fifteen counties had a total area greater than about 5,000 km^2^ and 35 counties had areas of water greater than 1,000 km^2^. For total land area, we found another non-monotonic function increasing at the low values of the land area until about 5,000 km^2^ and then decreasing. The rate ratio from a minimum of 20 until 4,500 km^2^ was 1.53 (95%CI: 1.22-1.91) and from 4,500 to 10,000 km^2^ was 0.39 (95%CI: 0.26-0.58).

For area of water, rates increased at lower values and then dropped off rapidly; for example, from 10 to 100 km^2^ the rate ratio was 1.69 (95%CI: 1.56-1.83) and from 100 to 2000 km^2^ the rate ratio was 0.84 (95%CI: 0.72-0.99). Although there are few observations for larger areas of water, the decreasing trend covers the entire range.

### Sensitivity analyses

We investigated the interaction between average NDVI and the standard deviation of NDVI. The likelihood ratio test adding in the interaction term between these two variables indicated an important synergistic effect. We then categorized average NDVI into three groups (0-0.4, >0.4-0.6, >0.6-1) and we estimated the adjusted response curves for the standard deviation of NDVI for each separate category of average NDVI (Figure 2). Although we found a u-shaped response in rates of Lyme disease with increasing standard deviation of NDVI in the lowest category of average NDVI (0 to 0.4), the wide confidence intervals do not admit any meaningful interpretation. Figure 2 shows that rates for Lyme disease were constant in the second category of average NDVI (>0.4 to 0.6) and increased marginally in the third category of average NDVI (>0.6 to 1).

**Figure 2.**
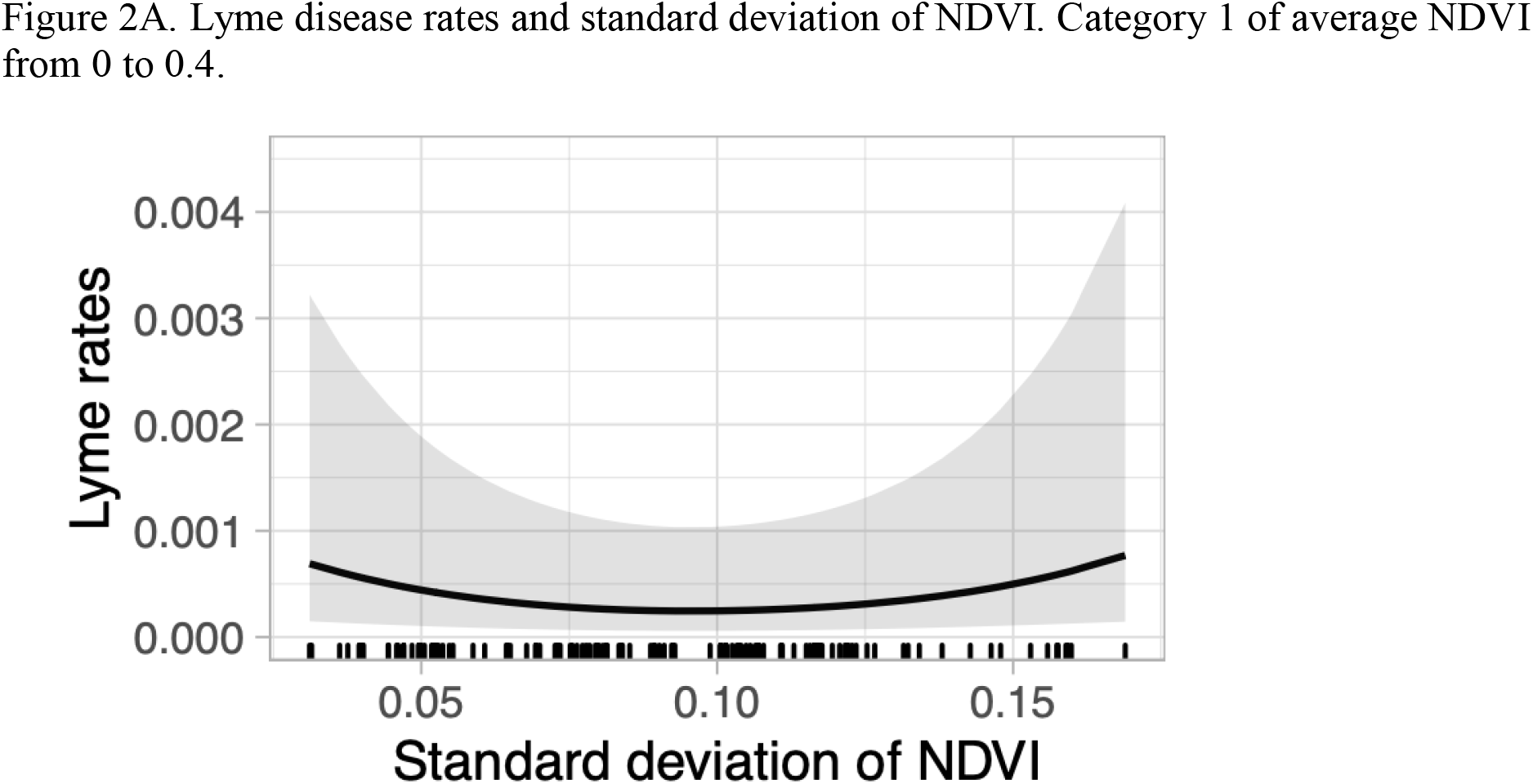
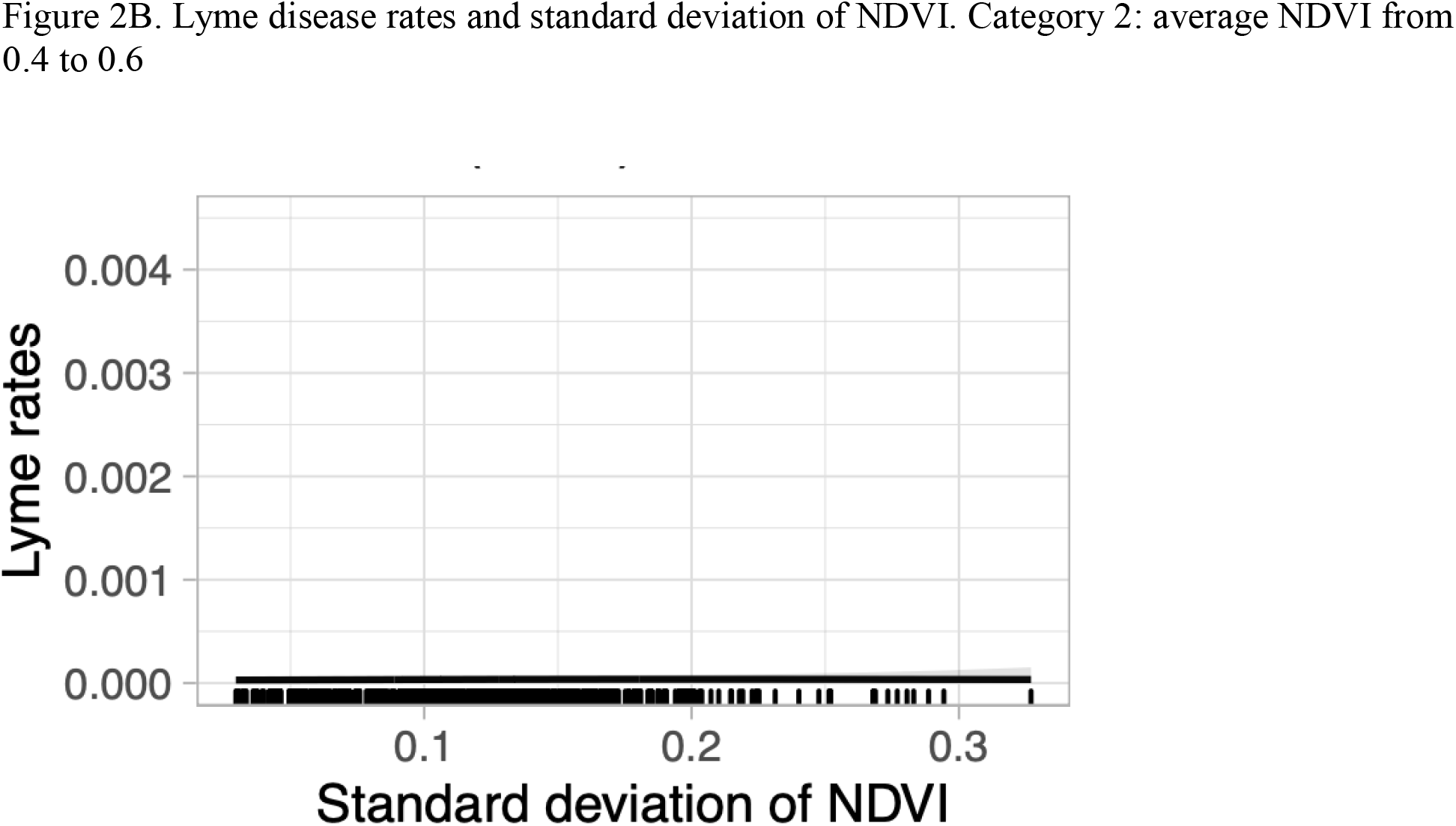
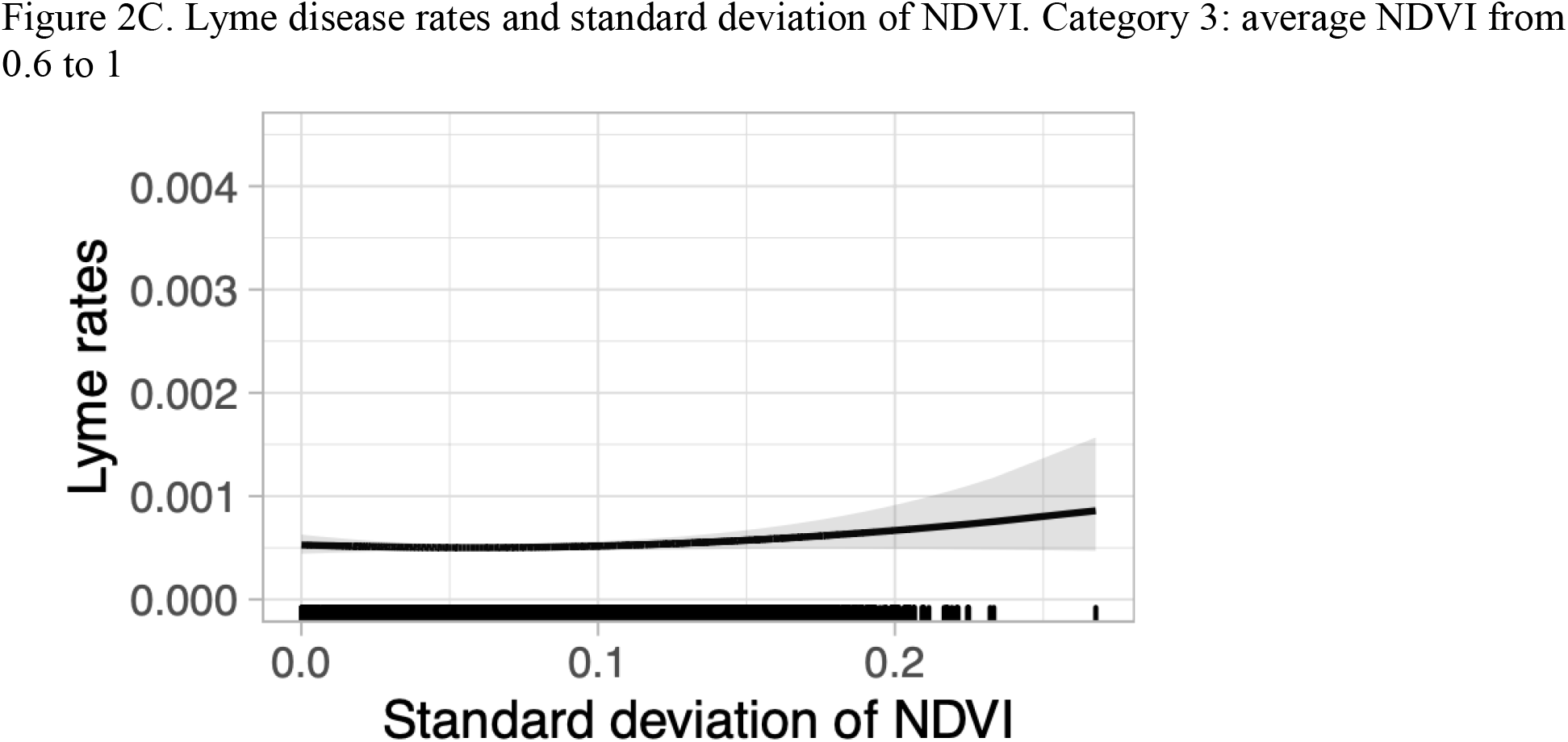
Interaction analysis of standard deviation of NDVI and rates of Lyme disease. Interaction between average NDVI and standard deviation of NDVI adjusted for other covariables. Average NDVI was split into 3 categories. Category 1 includes average NDVI from 0 to 0.4. Category 2 includes average NDVI from 0.4 to 0.6. Category 3 includes average NDVI from 0.6 to 1. For each category of average NDVI, the fitted response curves of the interaction analysis are shown.

We excluded counties with populations greater than 100,000 and populations greater than 381,000 but they exhibited functional forms and rate ratios comparable to the main analysis (Supplement Figure S4).

State-specific analyses for the two indices of NDVI are shown in the Supplement Figure S5. The patterns for NDVI differ between states and many states exhibit different patterns from the average shown above. The state-specific patterns are more difficult to interpret as there are few counties in each state, leading to considerable variability in these associations.

We conducted a series of separate analyses for different time periods in which the definitions used for Lyme disease changed, namely 2000-2008, 2008-2011, and 2011-2018. Because of the fewer numbers of observations, variability was increased but the general patterns found in the main analyses were similar (results not shown).

We also conducted analyses splitting the time period according to when public health departments began to experience difficulties in recording cases (around 2013) (Rutz et al., 2018; Schiffman et al., 2018; White et al., 2018) Figure 3 shows the response patterns for average NDVI split between the time periods 2000-2012 and 2013-2018. For the period prior to 2013 we found a flat response until around average NDVI of 0.8 (Figure 3A) whereas for the latter period we found higher rates below values of 0.4 and then subsequently a flat response for larger values (Figure 3B). We hypothesized that the change in pattern observed in this latter analysis may have been due to serious underreporting in certain states (New York, Maryland, Minnesota, and Connecticut) (Lukacik et al., 2018; Schwartz et al., 2017) so we excluded these states (Figure 3C) and indeed recovered the pattern seen in Figure 3A for the earlier period (2000-2012).

**Figure 3.**
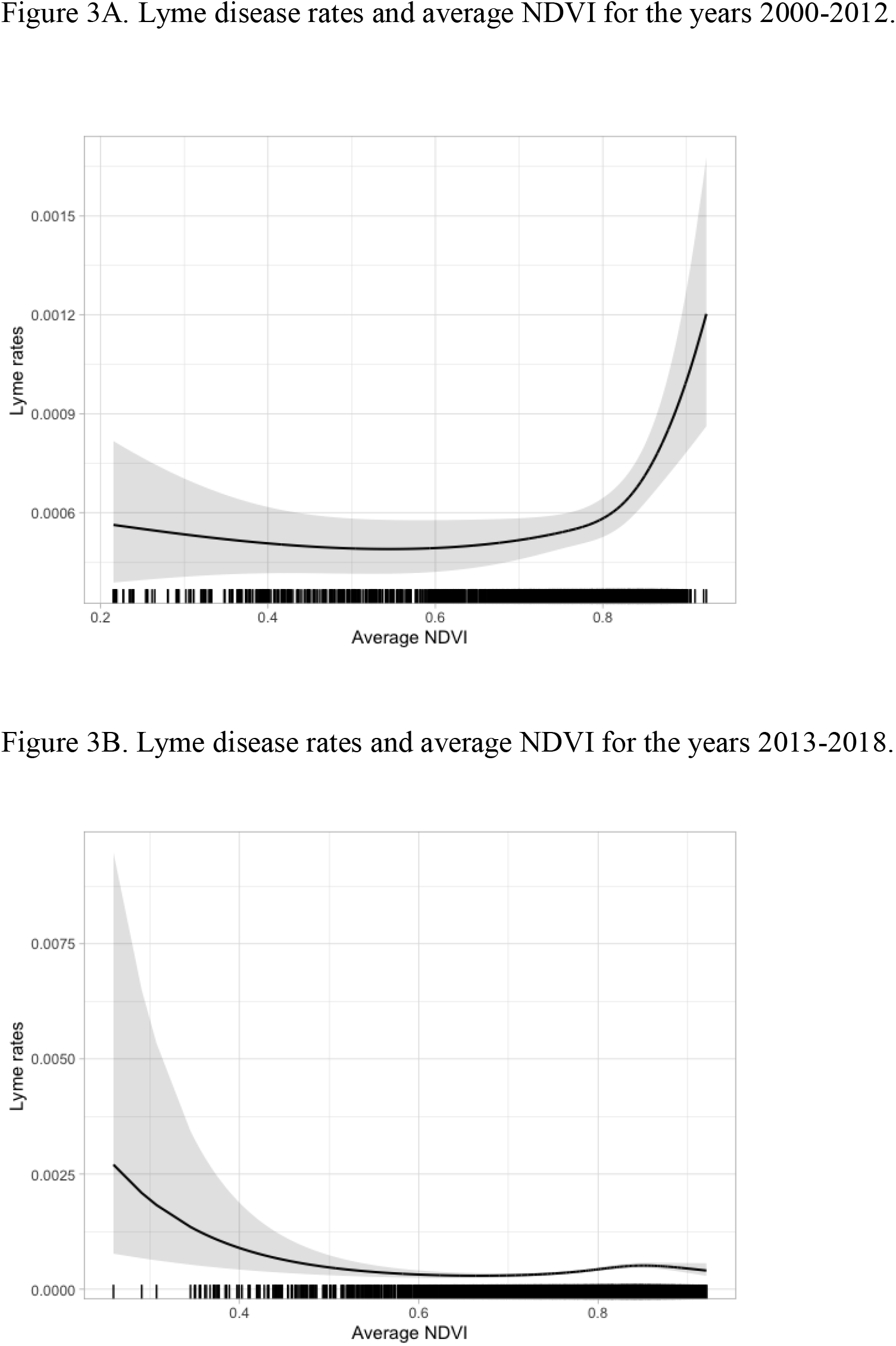
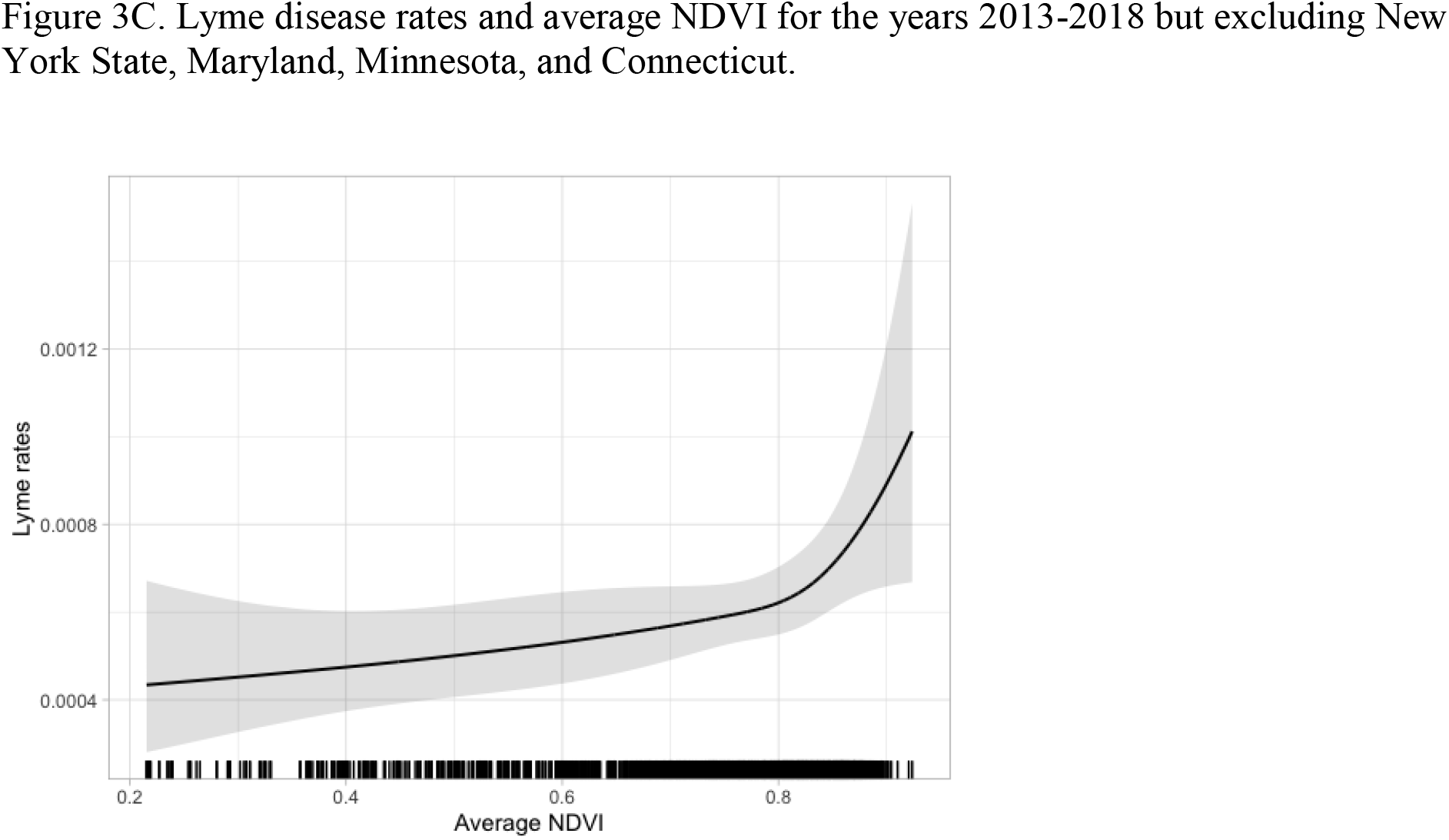
**Subgroup analyses of the associations between rates of Lyme disease and average NDVI and according to calendar year in which rates began to become underestimated (2013) as well as in different states.**

## Discussion

We found that the reported incidence of Lyme disease increased by 7.1% per year for the period 2000 to 2018, consistent with other reports (CDC, 2018; Kugeler et al., 2015; Murphree Bacon et al., 2008).

The definition of Lyme disease changed in the years 2008 and 2011. Before 2008, a confirmed case of Lyme disease was “a case with erythema migrans or a case with at least one late manifestation that is laboratory confirmed” (CDC, 2019). From 2008-2011, a confirmed case was defined as “a case of erythema migrans with a known exposure, or a case of erythema migrans with laboratory evidence of infection and without a known exposure or a case with at least one late manifestation that has laboratory evidence of infection” (CDC, 2019). From 2011-2017, the definition of a confirmed case remained the same, but the laboratory criteria for diagnosis became more specific as the presence of antibodies for *Borrelia burgdorferi* were used (CDC, 2019). Although these modifications were minor, these changes may have influenced the number of reported cases.

Part of the secular increase was due to improvements in reporting but also likely reflects increased rates of infection (Kugeler et al., 2015; Murphree Bacon et al., 2008). There has been under-reporting (Rutz et al., 2018; Schiffman et al., 2018; Schwartz et al., 2017; White et al., 2018) because public health departments could not deal with the volume of cases, healthcare providers could not order tests to detect Lyme disease and, thus, many cases were missed because the complete clinical information was not available. For example, (White et al., 2018) found that over a three-year period, three counties in New York State failed to report 20% of Lyme disease cases through standard surveillance. Our reported response functions could be biased if under-reporting was also associated with the variables in the model. The sensitivity analyses shown in Figure 3 indicated that this was the case, especially after 2013, but that the overall patterns for average NDVI that we found (Figure 1) likely are correct.

We found complex relationships in the functional forms of rates of Lyme disease and various indicators of greenness and land cover. We found that for increasing values of average NDVI from 0 to 0.6 lower rates of Lyme disease were observed at higher values of average NDVI, and for increasing values of average NDVI from 0.6-1.0 we found increases in disease rates. Low values of average NDVI, from 0.2-0.3 reflect areas in which there is relatively sparse vegetation (NASA, 2000). This may include areas such as urban centers, unvegetated areas, or sparsely vegetated meadows. For example, our data showed the average NDVI value for Kings County, NY, which comprises Brooklyn, NY, had an average NDVI value of 0.25 in 2002 which is comparable, for example, to desert-like regions.

Low values of average NDVI should imply that vegetation is largely absent, which should in theory translate to an absence of habitats for ticks and the species that carry ticks. Indeed, living in urbanized areas is typically thought to be protective of Lyme disease (Glass et al., 1995), but our model found the highest rates of Lyme disease in areas with little vegetation and areas with dense vegetation. Higher rates of Lyme disease in areas of little vegetation may be partly the result of travel-related behaviour, such as when a person acquires Lyme disease elsewhere, and the infection is reported in their urban/residential county. However, various studies have shown that most cases of Lyme disease are acquired around people’s homes, such as in one’s lawn or backyard (Falco and Fish, 1988; Maupin et al., 1991; Smith et al., 2001). Furthermore, our sensitivity analysis suggests that after excluding large cities we observed the same functional form, where the highest rates of Lyme disease were associated with the lowest and highest levels of average NDVI (Supplement Figure S4B). This suggests that travel-related behaviour may not fully explain the relationship between higher rates of Lyme disease in areas of little vegetation.

The incidence of Lyme disease in US cities is understudied and is likely more prevalent in cities than thought previously (Hamer et al., 2012; Jobe DA, 2007; VanAcker et al., 2019). VanAcker et al. (2019) found that 17 out of 24 parks in New York City contained the *Ixodes scapularis* tick, and of all the ticks sampled, an average of 26.6% of all ticks were infected with the Lyme disease bacteria. Cities contain a high density of people, making it especially important to be aware of the expansion of this zoonotic disease into cities. Additionally, lower to moderate levels of NDVI averaged in a county reflect a mixture of residential/urban areas and a small amount of forested area, such as the edge of forests. Many of these areas may be in proximity to forest edge habitats that may increase the interaction between disease vectors and humans and thus increase risk (Brownstein et al., 2005; Jackson et al., 2006a).

The diversity of species (DeLong, 1996) may affect the ecology and epidemiology of vector-borne zoonoses through a process called the “dilution effect” (Ostfeld and Keesing, 2000) which posits that a greater variety of species dilutes the proportion of infected rodents for ticks to feed on. A loss of biodiversity can indirectly increase risk of disease, as species carrying the Lyme disease bacteria are predominately small rodents that are better able to survive and adapt to loss and change of habitats. Although biodiversity is difficult to measure across large spatial scales, indirect measures that relate to land cover have been used (Bawa et al., 2002; Grantham et al., 2008). The hypothesized dilution effect implies that the greater diversity of species in communities may dilute the number of infected white-footed mice and as a result reduce the incidence of Lyme disease (Ostfeld and Keesing, 2000). Habitat fragmentation and destruction of forests can be an important driving force in the loss of many species in a community (LoGiudice et al., 2003). Numerous studies (LoGiudice et al., 2003) have referenced the dilution effect as a means in which biodiversity can ameliorate Lyme disease risk, but in few studies using direct metrics of biodiversity has this hypothesis been studied or confirmed (Wood and Lafferty, 2013).

As average NDVI increased in the range 0.6 to 0.8 (which corresponds to highly vegetated counties), higher rates of Lyme disease were observed. This finding is compatible with various studies in the Northeastern United States which suggest that the risk of Lyme disease increases with increasing forestation (Brownstein et al., 2005; Glass et al., 1995; Kitron and Kazmierczak, 1997). A review on spatial patterns and environmental correlates of human cases of Lyme disease from Killilea et al. (2008) found the only environmental variable consistently associated with increased Lyme disease risk was the presence of forests. However, possible non-linear relationships between forest coverage and Lyme disease were not assessed in the studies included in this review (Kitron and Kazmierczak, 1997; Orioski et al., 1998). Our model is consistent with previous findings where higher rates of Lyme disease were associated with greater forestation.

We also found increased rates of Lyme disease with higher values of the standard deviation of NDVI. Higher variability in NDVI reflects fragmentation of landscapes and/or the presence of large urban to natural interfaces. Landscape likely contributes to the incidence of Lyme disease through the reservoir species it supports, namely the white footed mouse (Brownstein et al., 2005; Falco and Fish, 1988). White footed mice can tolerate fragmented habitats while many other species cannot (LoGiudice et al., 2003). This is a possible explanation for higher disease rates in areas with a higher standard deviation of NDVI. As it has been found that most Lyme disease cases are contracted peri-domestically, the landscape can further contribute to Lyme disease risk through promoting the interaction between infected ticks and humans. An area with an increasing variation of NDVI may have a greater area of interface between residential property and forest edge. This is supported by several studies showing that fragmented landscapes are associated with either greater disease rates or entomological risk (Allan et al., 2003; Brownstein et al., 2005; Diuk-Wasser et al., 2021; Tran and Waller, 2013). It is likely important to consider educating people living in or interacting with areas of highly fragmented vegetation.

The interaction analysis (Figure 2) of average NDVI and standard deviation of NDVI suggests that the relationship between standard deviation of NDVI and rates of Lyme disease at low levels of average NDVI have too much variability to make conclusions. At moderate levels of average NDVI, increases in standard deviation of NDVI showed constant rates of Lyme disease. The highest levels of average NDVI shows slight increases of rates of Lyme disease with increasing standard deviation of NDVI, which is similar to the finding of the main analysis. This may suggest that fragmentation of forests within counties of high levels of NDVI have higher rates of Lyme disease, which may provide support of the dilution effect.

We found increased rates of Lyme disease with counties with lower population densities, compatible with a previous study (Seukep et al., 2015), and also consistent with our findings for average and the standard deviation of NDVI as one would expect that more sparsely populated areas would have more forested areas and a greater presence of infected ticks. There was a suggestion, however, of increased rates in high density populations that may be due to the presence of ticks in city parks or to people being infected while travelling. As Lyme disease continues to expand into cities, we may see a shift in this relationship between population density and Lyme disease rates.

Rates of Lyme disease increased with the greater total land area until 5000 km^2^. The size of the county has not been found previously to predict Lyme disease risk (Jackson et al., 2006b), but larger counties likely indicate a higher probability they harbor more infected ticks and hence higher rates of Lyme disease. For example, Maine has both the highest rates of Lyme disease and relatively large counties which contribute to the finding that the relationship between county area size and Lyme disease risk is greater. On the other hand, the largest counties exhibited rates that diminished with land area. The decrease in rates of Lyme disease in counties with land areas above 5,000 km^2^ occurred in 15 counties: six in Maine, eight in Minnesota, and one in New York State. Collectively, they have a median population of 15,000, a median population density of 6.1 per km^2^, and a rural area that is about 46% of the total. These lower rates is unlikely due to lower densities of *Borrelia burgdorferi* spirochetes (e.g., 50% of deer ticks tested positive in St. Lawrence County, NY) but could be due to populations protecting themselves better (County, 2019).

Finally, as water area increased, Lyme disease rates initially increased in smaller areas, and steadily decreased afterward. Naturally, in areas with large tracts of water, where there is sparse to no vegetation, there will be no viable habitat for the ticks or disease vectors, and as we expect, there may be lower rates of disease.

The strengths of this study include the utilization of the MODIS sensor that allowed us to include a long time series of observations in this analysis. These spaceborne platforms provide quasi-daily observations about the Earth’s surface and enable novel data fusion techniques and insights into how this disease is moderated by environmental factors. Our study considered only the month of July, when the vegetation is at its peak activities, peak habitat quality, while minimizing NDVI noise, like clouds, atmosphere contaminants, and maximizing illumination. Instruments like MODIS have been providing consistent and accurate measures, like NDVI, a proxy of vegetation health and productivity, at a resolution of 250 m. While we have used coarser spatial resolution NDVI data (5.6 km) that were derived from the native finer 250 m resolution observations making our estimates of county-specific variability reasonably accurate and robust. We note here that some counties were simply too small to estimate variability in NDVI and were omitted from the multivariable analyses.

We used standard statistical models for rates of diseases, namely the quasi-Poisson and negative binomial regression models that account for the variability in Lyme disease counts and rates, and both models yielded similar results. Moreover, we did not assume one value for any of the covariables, but allowed them to vary by year over the 19 years of observation, thereby reducing measurement error. Lastly, we did not assume any specific functional form for the associations between Lyme disease and the covariables, such as linear relationships, and by doing so we reduced bias from mis-specifying the regression model.

Our study is limited by the complex processes underlying the epidemiology of Lyme disease. As mentioned above, cases of Lyme disease are quantified based on their county of residence, which assumes that the person contracting Lyme disease will visit the medical centre in closest proximity to where they acquired Lyme disease. As well, where Lyme disease is endemic, it is frequently acquired at home from activities in the lawn or backyard (Connally et al., 2006; Falco and Fish, 1988). Our data, however, cannot be used to establish the place of infection. As well, we did not have information on their specific characteristics, such as age, sex, and income.

As the age distribution is bimodal (Mead, 2015), it is possible that different relationships by age may have been obtained that could be related to behavioural-related differences and age-related differences in susceptibility and care-seeking behaviors (Mead, 2015).

We focused only on land cover, the habitat of the vector and hosts, while numerous other factors contribute to the dynamics of the infection such as changes in temperature and human interactions within ecosystems. We did not account for temperature as we only had annual data for Lyme disease. As well, temperature does not vary dramatically between areas; for example, the mean maximum temperature in July in Maine is about 26°C and in Delaware it is 30°C.

Furthermore, we were only able to incorporate reported cases in the analysis, as Lyme disease is underreported and misdiagnosed in the United States (Rutz et al., 2018; Schiffman et al., 2018; Schwartz et al., 2017; White et al., 2018). The increased number of cases reported throughout our study period can be partly explained by true increases in rates, improved techniques to detect Lyme disease, and the changing definition of what constitutes a positive case of Lyme disease.

The present analysis contributes to the literature by confirming prior findings about Lyme disease dynamics while providing a simple framework for studying key parameters of the spatial dynamics of Lyme disease (Killilea et al., 2008). Our findings show clear non-linear relationships for many variables which may prove useful in designing future studies. One of the best-suited epidemiological designs for making causal inferences is the case-control study in which one can design detailed investigations of factors related to exposure in cases and appropriately selected controls (non-cases) (Connally et al., 2009; Orioski et al., 1998).

We found associations with land cover variables that can be utilized in predicting exposure and risk of Lyme disease and may suggest where it may be important to focus on prevention and early detection. Our results suggest that both urban areas (such as cities with large parks) along with suburban and natural settings may be critical areas of infection. We also found associations between increased rates of Lyme disease with higher values of the variability of NDVI within counties. Public health education programs would focus not just on rural areas but also on urban and suburban areas and those people traveling to natural settings in which there is the likelihood of infection, including those living in fragmented areas. Additionally, people may be less likely to be vigilant about Lyme disease infection whilst walking through a city park rather than those traveling to natural settings. As Lyme disease continues to expand into cities, future studies on Lyme disease should also have a strong focus on its dynamic in urban areas along with rural and natural settings.

## Conclusions

The relationships uncovered and reported in this analysis cannot be interpreted as causal, rather, they indicate that certain patterns of land cover may increase or decrease exposure to infected ticks and thereby may contribute indirectly to increased rates. Only properly conducted cohort or case-control studies can be used to make causal inferences. Generally, our study points to a complex relationship between the considered parameters and Lyme disease incidence rates, with the most critical being a segmented response to NDVI, a proxy of landscape vegetation greenness, showing a strong decrease of rates with increasing average NDVI (up to NDVI∼0.6), then a strong positive relation with rates increasing with increasing average NDVI. We suspect this divergent correlation could be capturing aspects of the structure of landscapes, in particular fragmentation and hence the quality of habitats. Furthermore, our study presented a simple framework to fuse remote sensing time series data and census data and the incidence of Lyme disease that were not explored before to elucidate how the distribution and trends in landscapes may be playing a role in moderating reservoirs for ticks and spread of disease.

## Data Availability

All data produced in the present study are available upon reasonable request to the authors

https://lpdaac.usgs.gov/products/vip15v004/

## Acknowledgments

This work was partially supported by NASA EOS-MODIS grant number 80NSSC18K0617 (K Didan, PI).

