## Supplement for "The association between the incidence of Lyme disease in the United States and indicators of greenness and land cover"

**The association between the incidence of Lyme disease in the continental United States and the Normalized Difference Vegetation Index and selected indicators of land use, 2000-2018**

**Sydney Westra, Mark S. Goldberg, Kamel Didan**

**Figure S1. The study area is the continental United States. This represents July maximum NDVI levels for the year 2000, aggregated by county. Dark green indicates NDVI levels near +1, red and orange indicate NDVI levels near 0. (Source: Didan, 2019)**

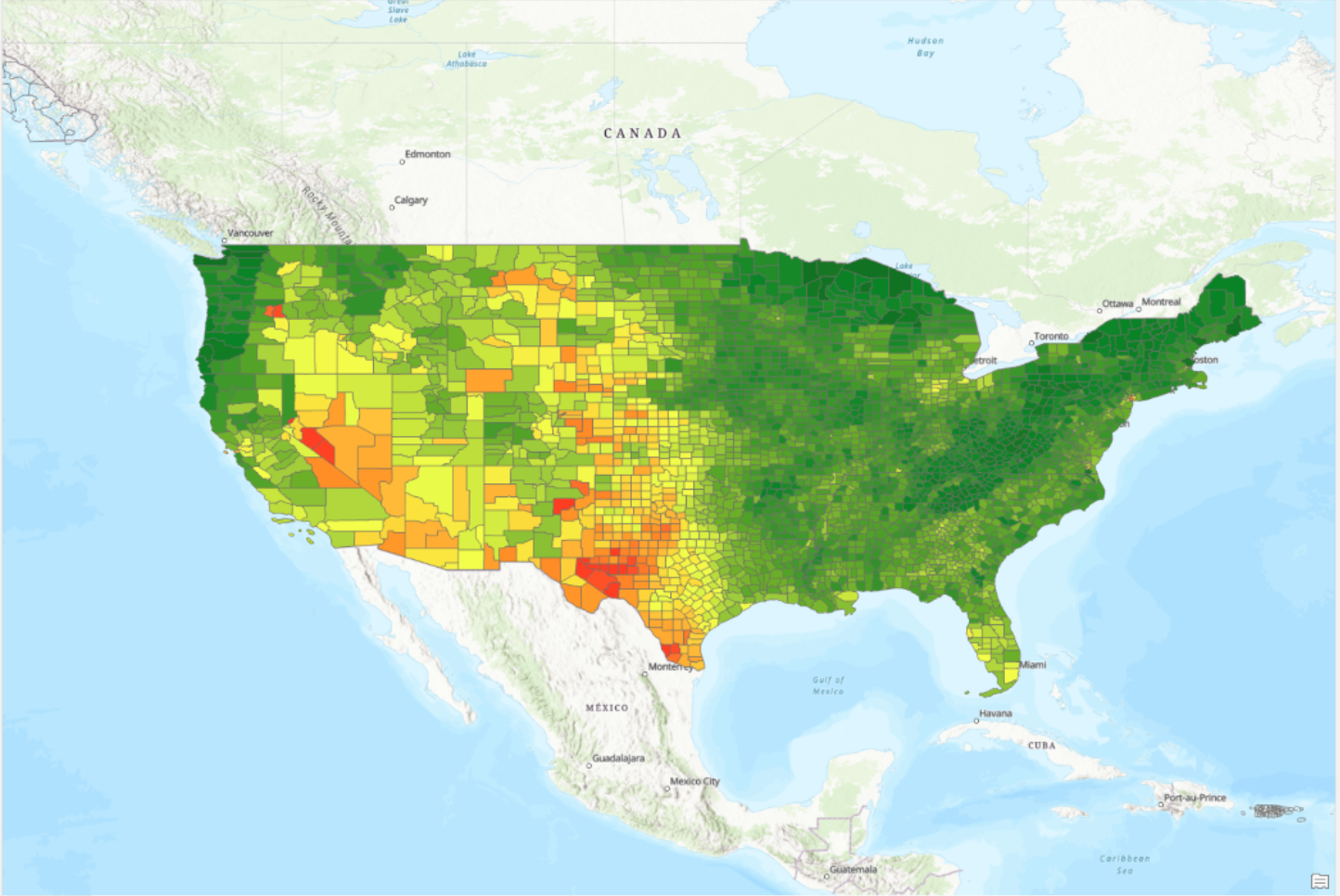

**Figure S2. Distributions of average and standard deviation of NDVI by year. The box “avg0016” and “stdev0016” represents the mean across years 2000 to 2016, “avg10” and “stdev10” represents that for the year 2010, and so on.**

Figure S2A. Average NDVI by Year

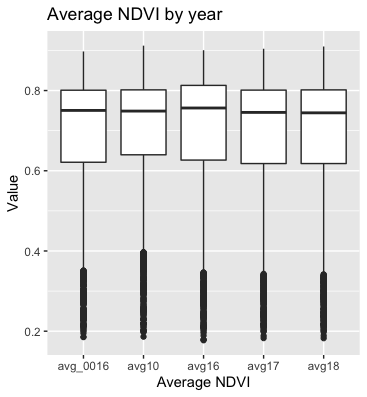

Figure S2B. Standard deviation of NDVI by year

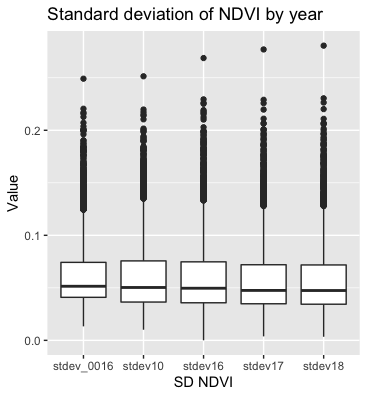

**Figure S3. Response functions using a negative binomial model. (The extra variance term (theta) from the negative binomial model was 0.48 (standard error was 0.007).)**

Figure S3A. Average NDVI

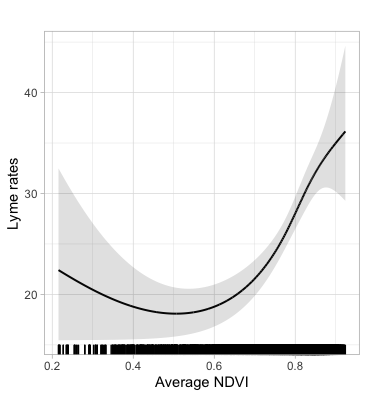

Figure S3B. Standard Deviation of NDVI

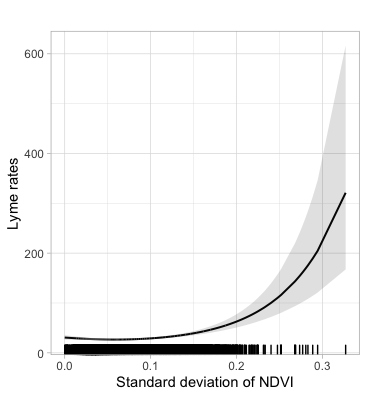

Figure S3C. Year

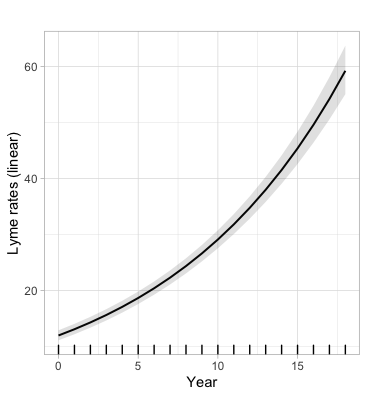

Figure S3D. Total Land Area

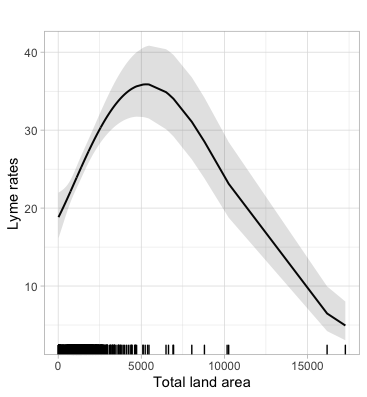

Figure S3E. Area of Water

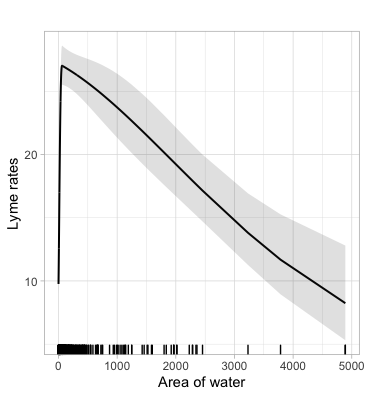

Figure S3F. Population density

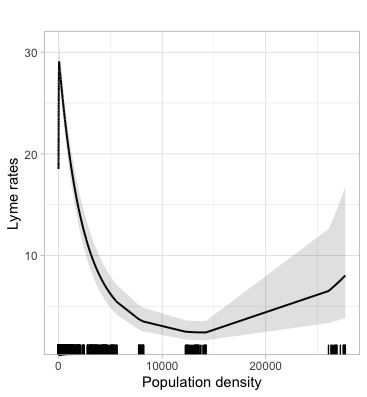

**Figure S4. Sensitivity analysis: Response functions** **from a quasi-Poisson model (that included all variables) for counties with populations under the 90^th^ percentile (381,000 people)**

Figure S4A. Year

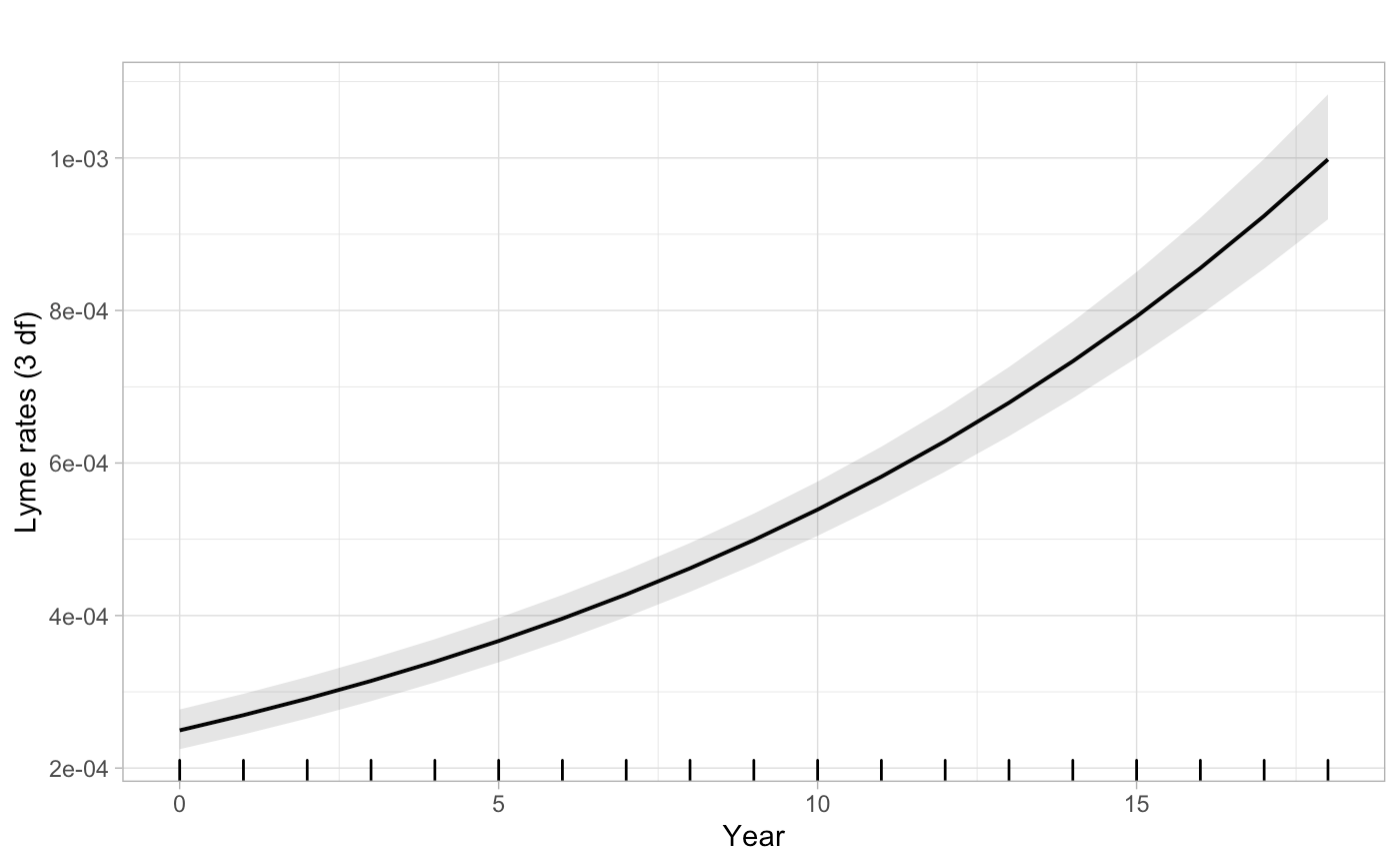

Figure S4B. Average NDVI

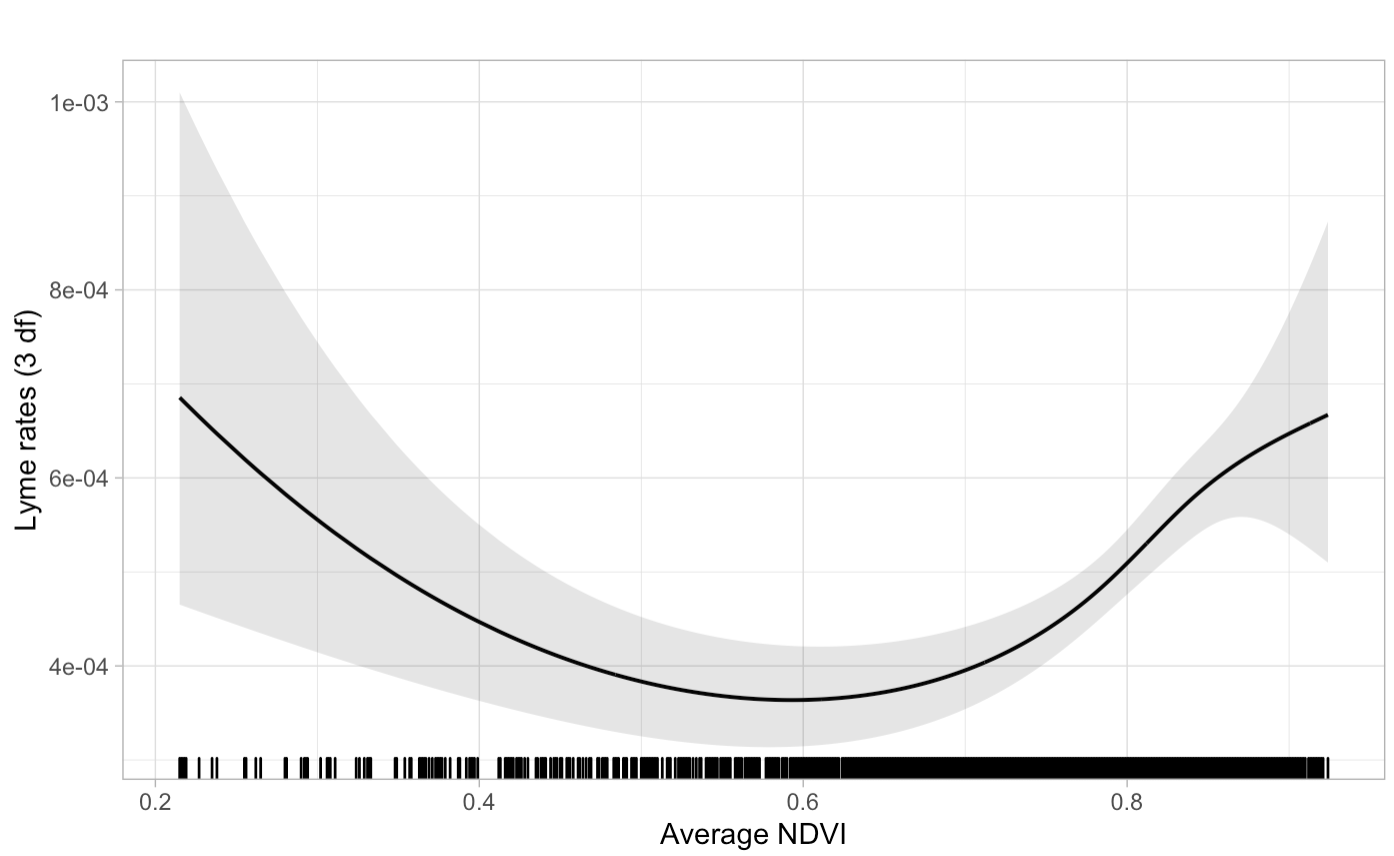

Figure S4C. Standard Deviation of NDVI

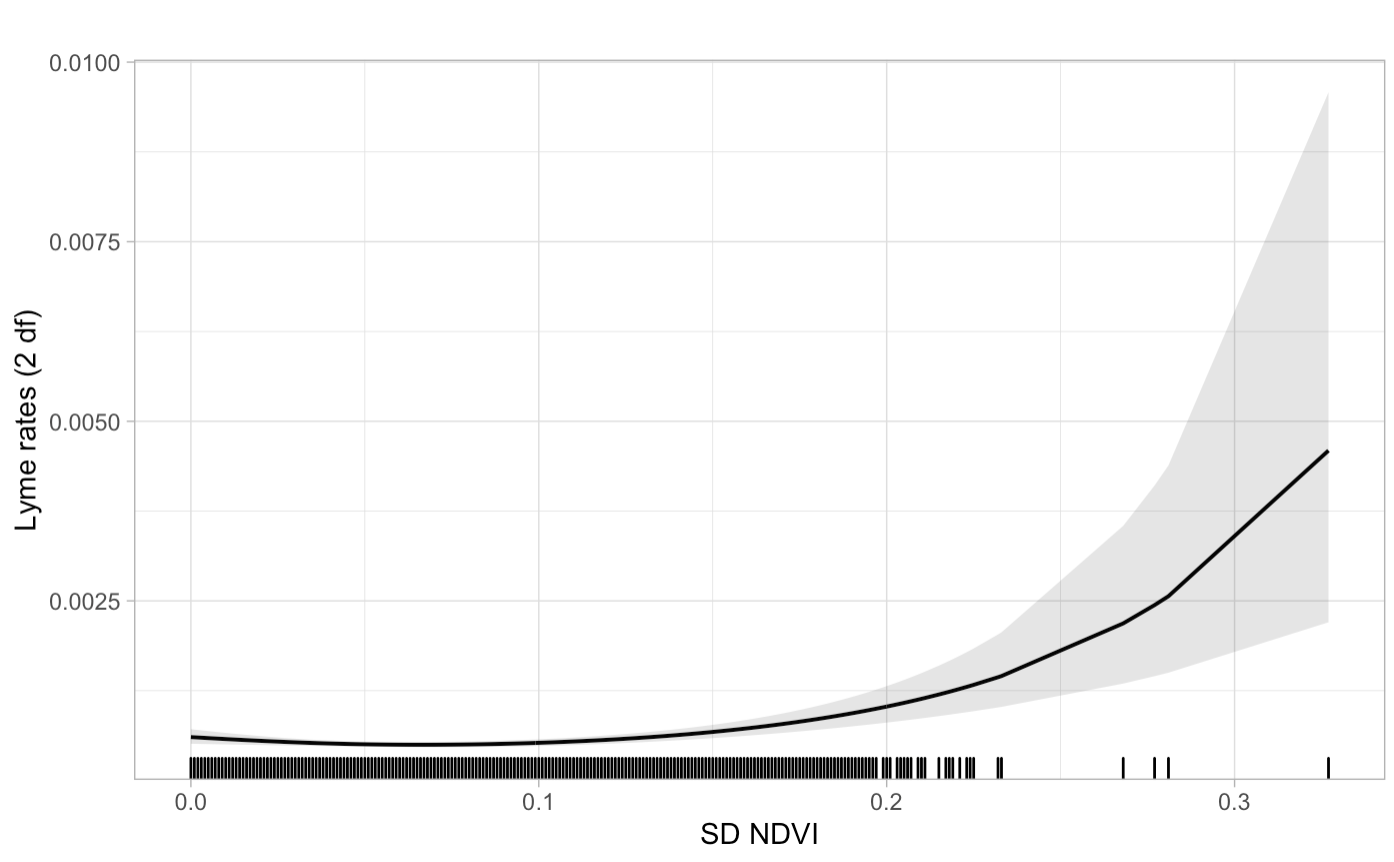

Figure S4D. Population Density

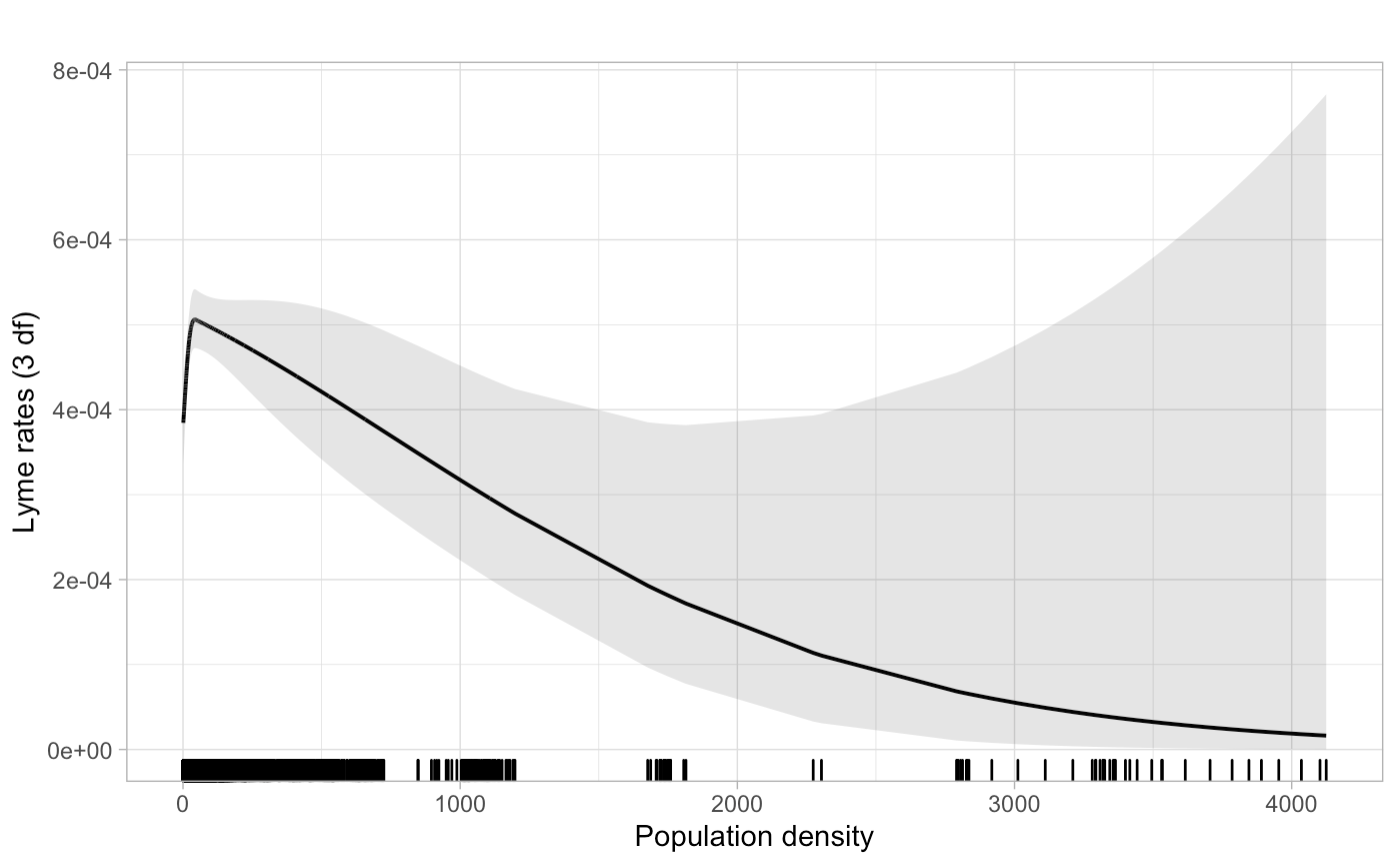

Figure S4E. Total Area of Land

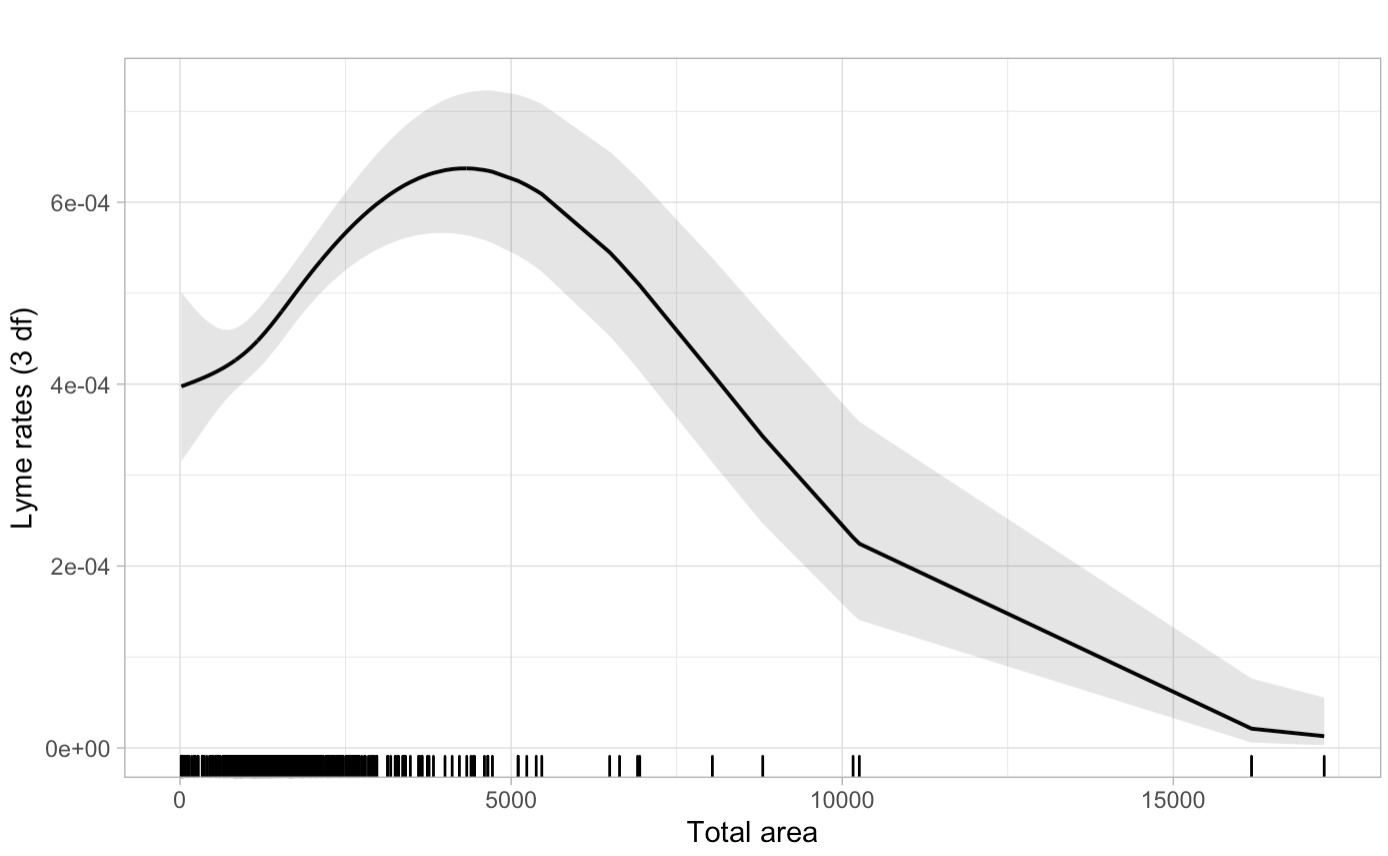

Figure S4F. Total area of water

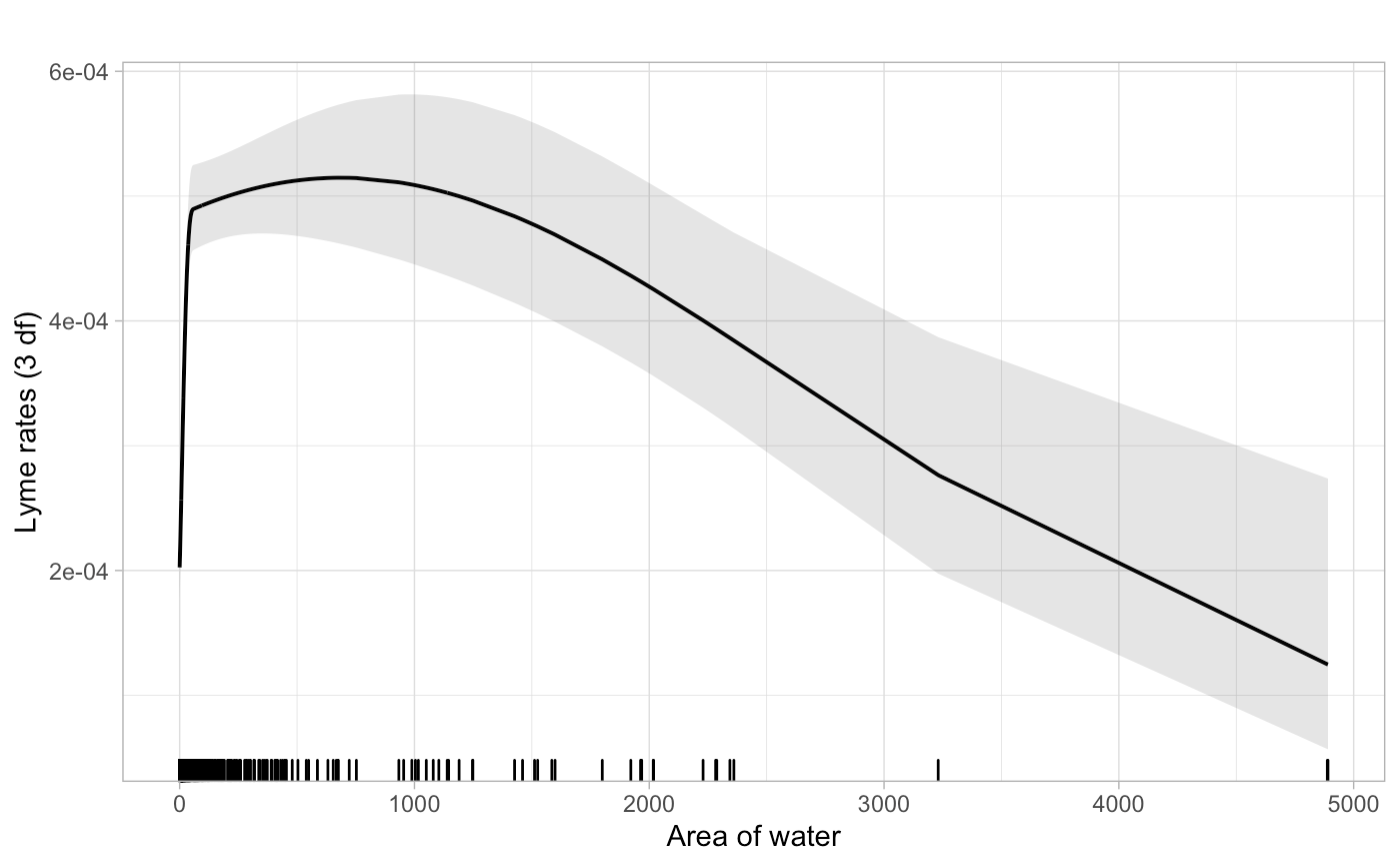

**Figure S5: State-specific analyses for average NDVI**

Figure S5A. Connecticut: Average NDVI

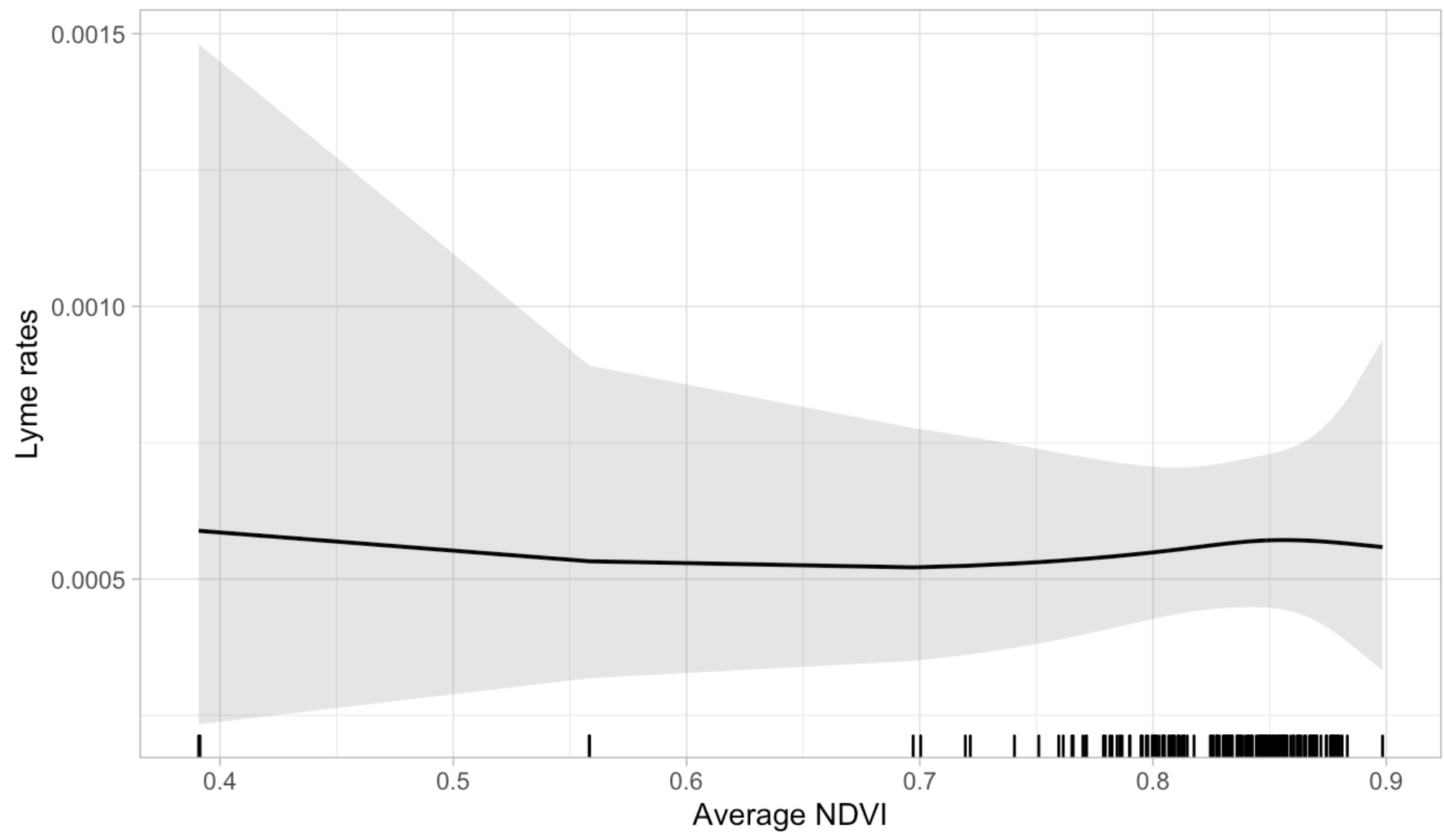

Figure S5B. Delaware: Average NDVI

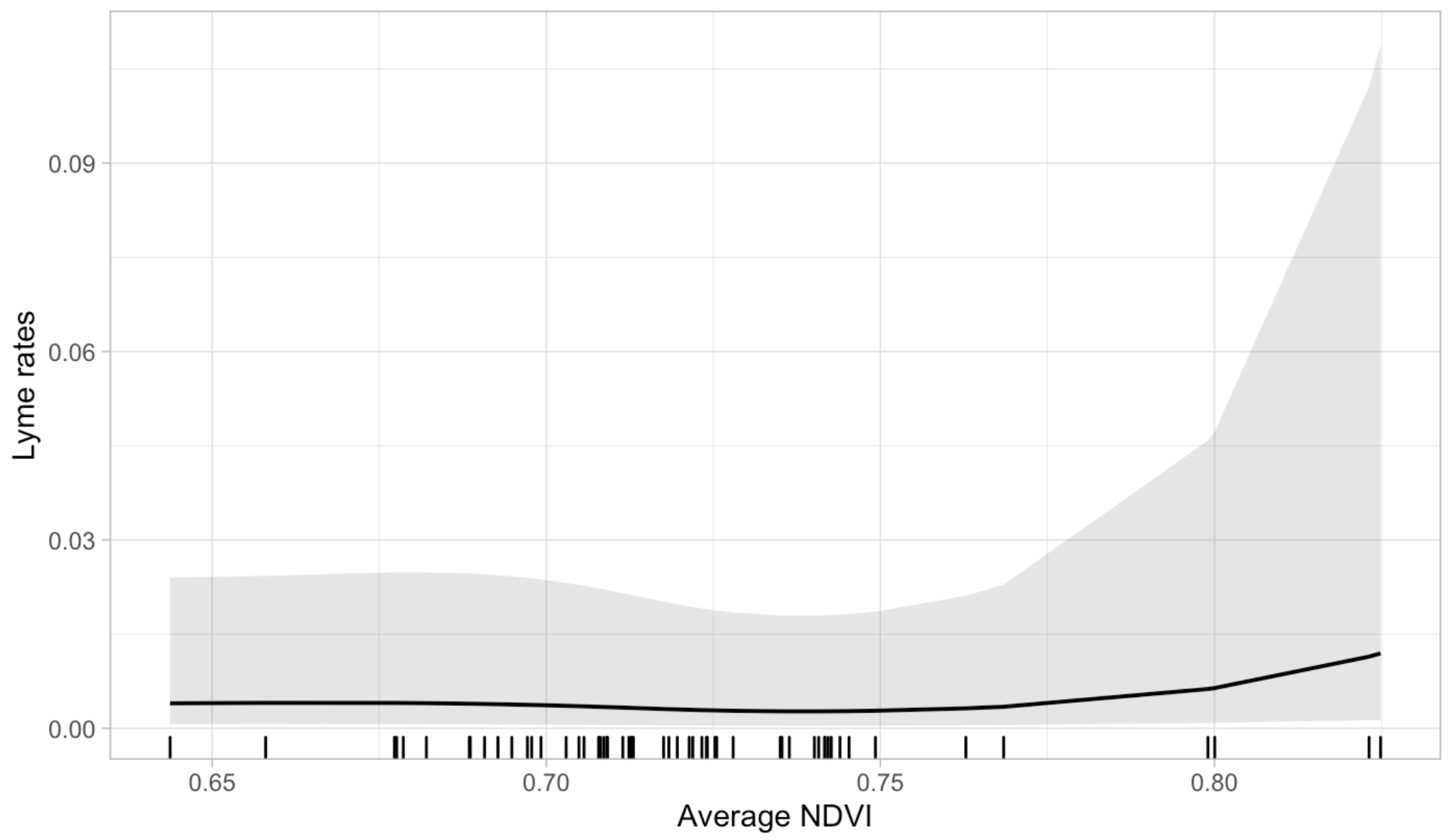

Figure S5C. District of Columbia: Average NDVI

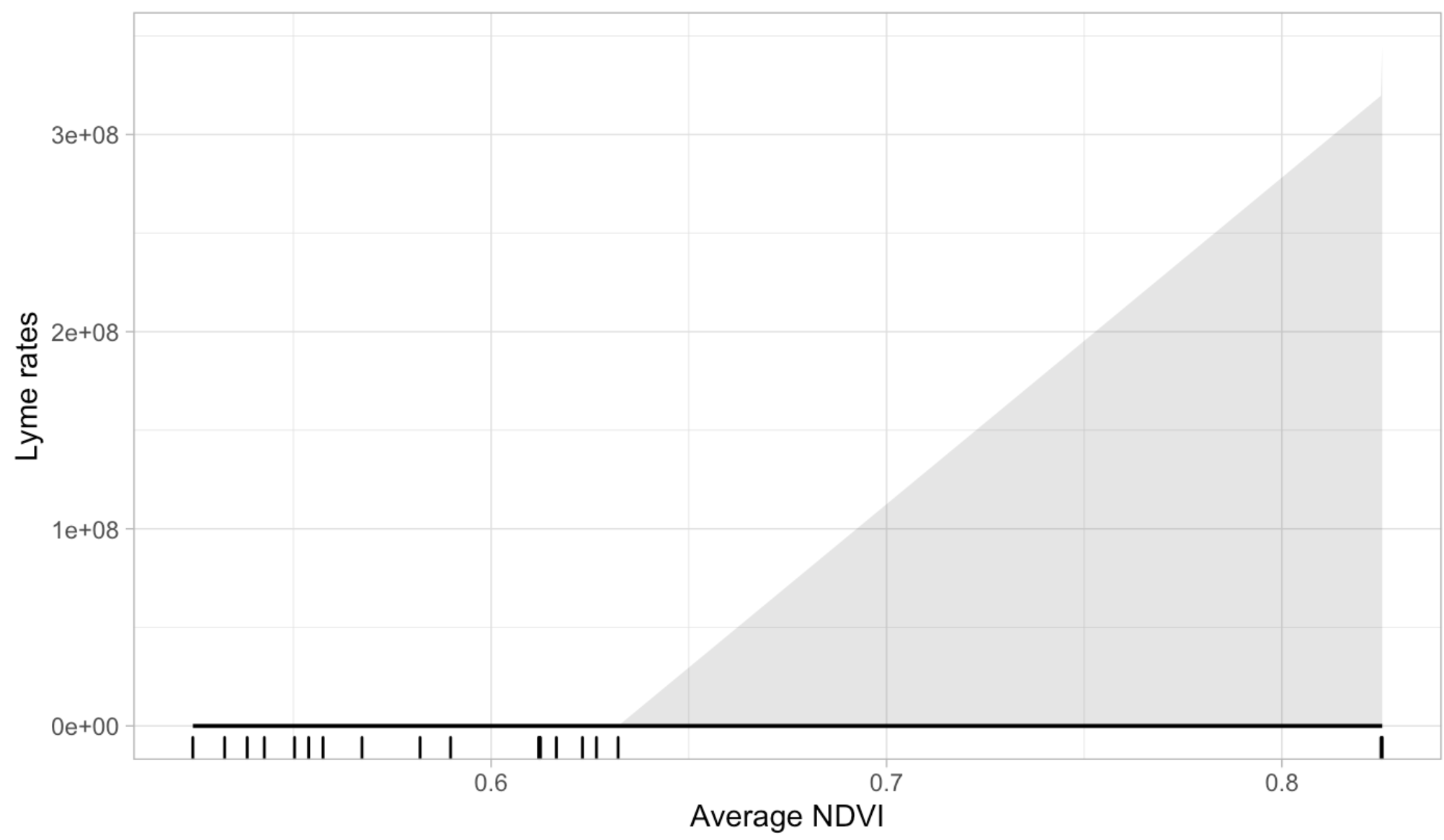

Figure S5D. Maine: Average NDVI

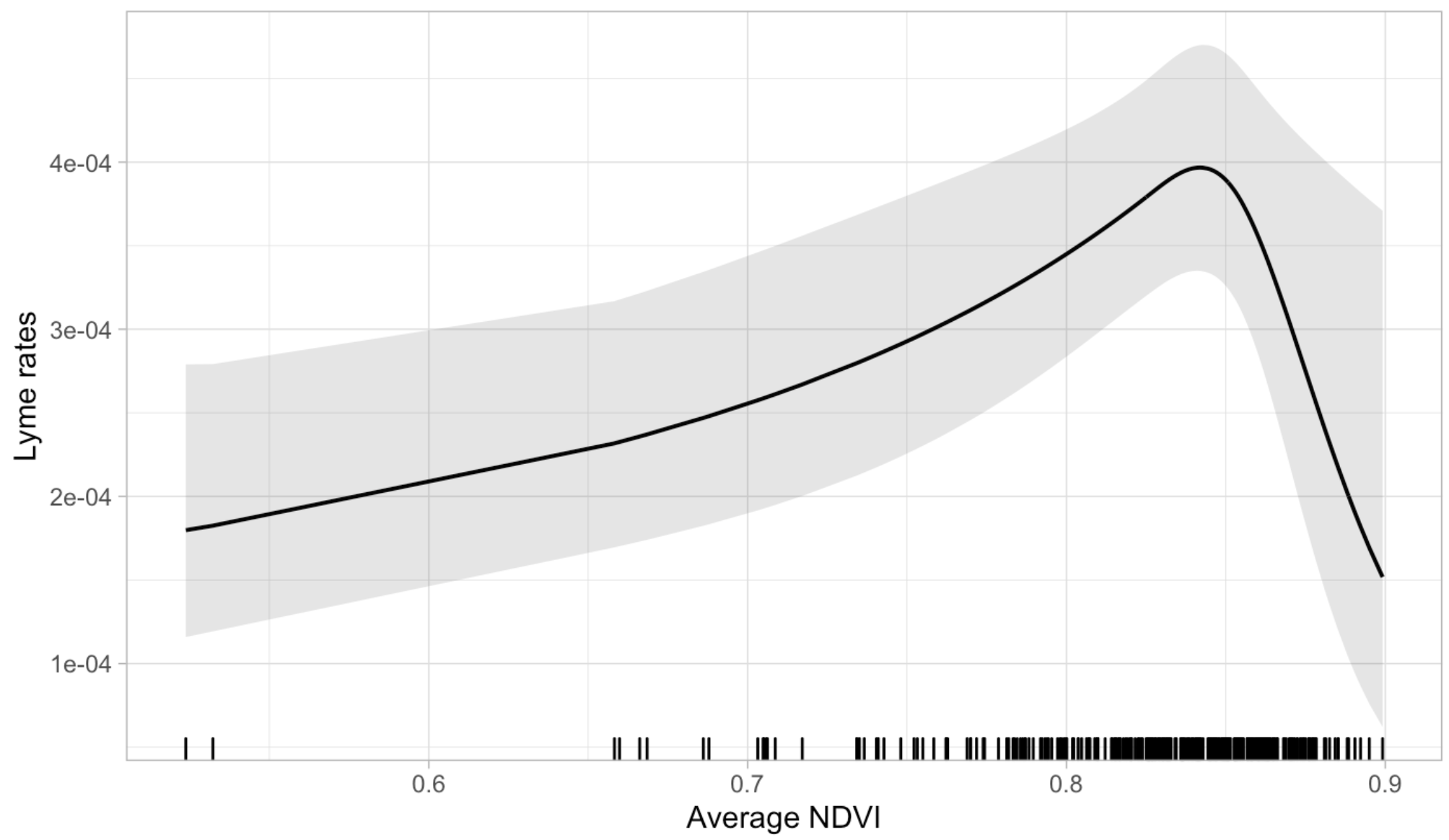

Figure S5E. Maryland: Average NDVI

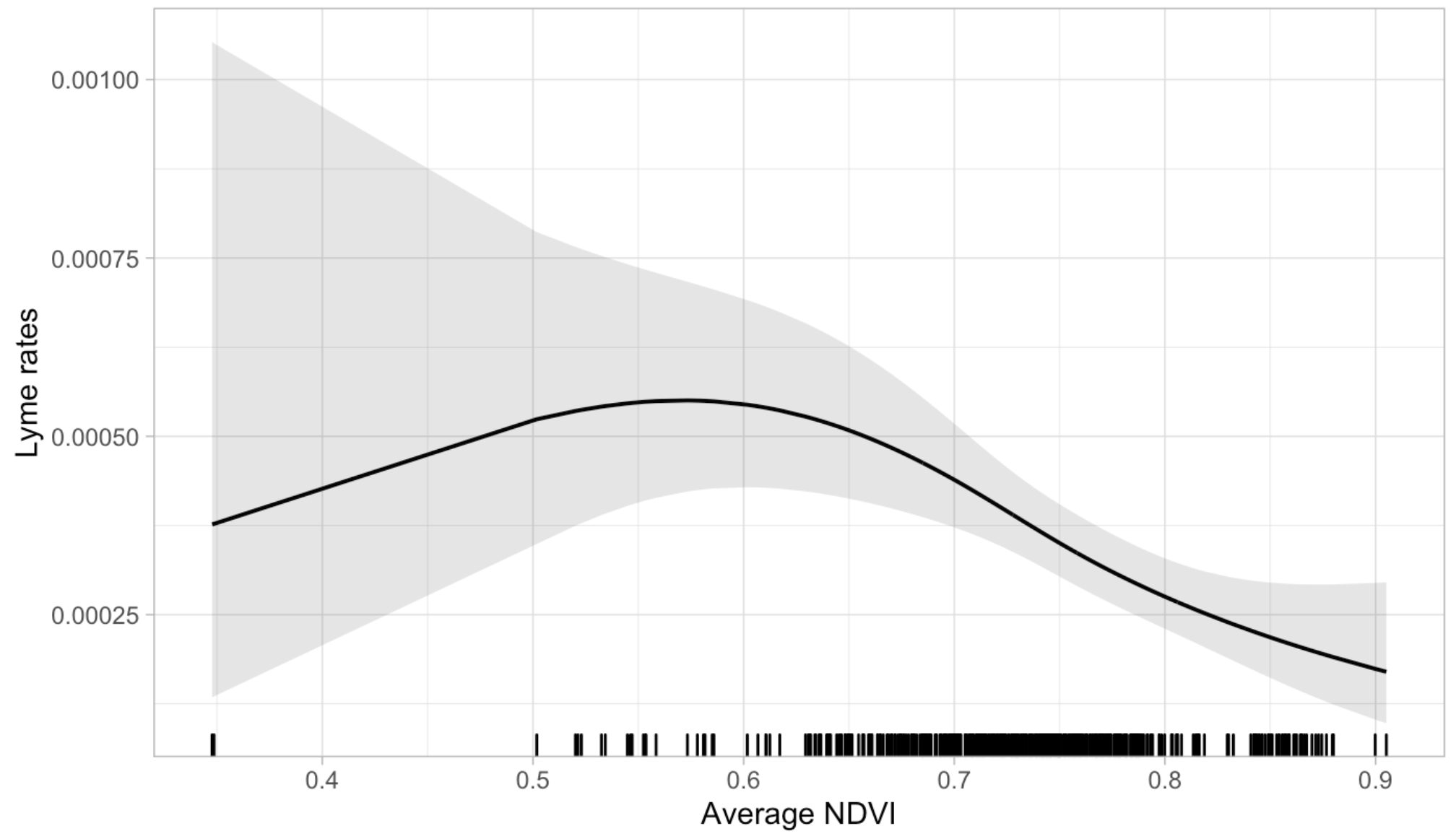

Figure S5F. Massachusetts: Average NDVI

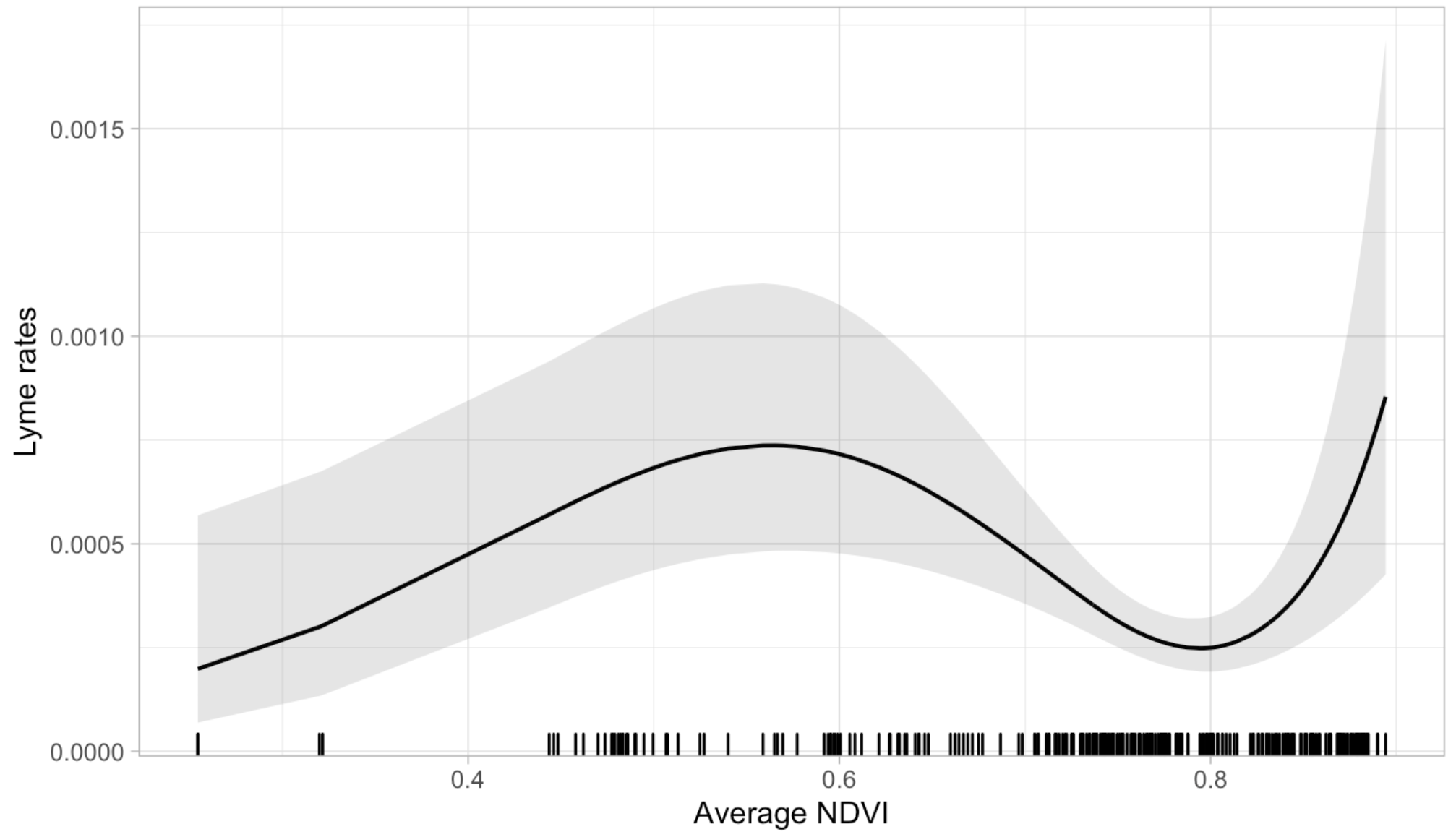

Figure S5G. Minnesota: Average NDVI

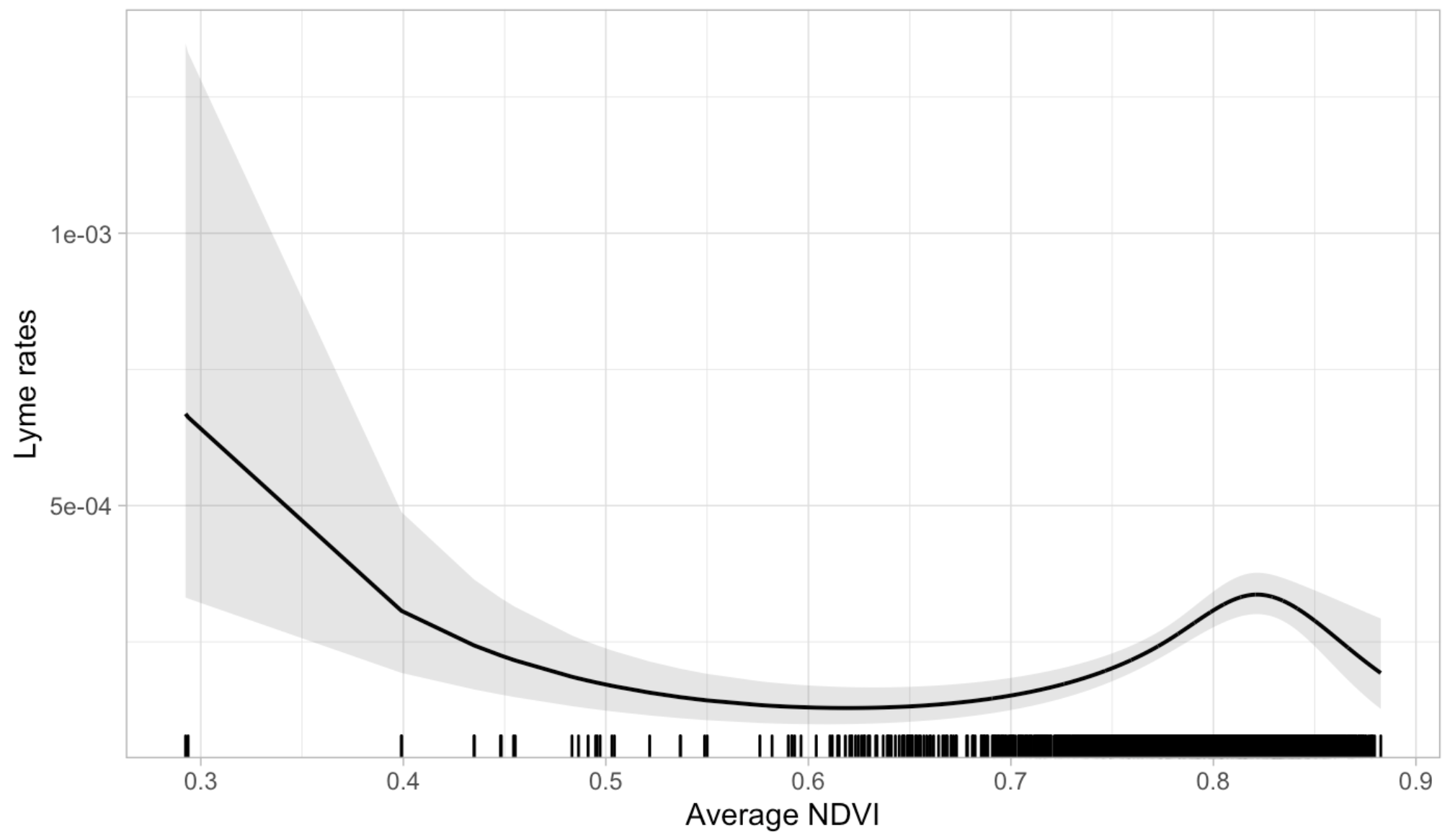

Figure S5H. New Hampshire: Average NDVI

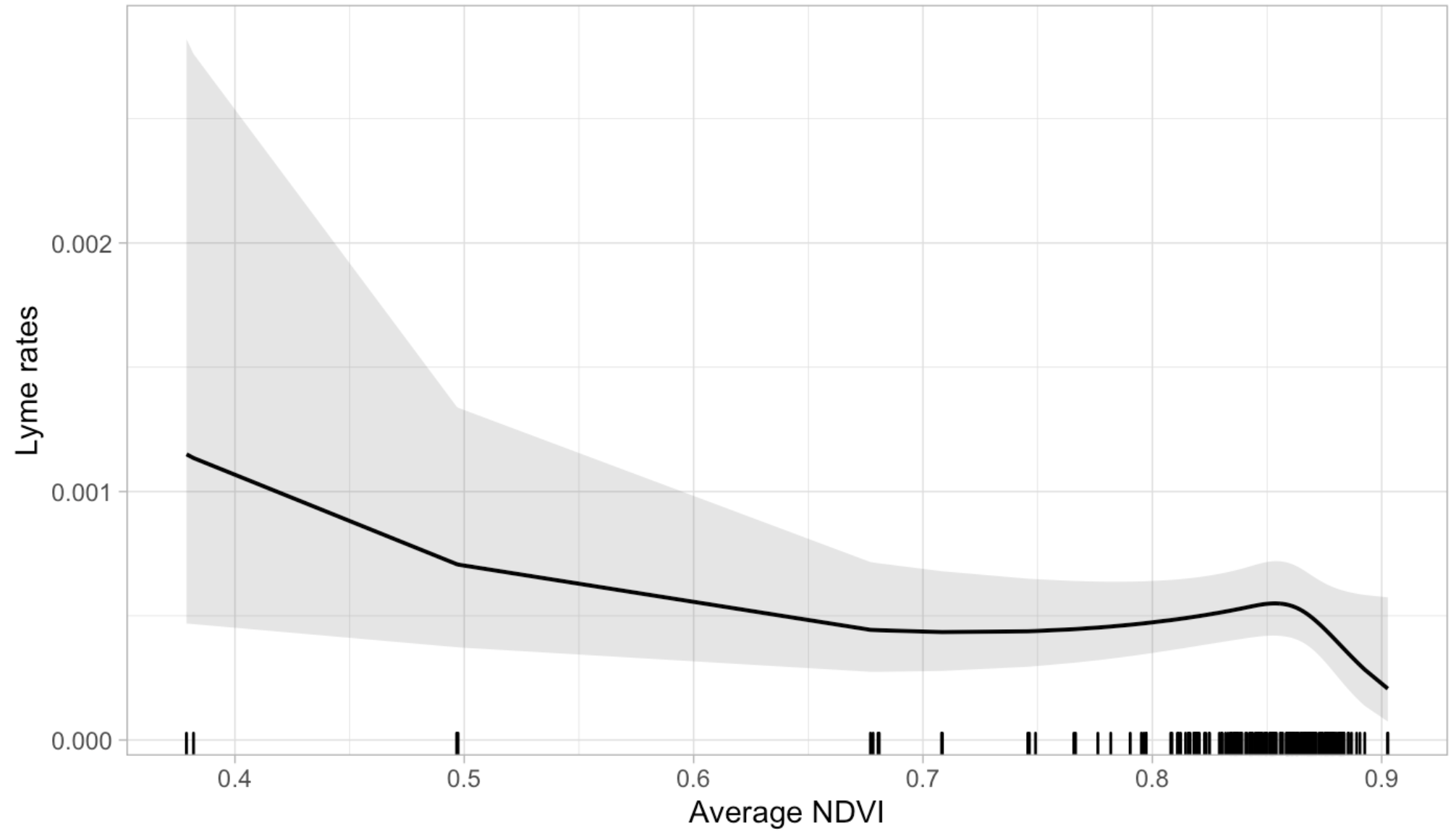

Figure S5I. New Jersey: Average NDVI

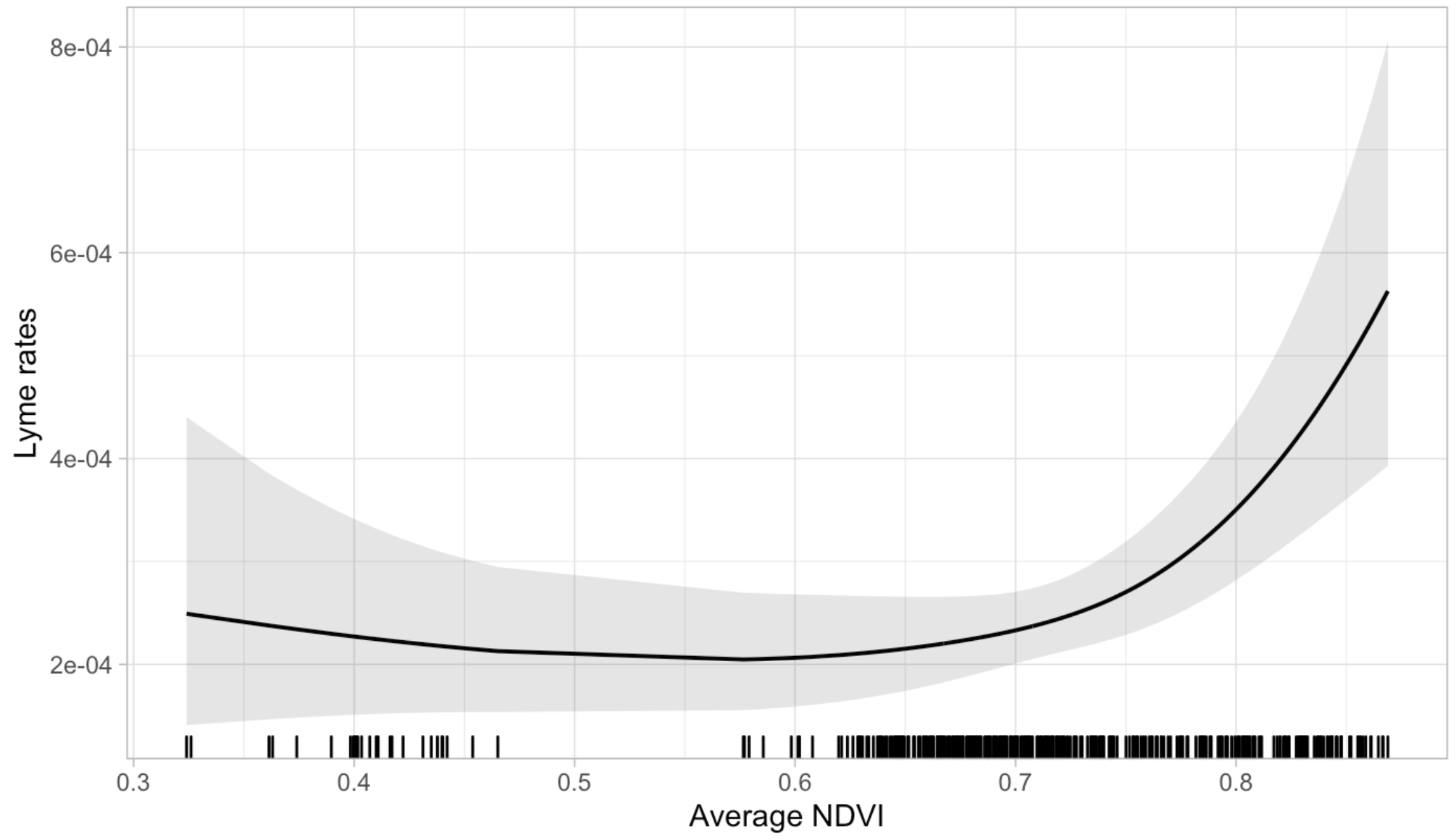

Figure S5J. New York: Average NDVI

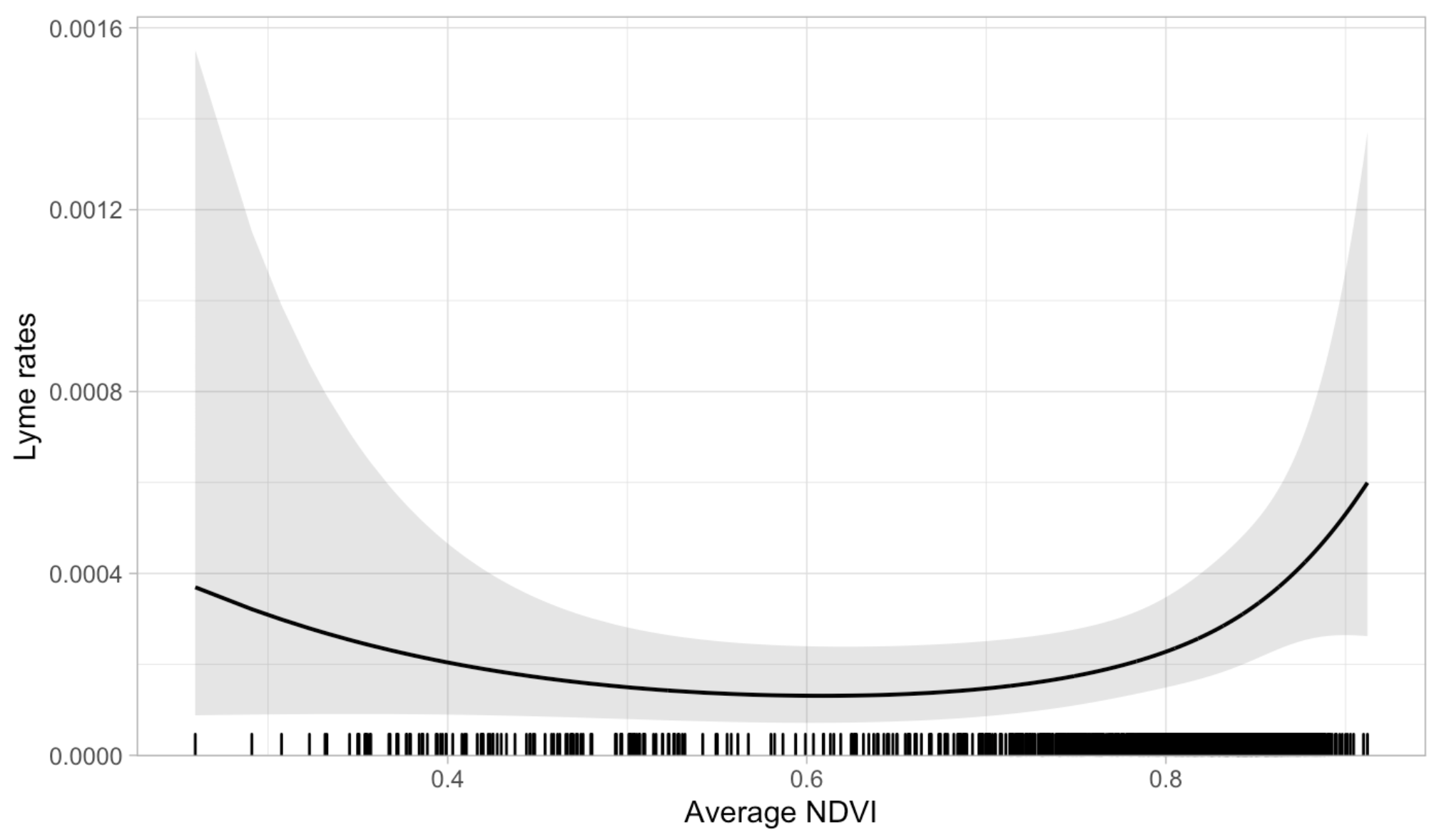

Figure S5K. Pennsylvania: Average NDVI

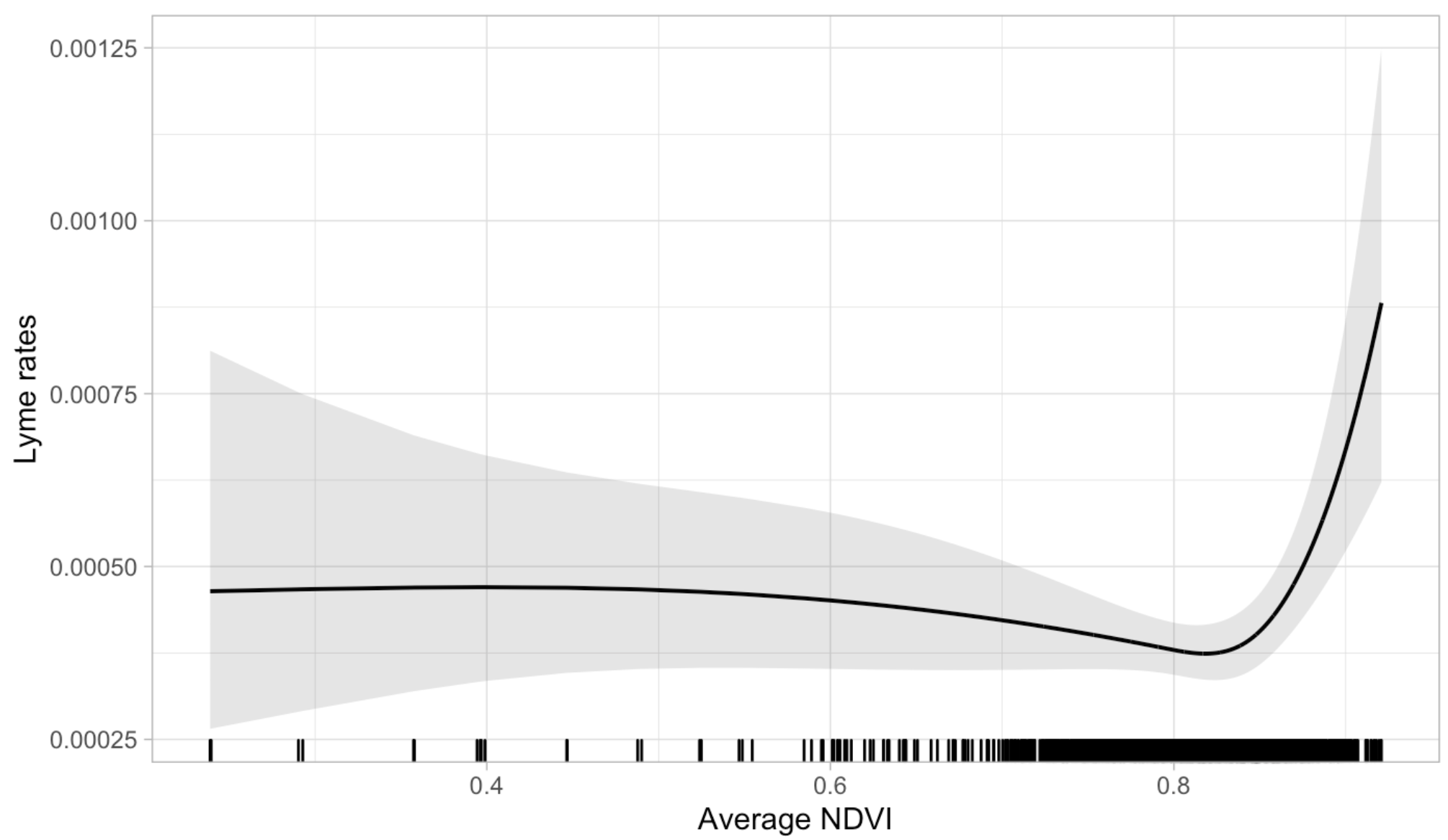

Figure S5L. Vermont: Average NDVI

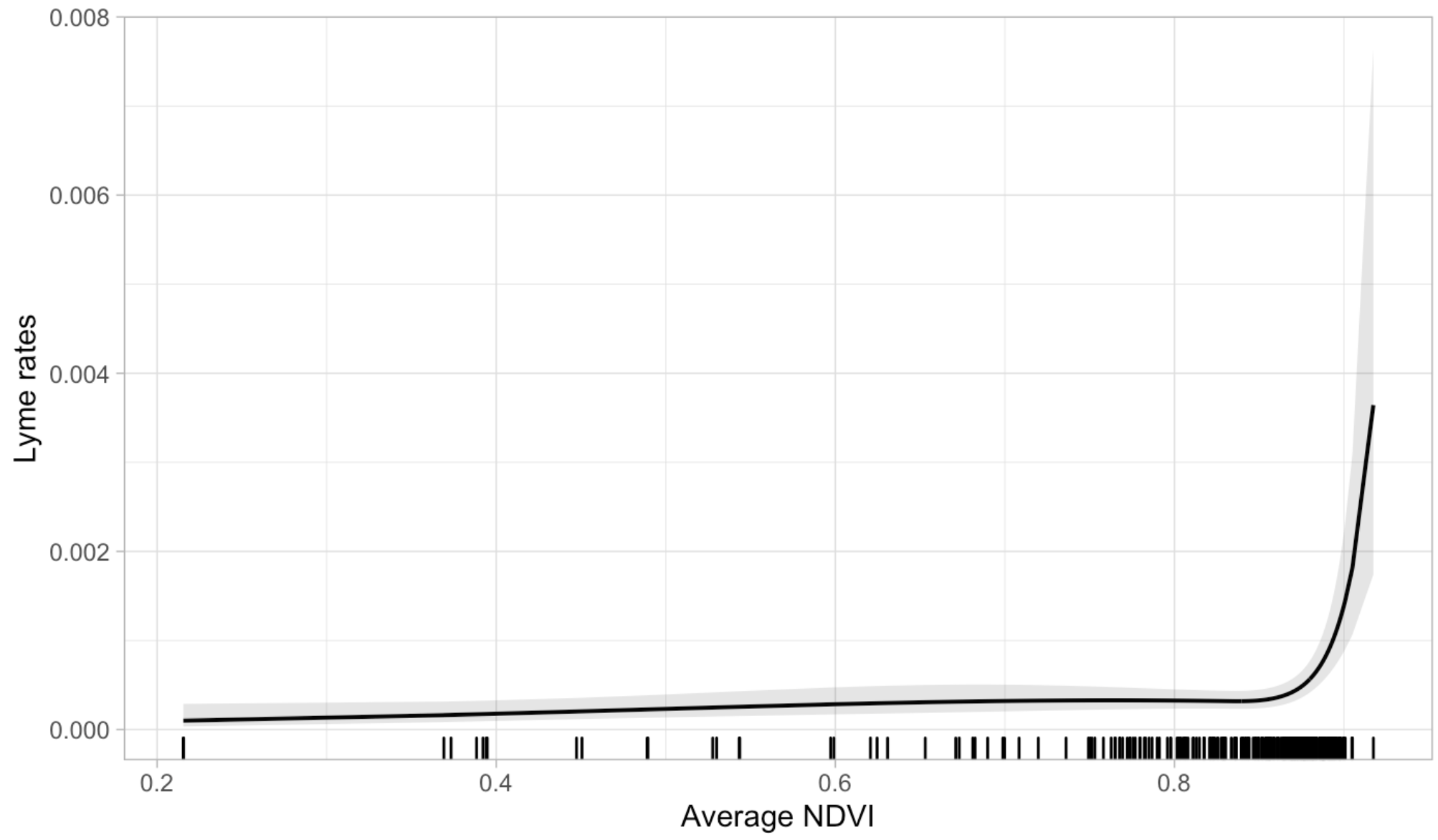

Figure S5M. Virginia: Average NDVI

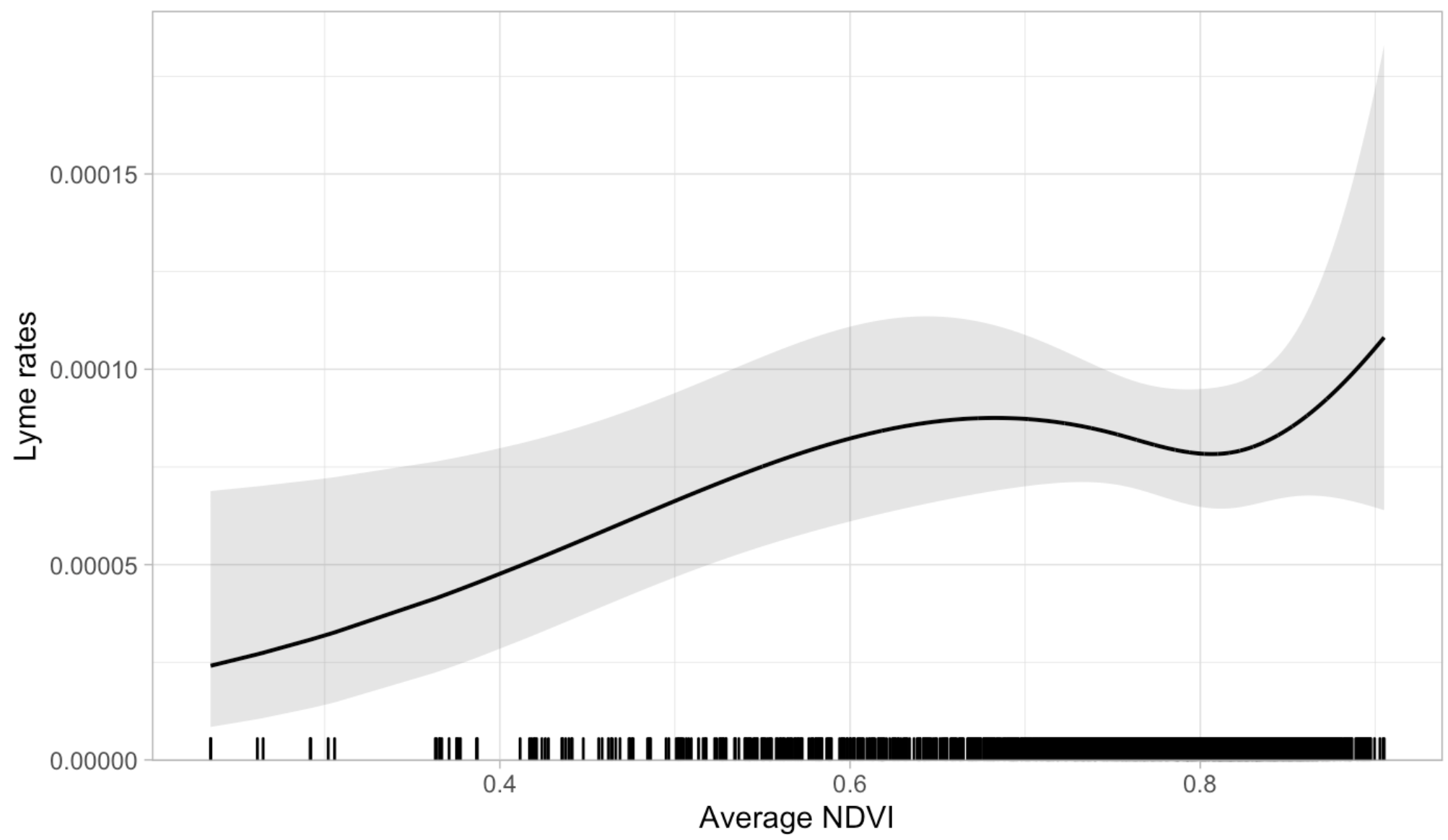

Figure S5N. West Virginia: Average NDVI
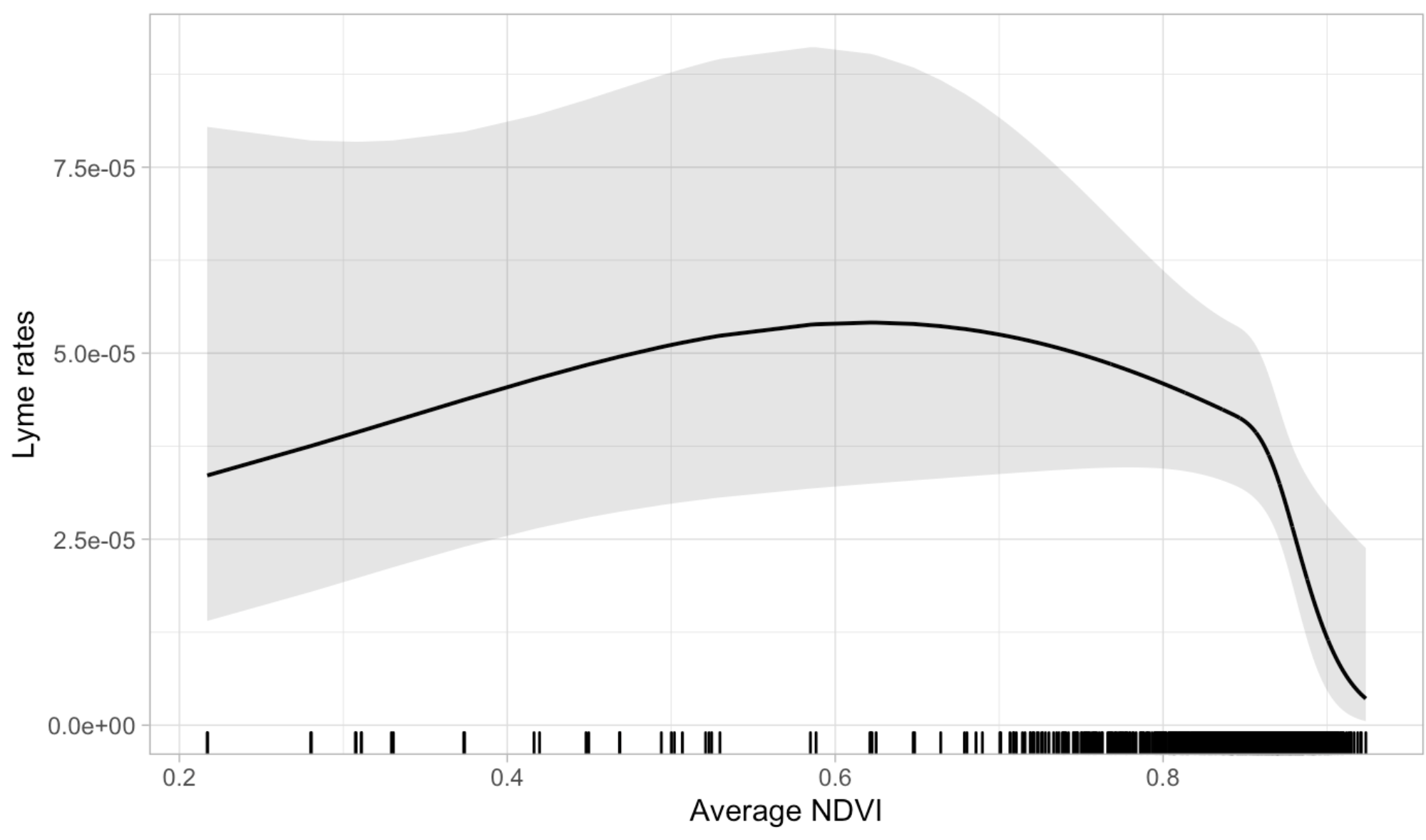

Figure S5O. Wisconsin: Average NDVI

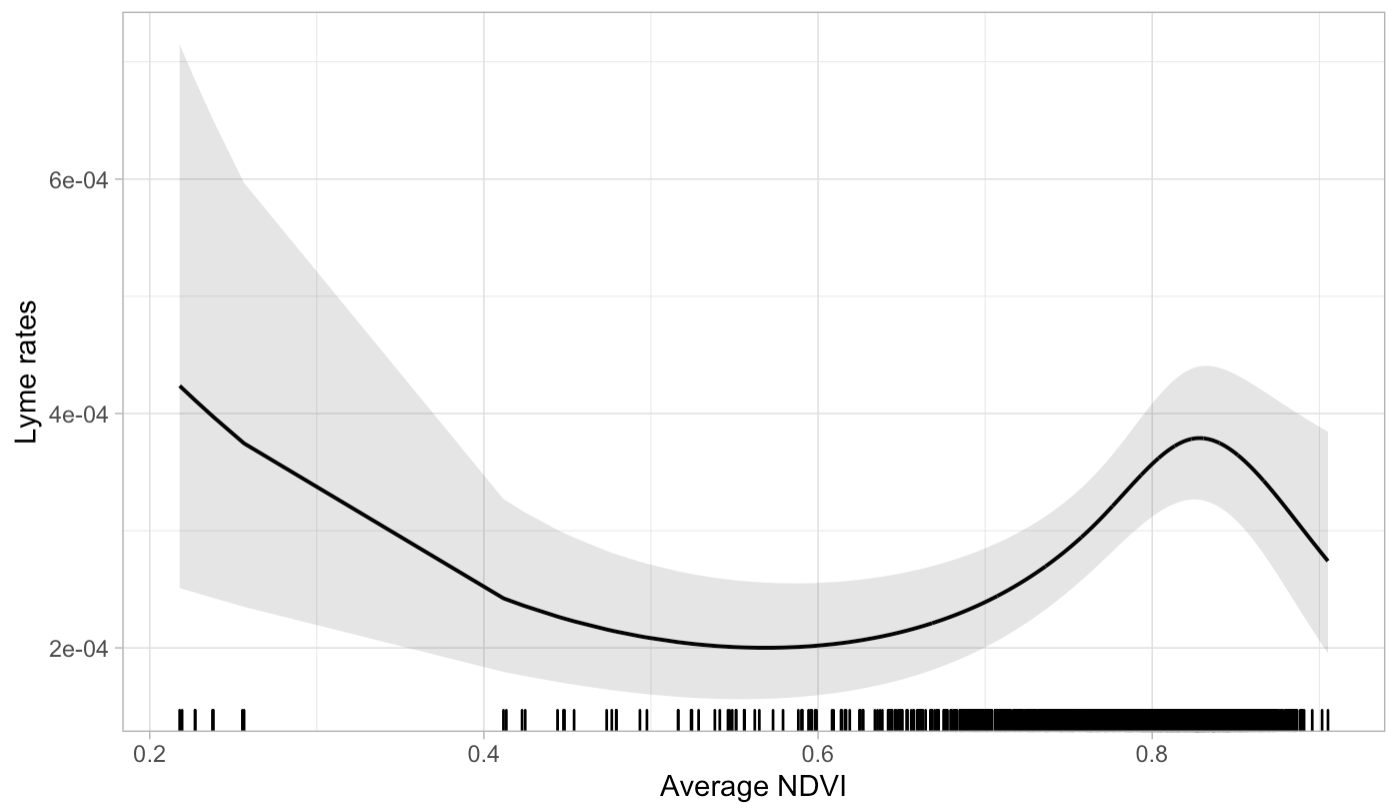

Figure S6A. Connecticut: Standard Deviation NDVI

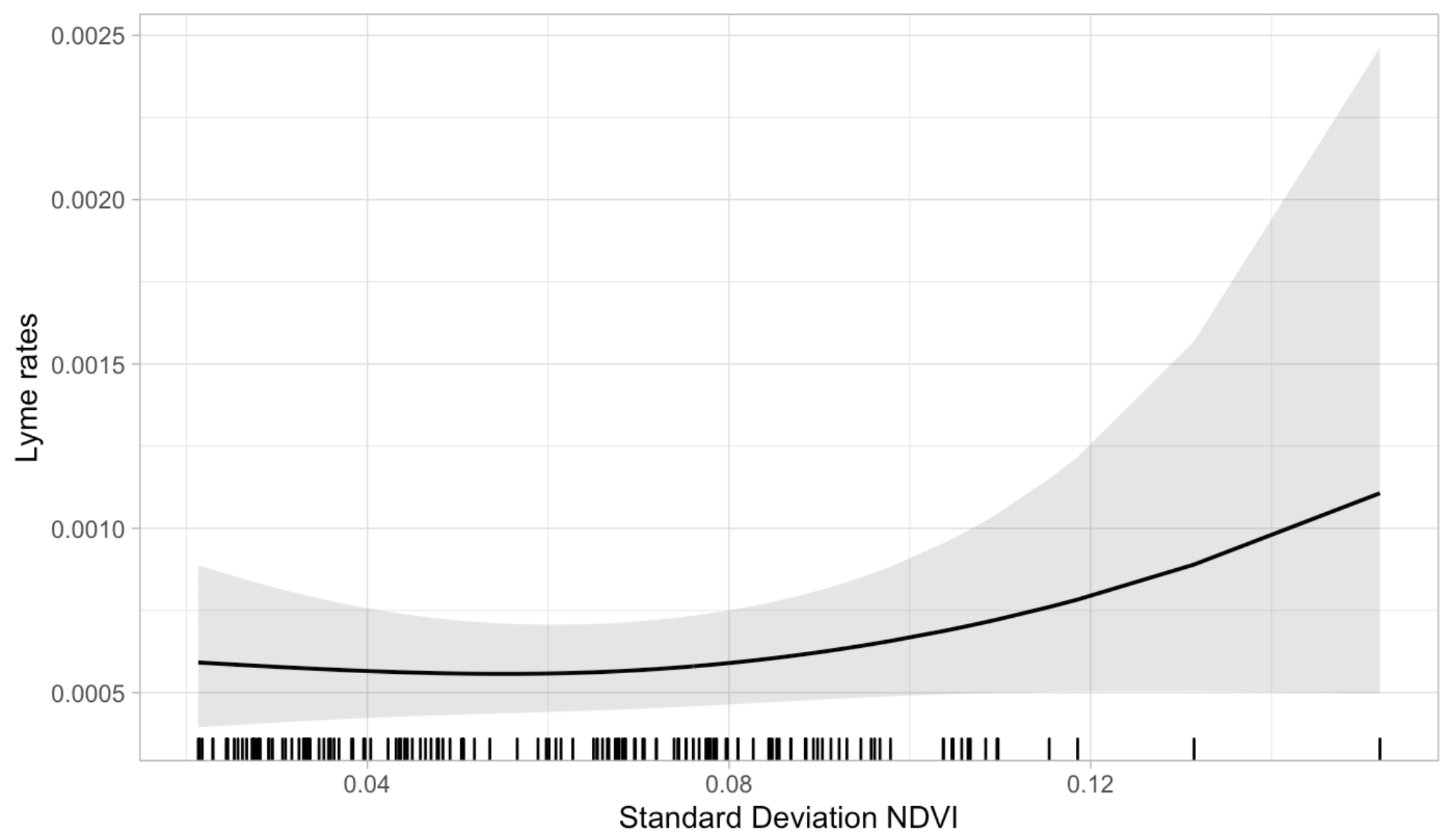

Figure S6B. Delaware: Standard Deviation NDVI

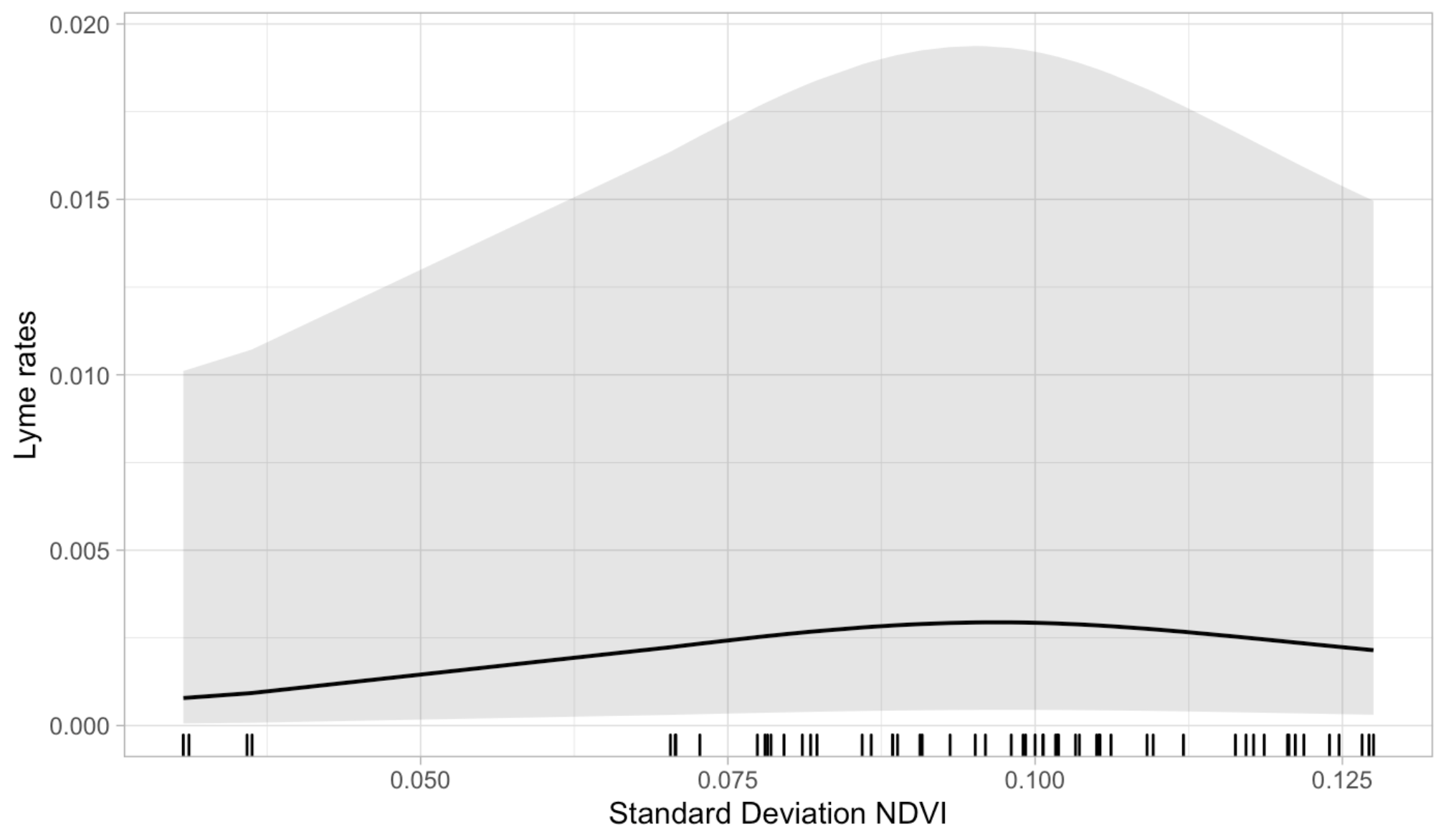

Figure S6C. District of Columbia: Standard Deviation NDVI

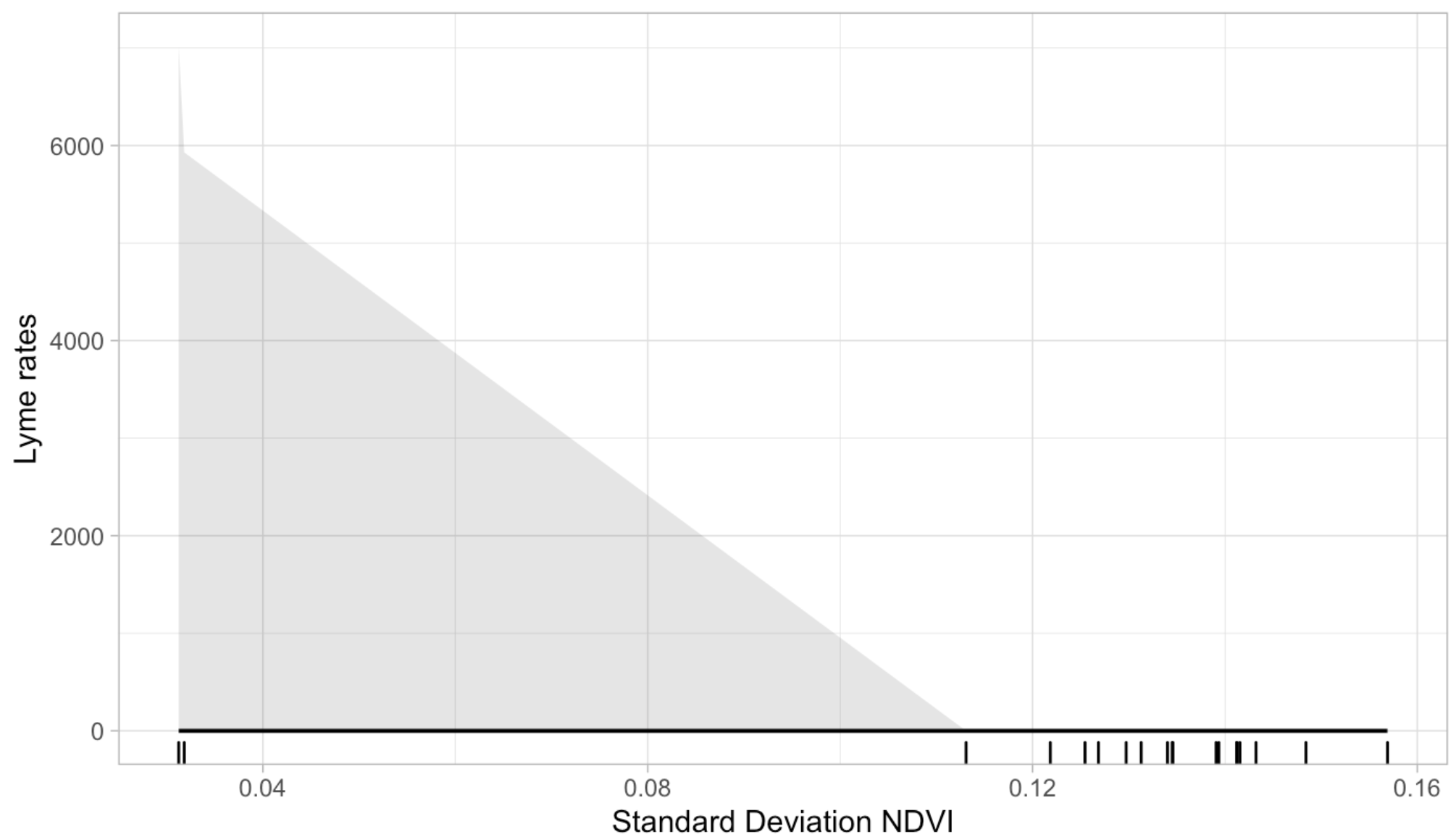

Figure S6D. Maine: Standard Deviation NDVI

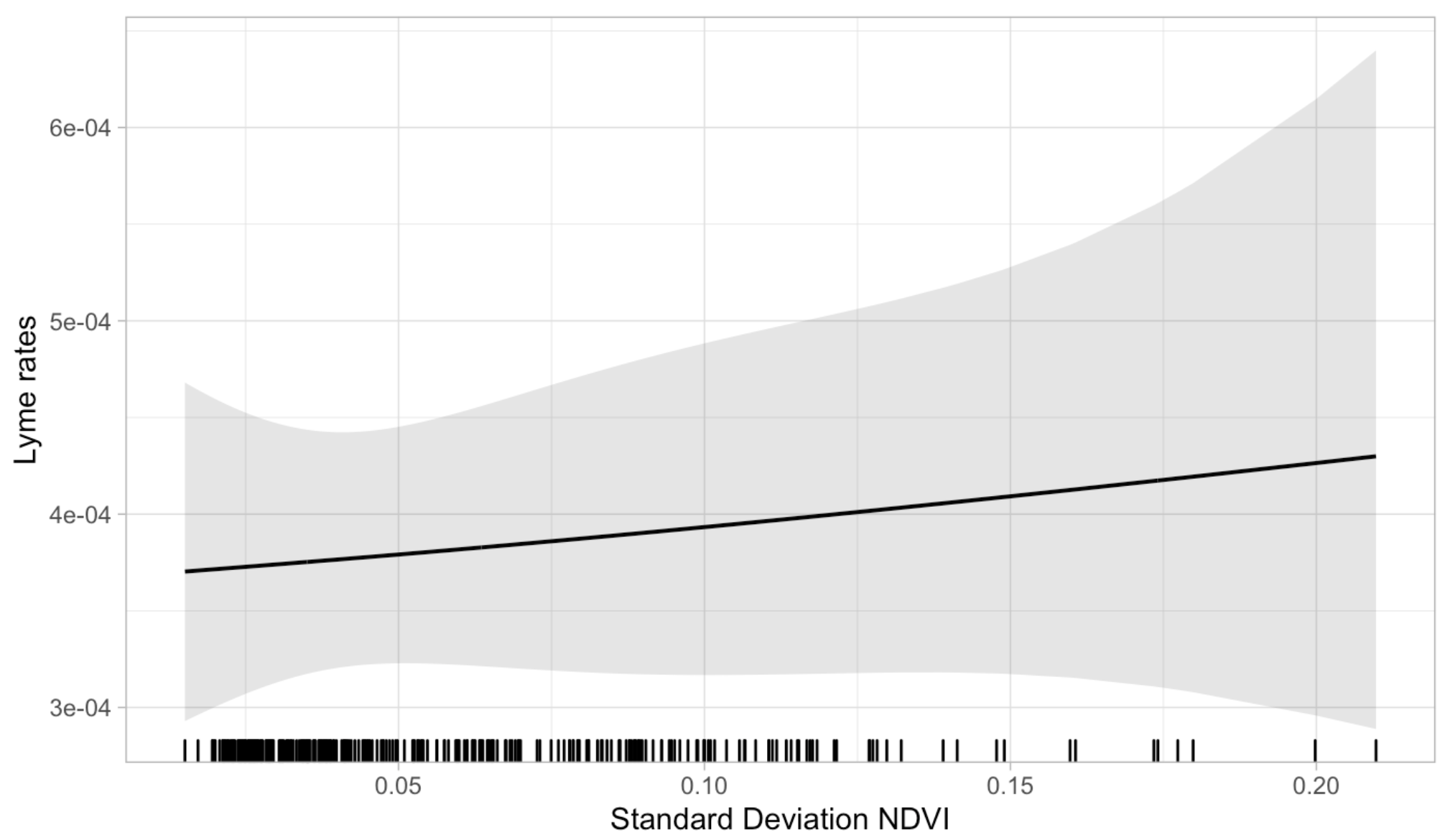

Figure S6E. Maryland: Standard Deviation NDVI

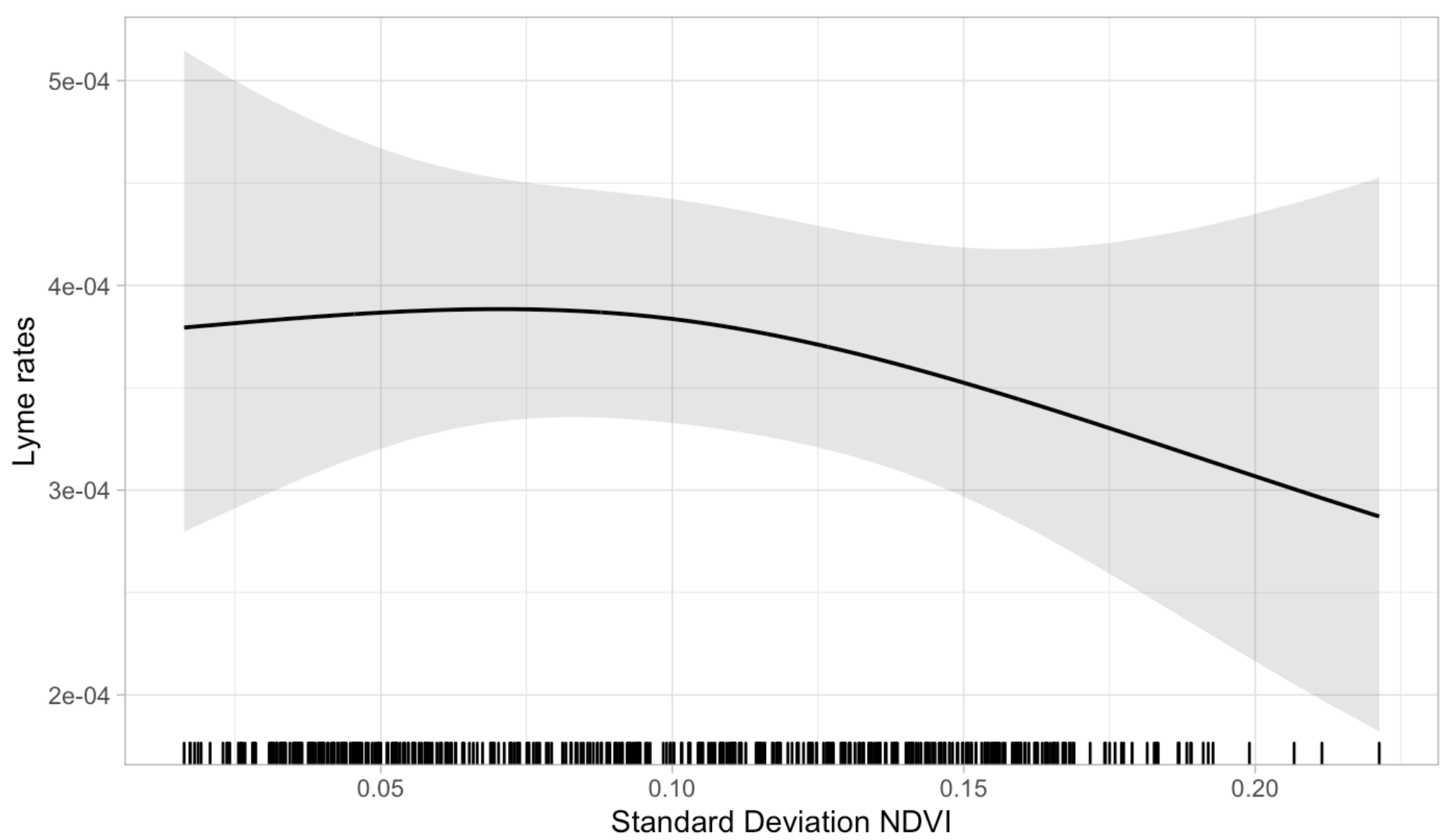

Figure S6F. Massachusetts: Standard Deviation NDVI

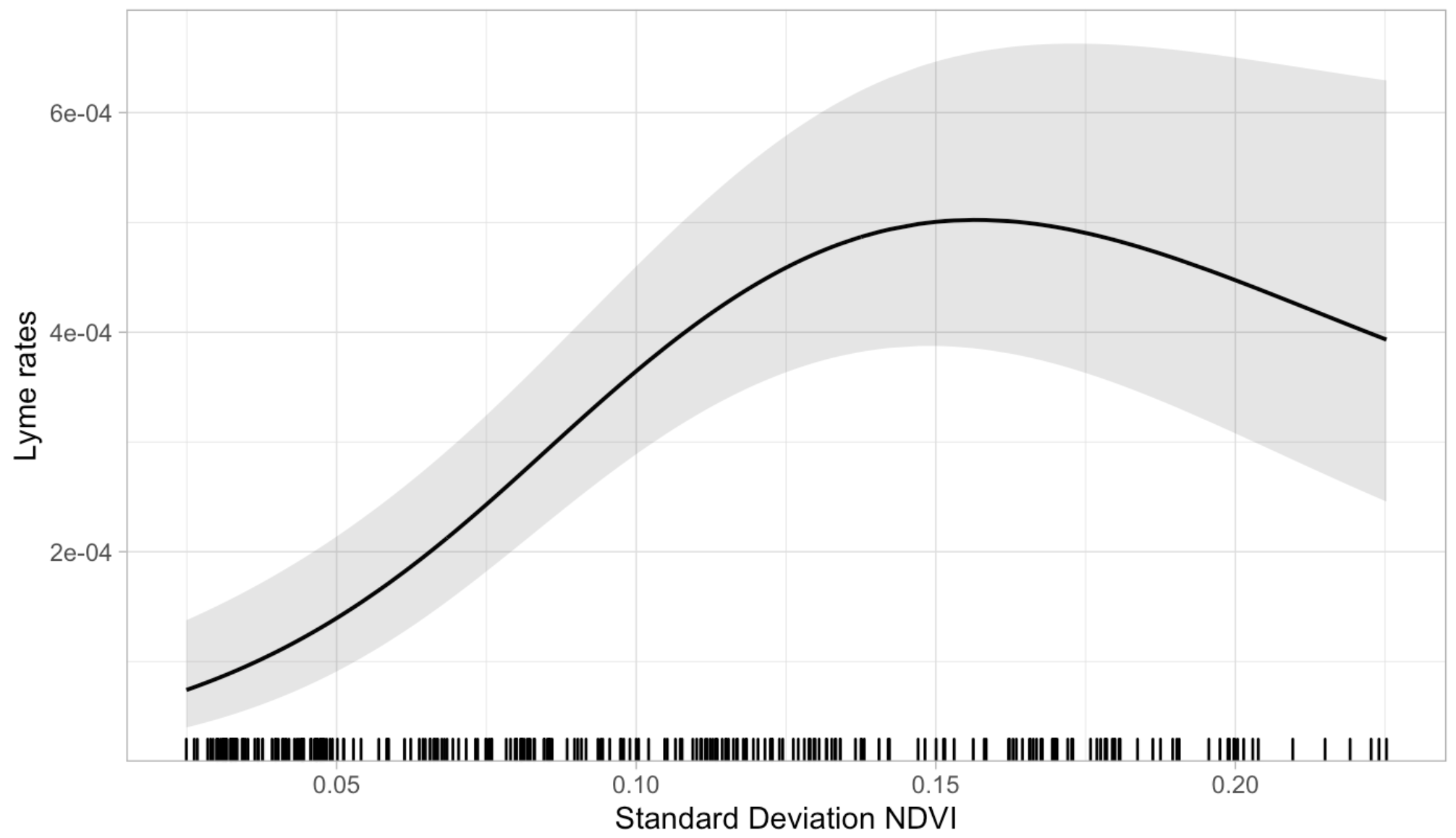

Figure S6G. Minnesota: Standard Deviation NDVI

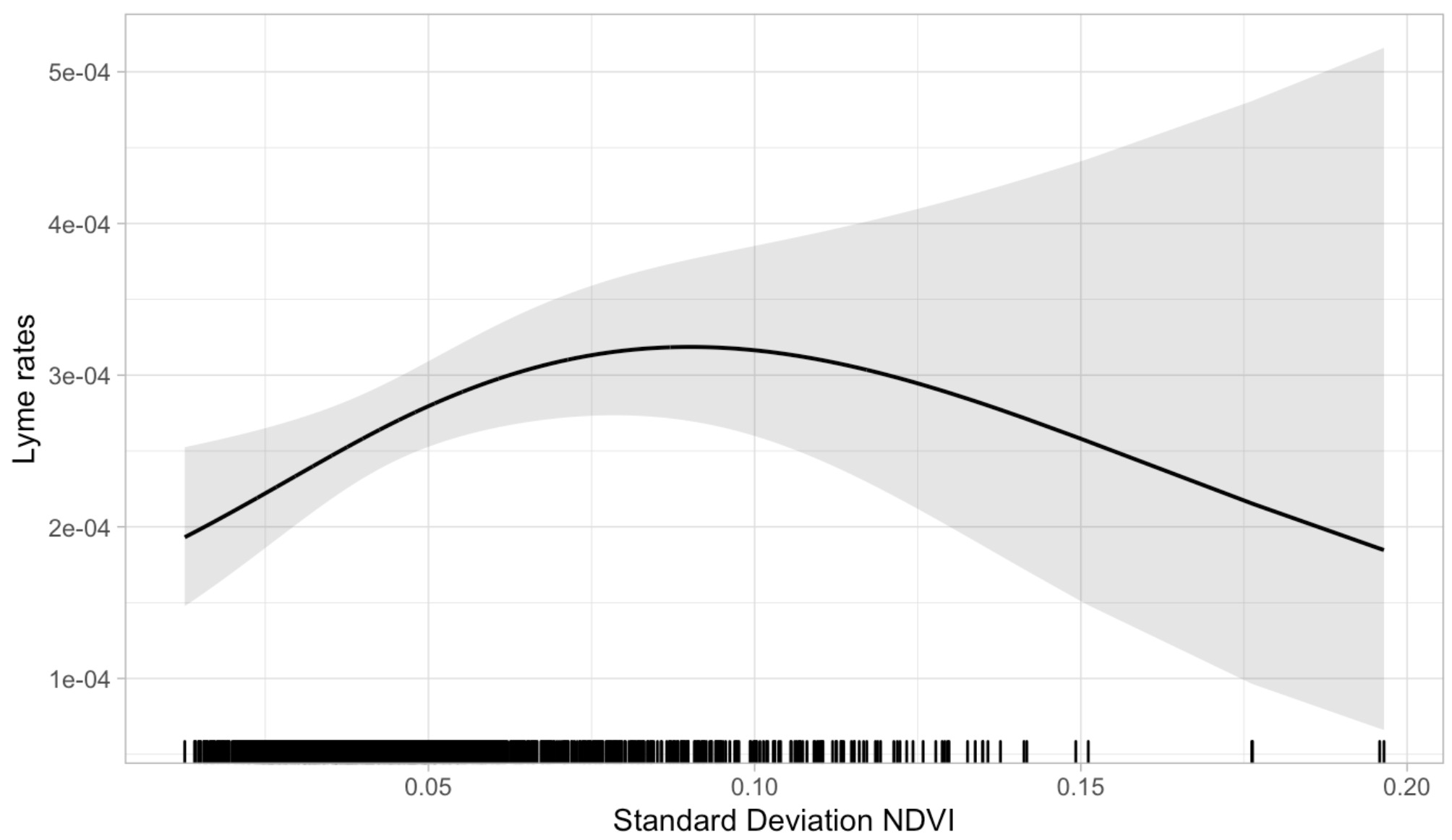

Figure S6H. New Hampshire: Standard Deviation NDVI

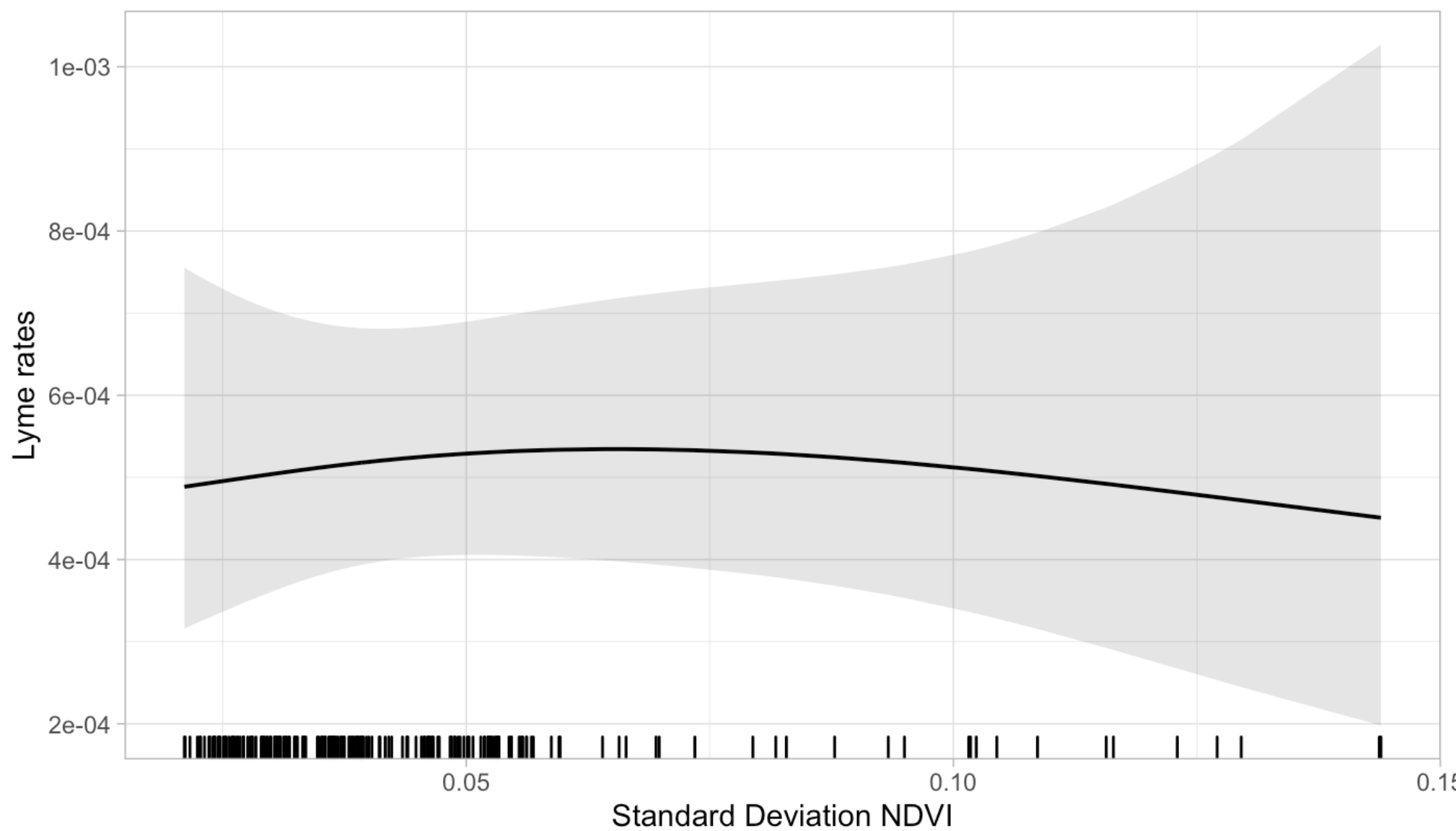

Figure S6I. New Jersey: Standard Deviation NDVI

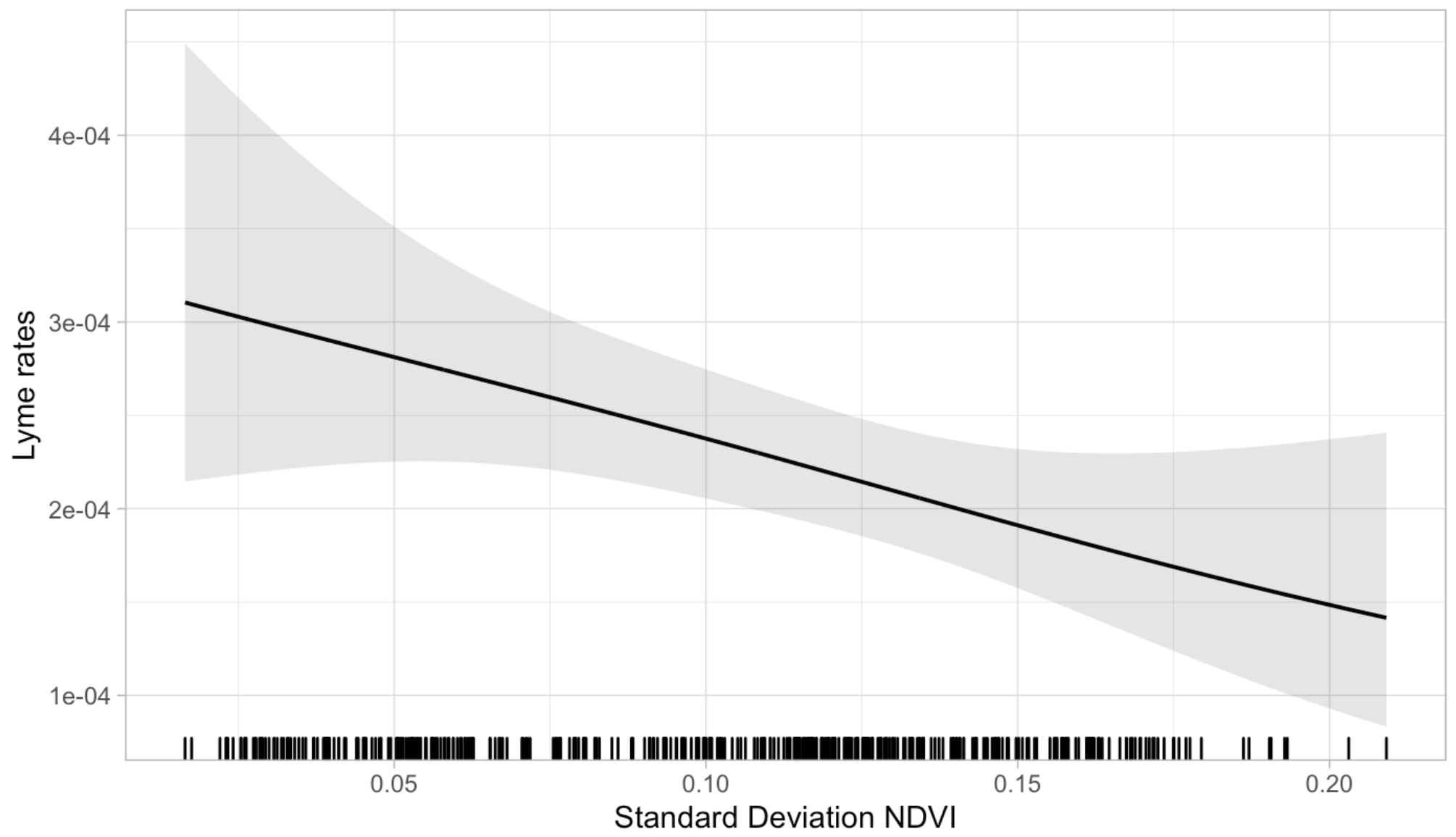

Figure S6J. New York: Standard Deviation NDVI

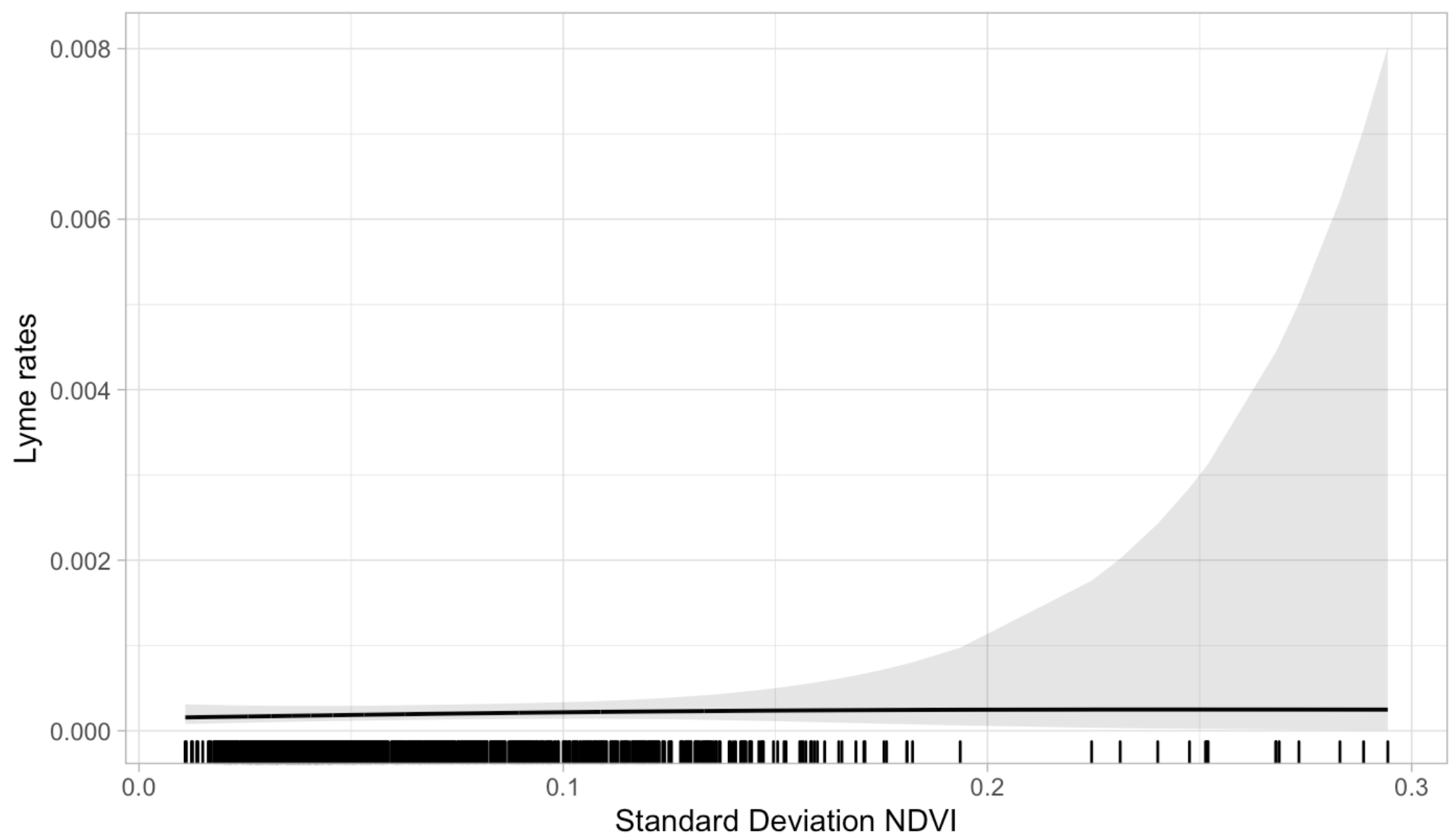

Figure S6K. Pennsylvania: Standard Deviation NDVI

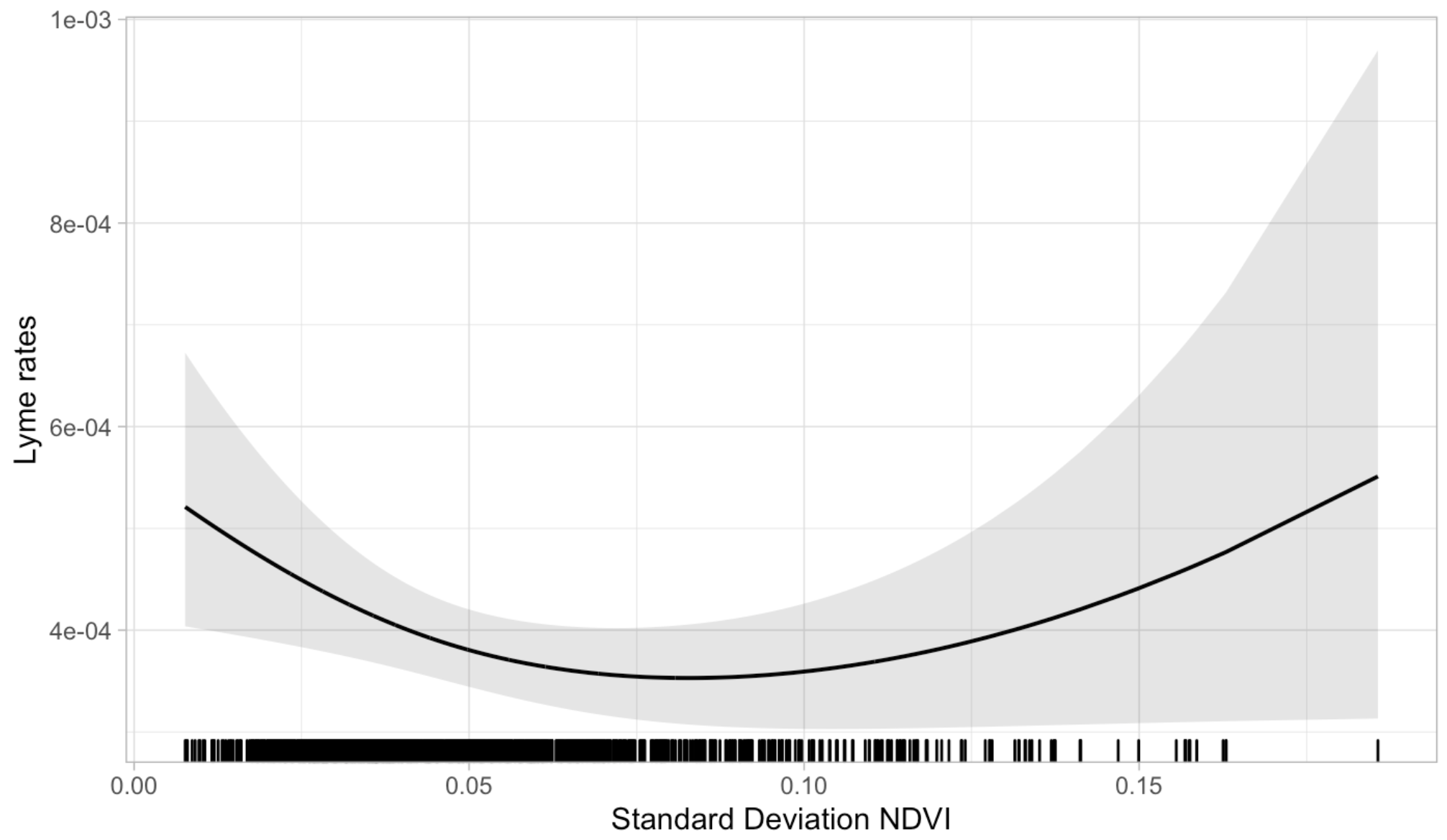

Figure S6L. Vermont: Standard Deviation NDVI

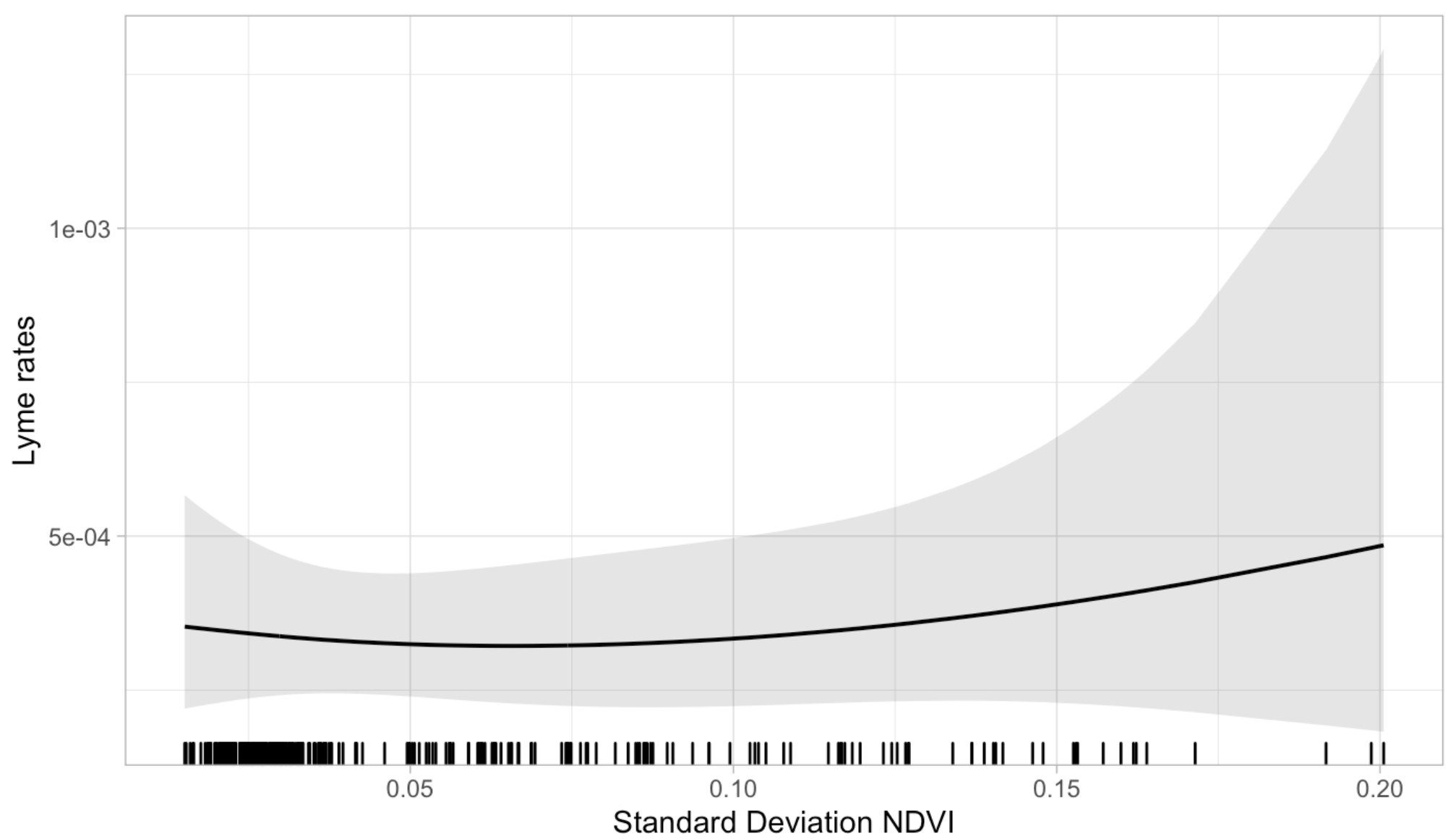

Figure S6M. Virginia: Standard Deviation NDVI

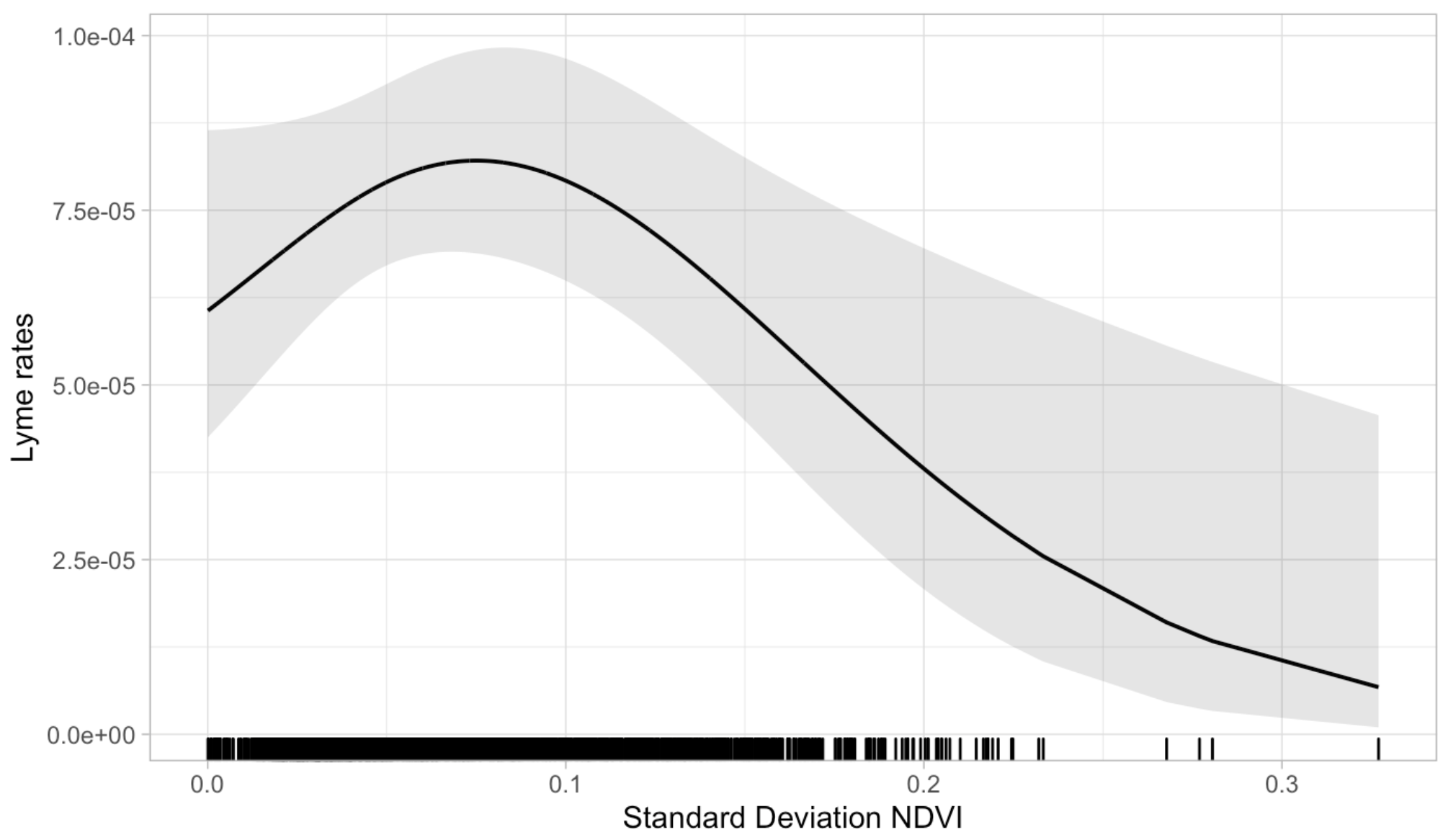

Figure S6N. West Virginia: Standard Deviation NDVI
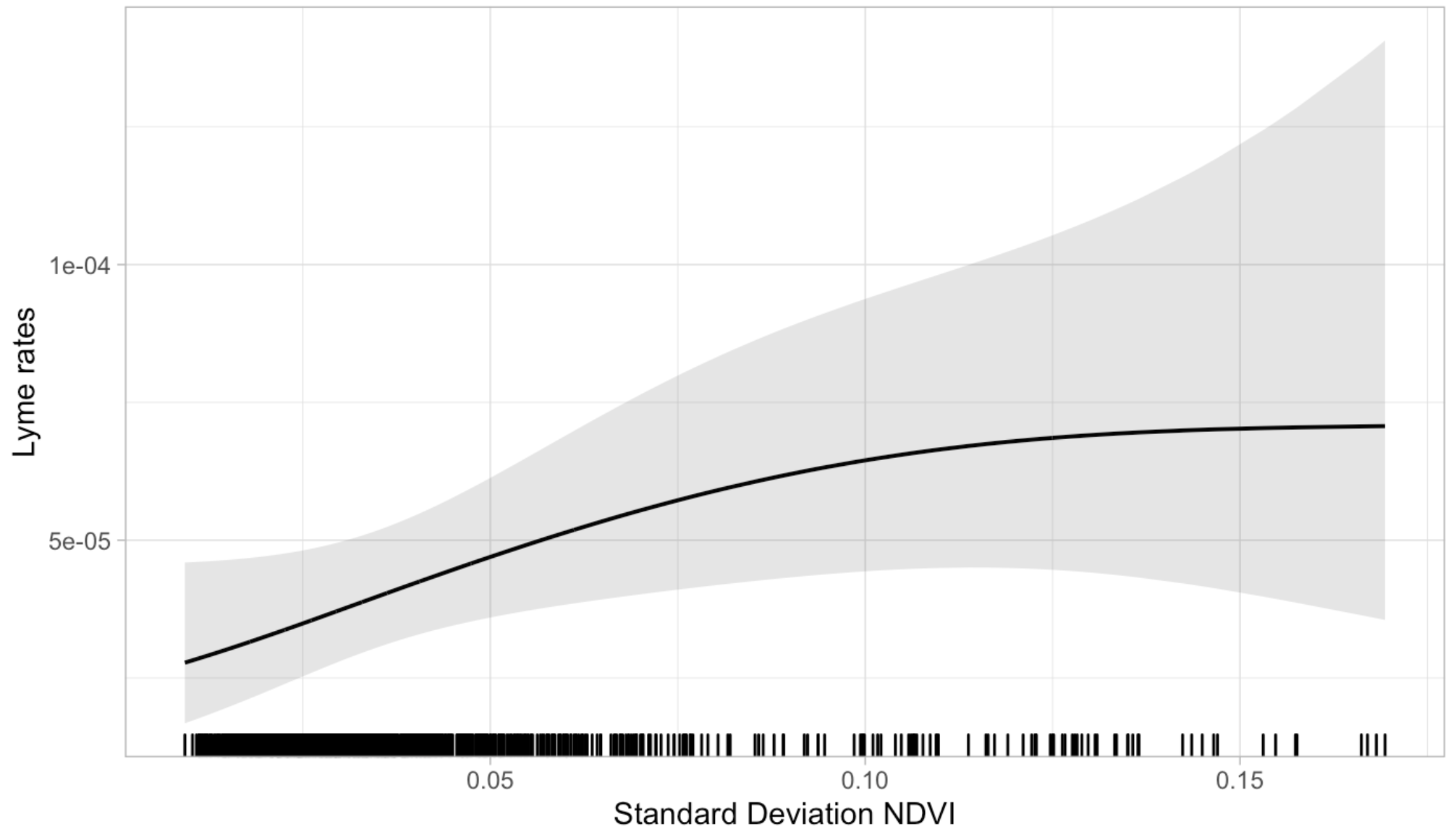

Figure S6O. Wisconsin: Standard Deviation NDVI

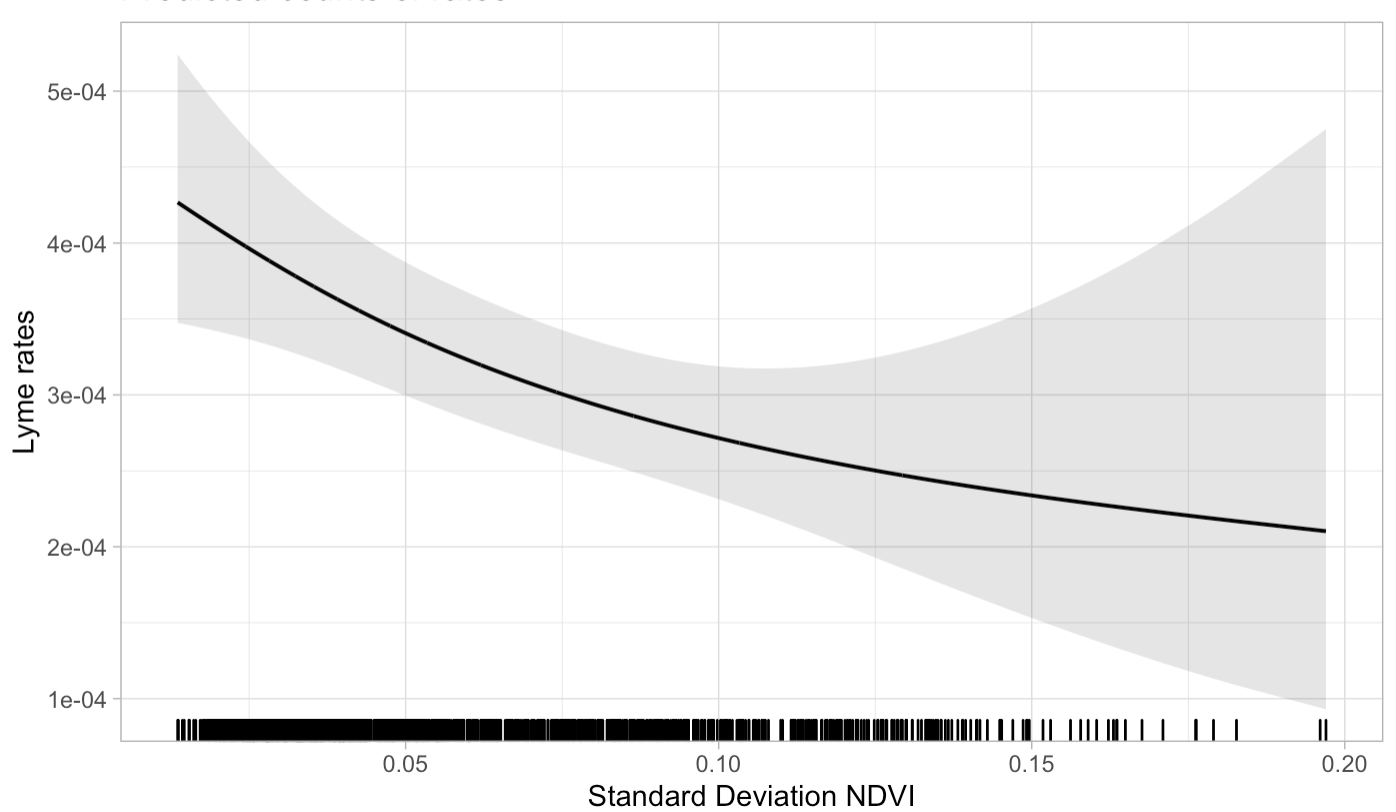
